# Limited overlap between genetic effects on disease susceptibility and disease survival

**DOI:** 10.1101/2023.10.10.23296544

**Authors:** Zhiyu Yang, Fanny-Dhelia Pajuste, Kristina Zguro, Yipeng Cheng, Danielle E. Kurant, Andrea Eoli, Julian Wanner, Bradley Jermy, FinnGen, Estonian Biobank research team, Stavroula Kanoni, David A. van Heel, Genes & Health Research Team, Caroline Hayward, Riccardo E Marioni, Daniel L. McCartney, Alessandra Renieri, Simone Furini, Genomics England Research Consortium, Reedik Mägi, Alexander Gusev, Petros Drineas, Peristera Paschou, Henrike Heyne, Samuli Ripatti, Nina Mars, Andrea Ganna

## Abstract

Understanding disease progression is of a high biological and clinical interest. Unlike disease susceptibility whose genetic basis has been abundantly studied, less is known about the genetics of disease progression and its overlap with disease susceptibility. Considering ten common diseases (N cases ranging from 17,152 to 99,666) across seven biobanks, we systematically compared the genetic architecture of susceptibility and progression, defined as disease-specific mortality. We identified only one locus significantly associated with disease-specific mortality and show that, at a similar sample size, more genome-wide significant loci can be identified in a GWAS of disease susceptibility. Variants that were significantly affecting disease susceptibility were weakly or not associated with disease-specific mortality. Moreover, susceptibility polygenic scores (PGSs) were weak predictor of disease-specific mortality while a PGS for general lifespan was significantly associated with disease-specific mortality for five out of ten diseases. We used theoretical derivation and simulation to propose plausible explanations for our empirical observations and account for potential index-event bias. Overall, our findings point to little similarity in genetic effects between disease susceptibility and disease-specific mortality and suggest that either larger sample sizes or different measures of progression are needed to identify the genetic underpinning of disease progression.

## Introduction

Genome-wide association studies (GWASs) have been successful in uncovering the genetic basis of human diseases by employing a relatively simple study design that compares diseased individuals with controls (Tcheandjieu et al., 2022; Wightman et al., 2021; H. Zhang et al., 2020). This approach is well suited to identify loci associated with disease susceptibility, but it remains unclear whether these results can also inform on the biology of disease progression. Studying the genetic basis of disease progression is relevant for at least two reasons. First, biological insights from the study of disease progression can be more relevant for drug target discovery since many medicines are developed to cure a disease rather than prevent its occurrence. Second, most individuals approach the healthcare system once they develop a disease or its symptoms, and predicting disease progression is in most diseases an important clinical challenge.

In the past years, several GWAS of disease progression have been performed (see **Table S1** for a detailed review), but the number of progression-specific loci discovered has been limited.

In cancer, GWAS have focused on disease survival and have been generally unsuccessful in identifying genome-wide significant signals. For example, a GWAS on breast-cancer survival in over 96,000 patients did not identify any robust association (Escala-Garcia et al., 2019) and failed to replicate two loci found in the previous largest GWAS of breast-cancer survival (Guo et al., 2015). Among neurological conditions, GWAS have focused on disease survival as well as cognitive or motor decline. In one of the largest studies, researchers have identified three novel loci associated with Parkinson’s disease progression (Tan et al., 2022). A recent study on multiple sclerosis progression has identified a locus pointing to involvement of the central nervous system in disease outcome as opposed to the enrichment for immunological-related signals observed for disease susceptibility (Harroud et al., 2023). However, it is worth noticing that older studies of multiple sclerosis outcomes have failed to replicate in larger ones (Pan et al., 2016; Vandebergh et al., 2021). In cardiovascular diseases, studies have focused on disease recurrence, and initial results from the GENIUS-CHD consortium showed the strongest GWAS signal for coronary artery disease was not associated with subsequent events (Patel et al., 2019). In Crohn’s disease, a study has identified four loci for disease progression, indicating distinct genetic contribution from disease susceptibility (Lee et al., 2017).

Apart from single-variant level effects, some studies examined the aggregate effect of many genetic variants. Most of them suggested that polygenic scores (PGSs) for disease susceptibility do not transfer well to disease progression (Barbieux et al., 2019; Lee et al., 2017; G. Liu et al., 2021), although they might outperform other disease-specific biomarkers in the case of cardiovascular diseases (Cho et al., 2023).

Some authors have highlighted the challenges in interpreting results from genetic studies of disease progression due to the bias induced when individuals are selected according to disease status. If common causes of susceptibility and progression are not accounted for, association results can be unreliable due to what is called an index event bias (Yaghootkar et al., 2017) and several approaches to detect and correct for index event bias have been proposed (Dudbridge et al., 2019; Mahmoud et al., 2022).

Large-scale biobanks linked with longitudinal electronic health records have accelerated the research into the genetic basis of disease progression and provide sufficient sample size to answer two key questions: 1) Do genetic predictors that influence disease susceptibility have a similar impact on disease progression? 2) Can we use PGSs for disease susceptibility to predict patients’ disease progression? In this study, we aim to provide empirical and theoretical answers to these two questions by focusing on a specific, but commonly used definition of disease progression: disease-specific mortality (Figure 1). Through an international collaboration across multiple large-scale biobanks, we systematically compared genetic architecture of disease susceptibility and mortality for ten common diseases focusing on both single variant and aggregated polygenic effects.

**Figure 1.**
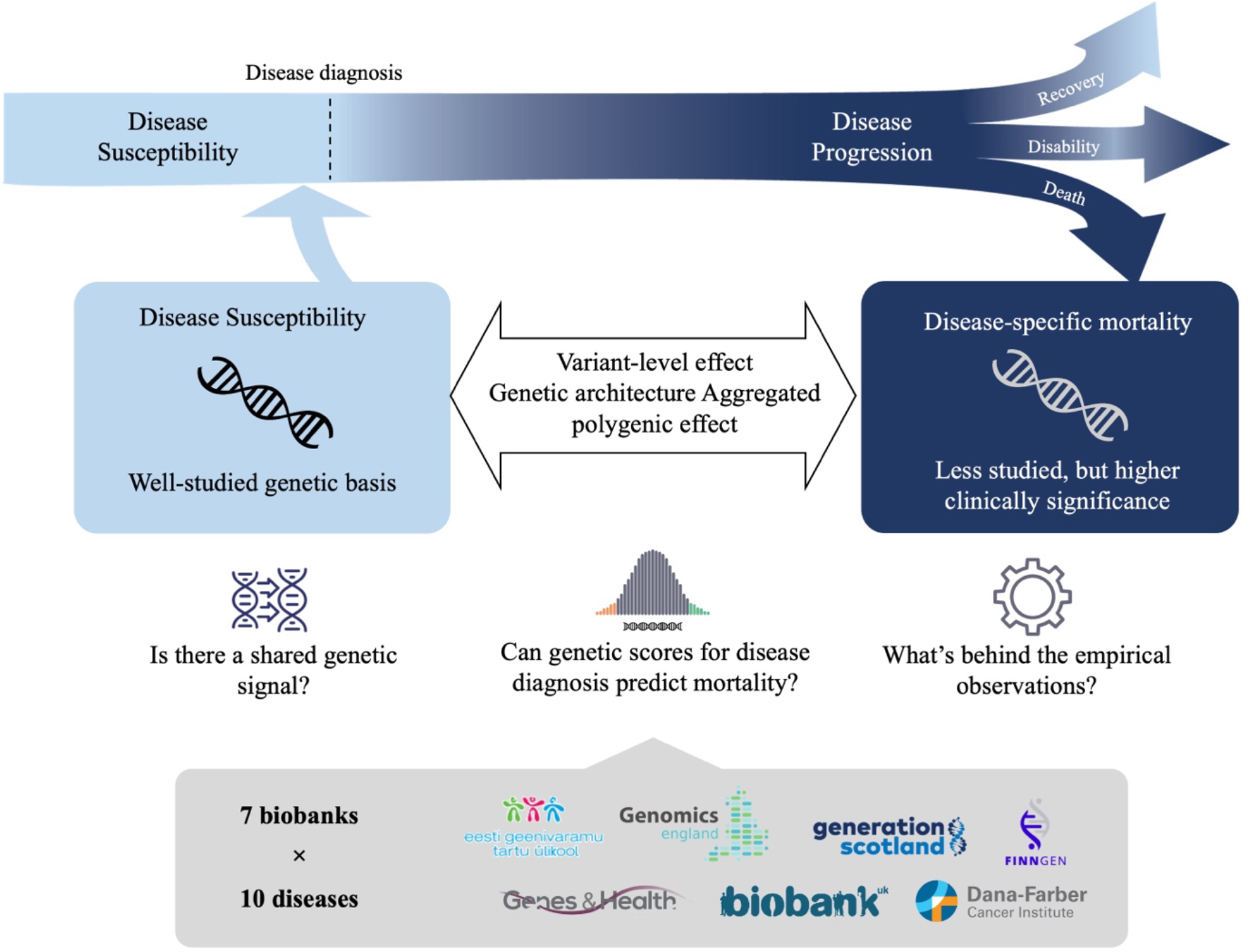
In this study, using data from seven biobanks, we investigated the genetic similarity between disease susceptibility and disease progression, defined as disease-specific mortality. We selected ten diseases and ran GWASs of disease-specific mortality among disease individuals. We then compared the genetic architecture of disease susceptibility and mortality focusing on both single variant and aggregated polygenic effects. Furthermore, we attempted to interpret our empirical observations with simulations and theoretical derivations.

## Results

### Participating biobanks and disease of interest

We considered ten common diseases that substantially increase mortality risk in the general population and have a large public-health impact (**Table 1**). We confirmed disease association with mortality using nation-wide Finnish data and observed a hazard ratio (HR) for 20-years mortality ranging from 1.31 for Type 2 diabetes to 3.61 for chronic kidney disease in females (Viippola et al., 2023) (**Table S2**). We identified diseased individuals based on consistent disease definitions captured via electronic health records or registry data across seven longitudinal studies: FinnGen (Kurki et al., 2023), UK biobank (Bycroft et al., 2018), Estonia biobank (Leitsalu et al., 2015), Generation Scotland (Smith et al., 2013), Genomics England (Turnbull, 2018), Genes & Health (Finer et al., 2020), Dana-Farber Cancer Institute (Gusev et al., 2021) and BioMe. The number of individuals included ranged from 99,666 individuals with type 2 diabetes to 17,152 individuals with Alzheimer disease (**Table 1**). All diseased individuals were followed up for at least three months, with a maximum follow up of 63.29 years in FinnGen. We defined disease-specific mortality based on death certificates where the disease of interest was mentioned as primary or secondary cause of death. One participating biobank did not have information on causes of death, and we used overall death instead (**Supplementary Material**). We observed the highest cause-specific mortality rate for Alzheimer’s disease (28.3%) in FinnGen and the lowest for Type 2 diabetes (3%) in Estonia Biobank.

**Table 1.** Total sample sizes for disease-specific mortality GWAS for each disease and percentage of mortality by year. Also see **Table S2** for details.

| Disease | Sample Size |  | percentage of death within 2y | percentage of death within 5y | percentage of death within 10y |
| --- | --- | --- | --- | --- | --- |
|  | N of disease specific deaths | N diseased individuals |  |  |  |
| Prostate cancer | 3045 | 27682 | 3.02 % | 6.35 % | 9.13 % |
| Breast cancer | 2886 | 34849 | 1.70 % | 4.50 % | 6.59 % |
| Colorectal cancer | 3635 | 17787 | 9.69 % | 17.13 % | 19.73 % |
| Coronary artery disease | 10640 | 89249 | 1.74 % | 3.86 % | 6.79 % |
| Type 2 diabetes | 4886 | 106405 | 0.45 % | 1.20 % | 2.50 % |
| Atrial fibrillation | 3612 | 85824 | 0.92 % | 1.99 % | 3.14 % |
| Chronic kidney disease | 1751 | 30143 | 1.98 % | 3.94 % | 5.47 % |
| Alzheimer's disease | 4659 | 17152 | 5.31 % | 15.50 % | 24.89 % |
| Heart failure | 1757 | 35285 | 1.72 % | 2.92 % | 4.08 % |
| Stroke | 6757 | 89435 | 1.26 % | 2.89 % | 5.05 % |

### Variants affecting disease susceptibility do not have similar effects on disease-specific mortality

For each disease, we carried out a GWAS of disease-specific mortality among disease individuals using Cox proportional hazard model as implemented in GATE (Dey et al., 2022) or SPAcox (Bi et al., 2020) (**Figure S1-10**). On top of all common GWAS covariates, all analyses were also adjusted for age at disease diagnosis for two reasons: 1) Patients’ age is strong predictor of one’s mortality; 2) Age of onset has non-trivial genetic contribution partially overlapping with disease susceptibility (Feng et al., 2020) and we are instead interested in genetic effects on disease-specific mortality.

Out of all ten diseases studied, we only identified one locus associated with disease-specific mortality at genome-wide significant level (*p* < 5x10^−8^). The locus (rs7360523) on chromosome 20, close to *SULF2*, was associated with disease-specific mortality among patients with heart failure.

We asked whether well-established signals for disease susceptibility were associated with disease-specific mortality (**Figure 2**). For each disease, we compared the effect sizes from the largest published GWAS with the result from our GWAS of disease-specific mortality. Using a Bayesian approach (Pirinen, 2023) we could not confidently assign any genetic variant as having the same magnitude of effect on disease susceptibility and disease-specific mortality. In total 888 leading variants were reported from all susceptibility GWAS, whereas none of them was significantly associated with disease-specific mortality after multiple testing correction (*p* < 0.05/888 = 5.63 x10^−5^). Nonetheless, 482 showed the same effect direction, which is marginally more than expected by chance (probability of observing same direction of effect direction 0.54 [95% CI: 0.51 - 0.58], binomial test against 0.5 *p* = 0.01).

**Figure 2.**
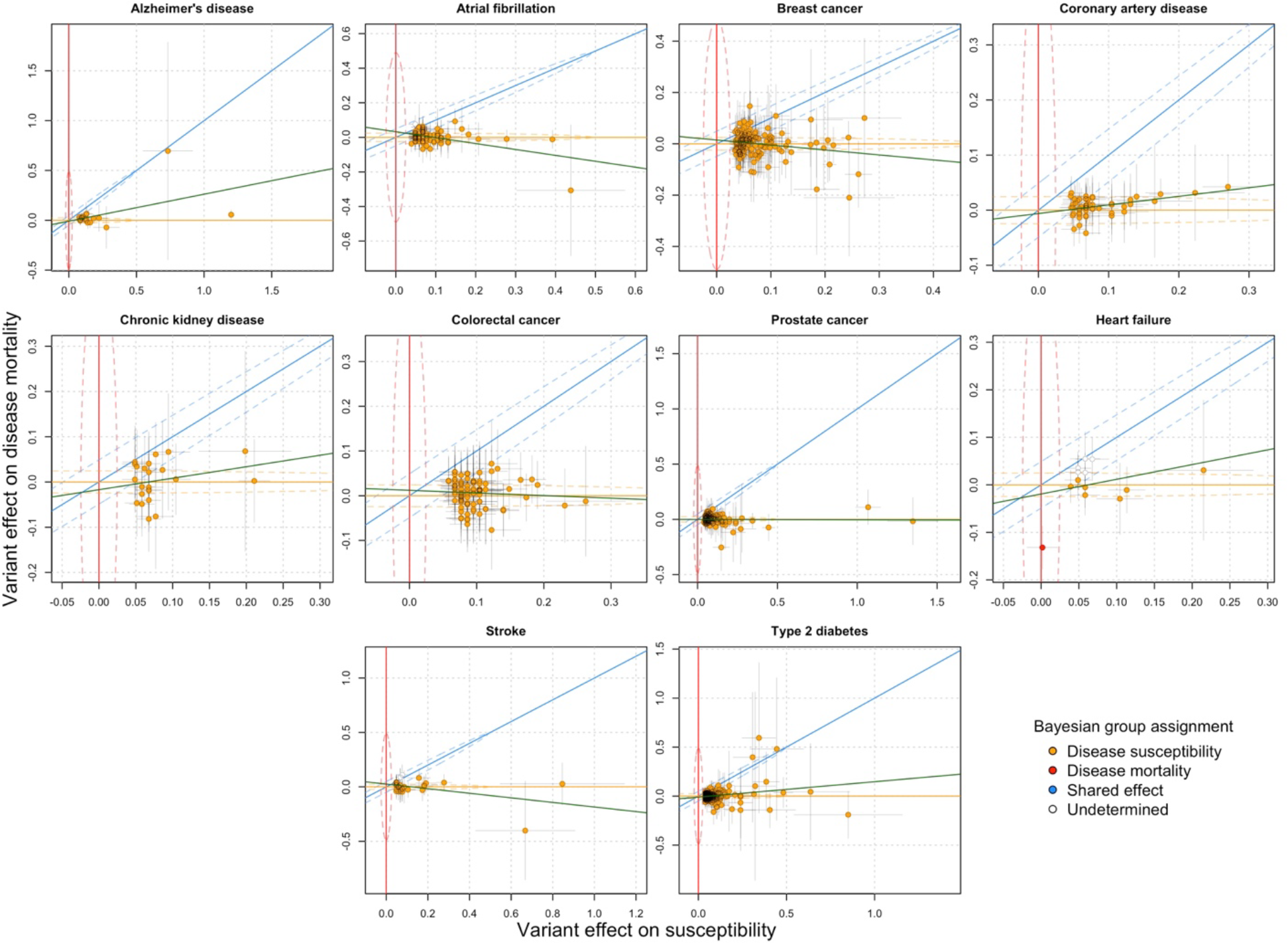
Relationship between variant effects (one for each locus) on disease susceptibility (x-axis) and disease-specific mortality (y-axis). Variants were selected either because genome-wide significance for susceptibility in the largest disease specific GWAS or because genome-wide significance for disease-specific mortality in the current study. Only one locus for heart failure mortality was genome-wide significant. Point colour indicates group assignment for variants (disease susceptibility, disease-specific mortality, or both). Variants with assignment posterior probability > 0.9 are assigned to the group. Variants in white indicate assignment posterior probability is < 0.9 for all the three groups. Posterior probabilities are estimated using R package linemodels (Pirinen, 2023). Red line: x = 0; blue line: y = x; orange line: y = 0; Green line: linear fit for all independent variants in the plot. Dashed lines represent 95% highest probability regions for each group. Also see **Table** S3-S12 for quantitative results.

The only disease-specific mortality locus identified for heart failure also did not show comparable effect on heart failure susceptibility (*p* = 0.87 in susceptibility GWAS with opposite direction of effect). The low number of genome-wide signals for disease-specific mortality were consistent with the lower estimated heritability compared to the GWAS of disease susceptibility (**Table S2**).

### Statistical power does not explain the overall lack of genetic signals for disease-specific mortality

To find out if the overall lack of significant genetic signals for disease-specific mortality was simply due to lower sample size compared to the GWAS of disease susceptibility, we performed a down-sampling experiment in FinnGen and UKBB by imposing the same effective sample size for both analyses. To further make the two analyses comparable, the GWAS of disease susceptibility was conducted using survival analysis with age as time scale and disease diagnosis as outcome. The GWAS of disease susceptibility returned 30 genome-wide significant loci across all diseases, except colorectal cancer and heart failure, while the GWAS of disease-specific mortality returned no genome-wide significant results (**Table 2**).

**Table 2.** GWAS power comparison between disease-specific mortality and disease susceptibility under the same sample size and GWAS model in FinnGen and UK biobank. Number of independently associated genome-wide significant loci. *We report no significant loci for Heart Failure in contrast to what reported in Figure 2 because the GWAS was conducted only in FinnGen and UK biobank.

| Disease | N. disease-specific deaths | N. diseased individuals | N. of GW-significant loci |  |
| --- | --- | --- | --- | --- |
|  |  |  | Disease-specific mortality | Down-sampled disease susceptibility |
| Prostate cancer | 2623 | 24070 | 0 | 8 |
| Breast cancer | 1584 | 27204 | 0 | 2 |
| Colorectal cancer | 2311 | 11986 | 0 | 0 |
| Coronary artery disease | 9067 | 67051 | 0 | 3 |
| Type 2 diabetes | 4169 | 85010 | 0 | 5 |
| Atrial fibrillation | 2943 | 71693 | 0 | 3 |
| Chronic kidney disease | 1283 | 23744 | 0 | 1 |
| Alzheimer's disease | 4659 | 17152 | 0 | 7 |
| Heart failure | 4203 | 62717 | 0 * | 0 |
| Stroke | 1503 | 31414 | 0 | 1 |

### Polygenic scores for disease susceptibility are weak predictors of disease-specific mortality

We investigated the joint effects of genetic variants associated with disease susceptibility in predicting disease-specific mortality. For each disease we constructed a polygenic score (PGS) using results from the largest GWAS of disease susceptibility. All the PGSs were strongly associated with disease susceptibility. The hazard ratios (HR) for 1 standard deviation in the PGS ranged from 1.16 [1.14 - 1.17] for stroke to 1.90 [1.87 - 1.92] for prostate cancer (dashed line in **Figure 3**). On the contrary, the same PGSs were weakly or not associated with disease-specific mortality (orange dots in **Figure 3**). For example, although strongly associated with disease susceptibility, a PGS for breast cancer showed no association with breast cancer mortality (HR = 0.98 [0.93-1.04]). The strongest association was observed between the heart failure PGS and heart failure mortality (HR = 1.09 [1.06 - 1.12]), while the PGSs for chronic kidney disease and prostate cancer trend towards having a protective effect on mortality (HR = 0.95 [0.90 - 1.01] and HR = 0.96 [0.92 - 1.00], respectively).

**Figure 3.**
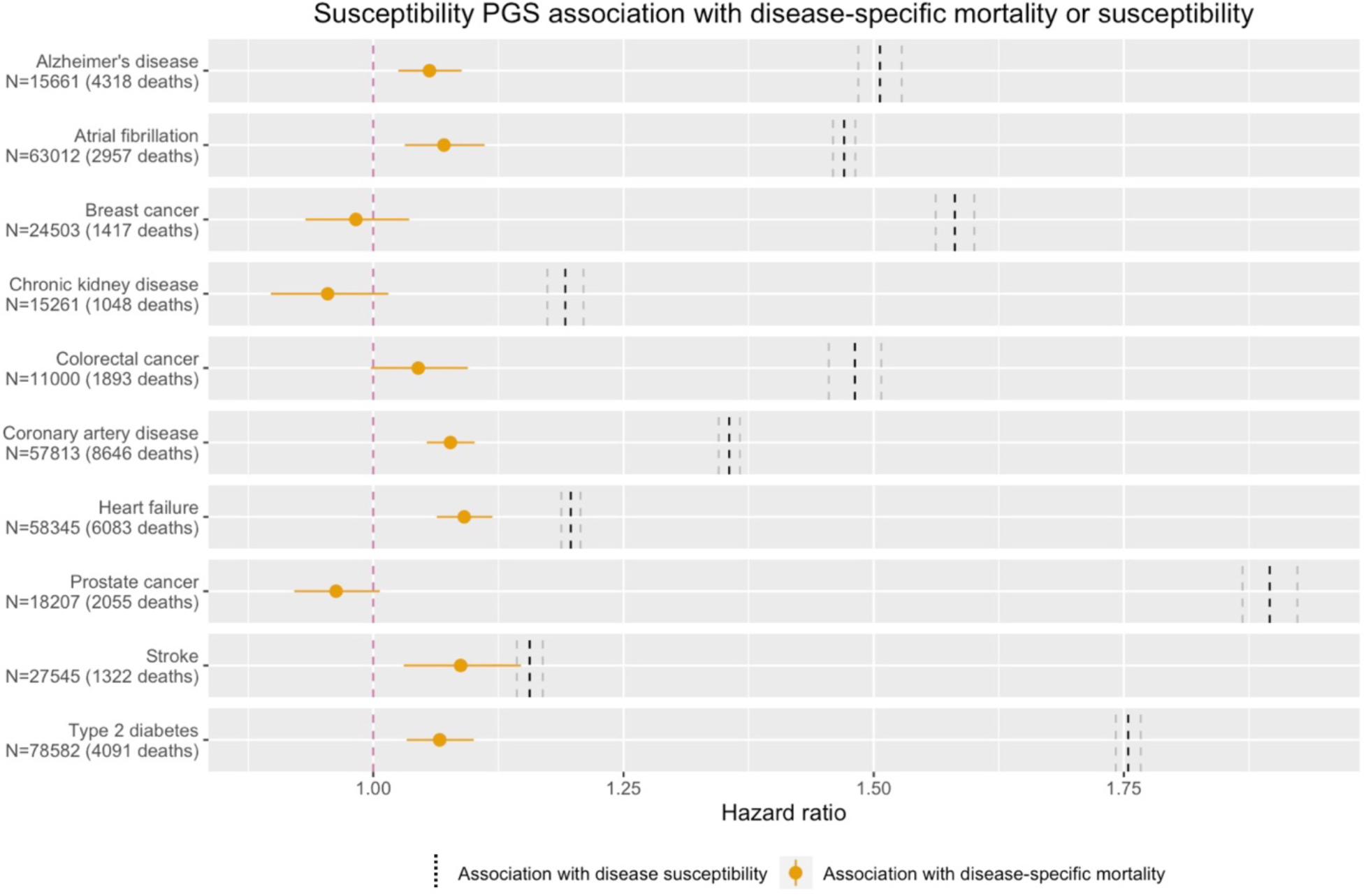
Association between PGS for disease susceptibility and either disease-specific mortality (orange dot) or susceptibility (dashed line). Disease susceptibility PGS was derived from published large-scale GWAS for each disease. PGS associations with both disease susceptibility and disease-specific mortality were carried out using a Cox proportional hazard model. The sample size reported on the y axis refers to the disease-specific mortality analyses, the sample size for association with disease susceptibility can be found in **Table** S13. Horizontal solid lines represent 95% confidence interval (CI). The vertical dashed in black and grey represent association with disease susceptibility HR and 95% CI respectively.

To understand the robustness of these results, we performed a variety of sensitivity analyses. First, we assessed if using a less specific definition of disease progression, namely all-cause mortality, would impact the observed results. We observed significantly larger correlation coefficients of susceptibility PGSs on disease-specific mortality than on all-cause mortality in five out of ten diseases (**Figure S11, Table S15**). Second, we only considered individuals who developed the disease after study enrollment (**Figure S12A, Table S16**) as a way to account for survival bias, which might explain some of the negative associations between PGSs and cause-specific mortality. Nonetheless, results were consistent (correlation coefficient *r* between effect sizes *β* in main analysis and sensitivity analysis = 0.94), and we continued observing a negative association between a PGS for prostate cancer and prostate cancer mortality. Third, we considered different maximum follow-up lengths (2, 5 and 10 years) because we reasoned deaths occurring shortly after disease diagnosis were more likely to be caused by the disease. However, results were overall comparable across follow-up lengths (correlation coefficient *r* between effect sizes in main analysis and sensitivity analysis = 0.68, 0.83 and 0.91 for 2, 5 and 10 years respectively. **S12B, Table S16**) and contrary to our expectation, some diseases (e.g heart failure) showed a stronger association between the susceptibility PGS and disease-specific mortality when considering longer rather than shorter follow-up lengths (effect size *β* = 9.61 x 10^−3^, 0.03 and 0.06 for 2, 5, 10 years respectively).Finally, we evaluated if adjusting the analyses for age at diagnosis could mask an age-specific effect of PGS on cause-specific mortality, for example because such effect was only observed among young or old patients. We observed a significant difference (*p* < 0.005 under Bonferroni correction) for PGS effect on disease-specific mortality between lower and upper 50% quantile diagnosed age groups only for Alzheimer’s disease (**Figure S13, Table S17**). The association between Alzheimer disease PGS and mortality was significant only among younger, but not older patient. Finally, we tested the effect using only unrelated individuals in FinnGen and found the result to be robust (**Figure S14, Table S15**). We have also carried out the same analyses using non-European individuals from Genes & Health. However, due to limited power no conclusion could be drawn (**Figure S16**). See **Figure S15** for forest plot of effects from each participant European biobanks.

### A polygenic score for longevity was significantly associated with disease-specific mortality for five out of ten diseases and showed larger effects than the polygenic score for disease susceptibility

Having established that susceptibility PGS are weakly associated with disease-specific mortality, we reasoned that other PGSs that are better proxies of disease-specific mortality could show stronger associations. First, we consider PGSs constructed directly from our GWASs of disease-specific mortality. For diseases where power allowed, we derived PGSs using weights from the meta-analysed GWAS results from all biobanks except for FinnGen and tested the association between PGS and disease-specific mortality within FinnGen. Surprisingly, none of the PGSs were associated with disease-specific mortality (**Figure S17, Table S18**).

Second, we considered a PGS for general longevity derived from the largest lifespan (Timmers et al., 2019) under the assumption that it might capture some of the genetic effects related to disease survival. The longevity PGS was significantly associated with disease-specific mortality for five out of ten diseases (*p* < 0.005 accounting for the number of diseases tested) and it shows larger HR than a PGS for susceptibility for six out of ten diseases (**Figure 4, Table S14**). For prostate cancer, the association with disease-specific mortality was significantly larger for the longevity than the susceptibility PGS (HR = 1.05 [1.01 - 1.10] vs HR = 0.96 [0.92 - 1.01], *t*-test on effect size differences *p* = 4.41 x 10^−3^).

**Figure 4.**
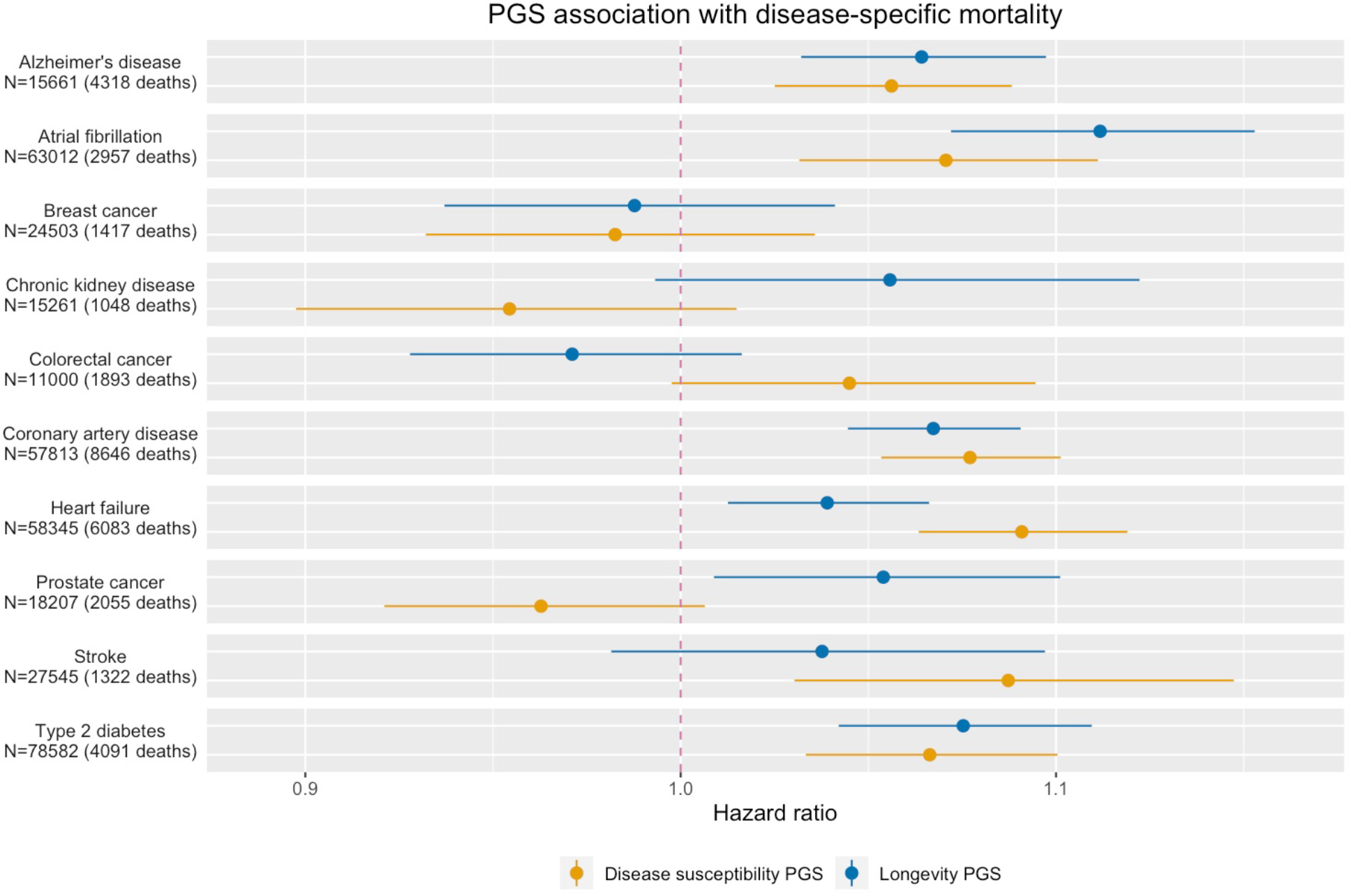
Association between a PGS for disease susceptibility (orange dots) and longevity (blue dots) with disease-specific mortality. Disease susceptibility PGSs were derived from published large-scale GWAS for each disease. Also see **Table** S14 for quantitative results. Longevity PGS was derived from (Timmers et al., 2019). Horizontal solid lines represent 95% CI.

### Theoretical framework and results from simulation suggests the observed results are consistent with low heritability of disease-specific mortality and modest index event bias effect

Towards better understanding of reasons behind our empirical observations, we proposed a simple framework to study the genetic effects on disease susceptibility and progression. We defined the liability to disease susceptibility under a polygenic risk model as random variable *S*

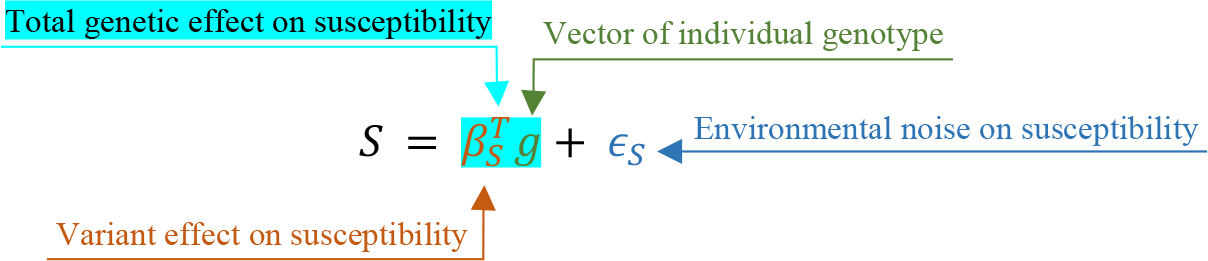

where *g* is the random vector for standardised genotype and *β*_*gi*_ is the random vector of their effect sizes on the diagnosis liability, *ϵ*_*S*_ is the zero mean residual independent to 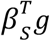.

Next, we defined liability to disease progression as a random variable *P* which depends both on the causal effect of diseases susceptibility (*c*) and some unique genetic effect on disease progression 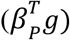

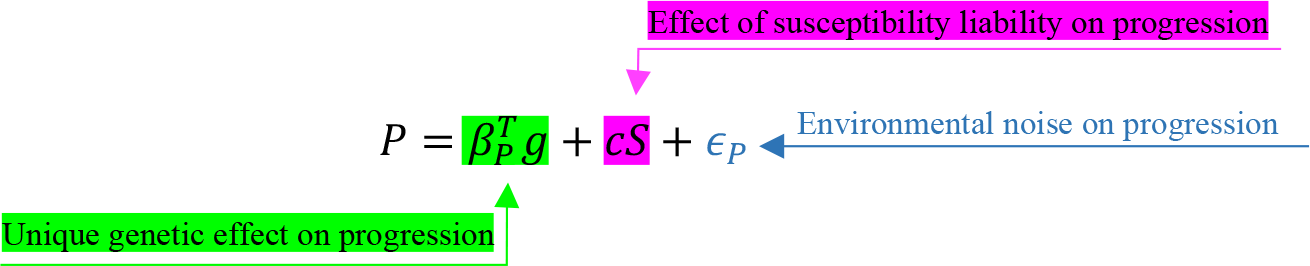

We define the heritability of disease susceptibility (*h*_*sus*_) and *unique genetic components* of disease progression (*h*_*prog*_) as:

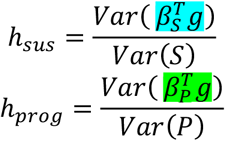

Last, we define ρ as the correlation of the polygenic effect between disease susceptibility and disease progression:

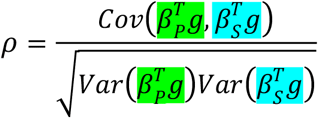

First, we performed simulations by varying two main parameters: *h*_*prog*_ and ρ and compared the theoretical results with the empirical results of **Figure 2**. The empirical results match a scenario with very low heritability of disease progression (**Figure 5**) regardless of the correlation of genetic effects between susceptibility and progression.

**Figure 5.**
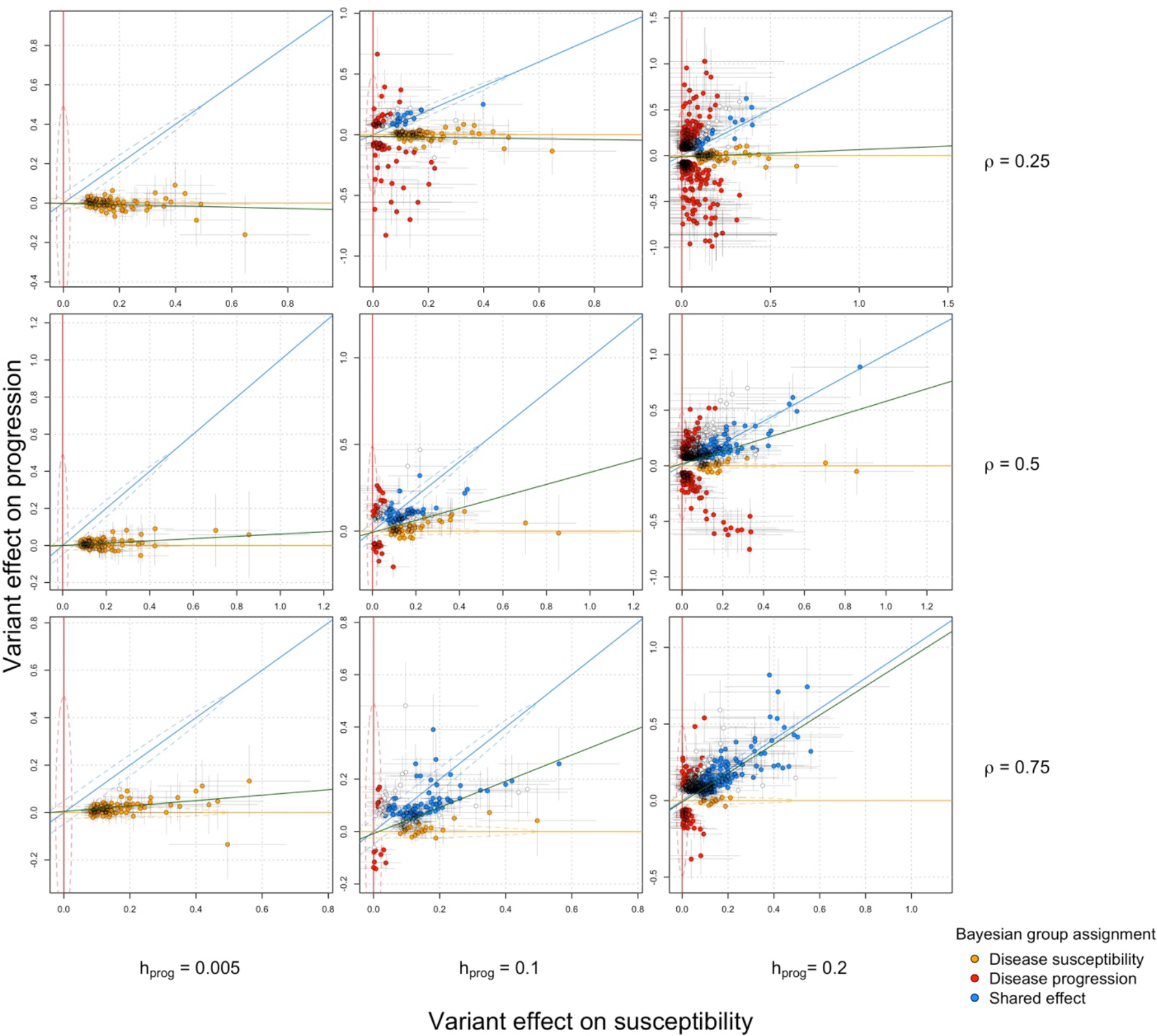
Relationship between variant effects (one for each locus) on disease susceptibility (x-axis) and disease-specific mortality (y-axis) in simulations when varying the heritability of disease progression (*h*_*prog*_) and the correlation of the polygenic effect between disease susceptibility and disease progression (ρ). Variants were plotted if they were genome-wide significant for susceptibility or progression. Point colour indicates group assignment for variants (susceptibility-specific, mortality-specific or both). Variants with assignment posterior probability > 0.9 are assigned to the group. Variants in white indicate assignment posterior probability is < 0.9 for all the three groups. Posterior probabilities are estimated using R package linemodels (Pirinen, 2023). Red line: x = 0; blue line: y = x; orange line: y = 0; Green line: linear fit for all independent variants in the plot. Dashed lines represent 95% highest probability regions for each group.

Second, we derived the theoretical heritability of disease progression (*h*_*prog*_) when measured within patients (**Supplementary material**) and showed a non-monotonic relationship with the genetic correlation (ρ) between disease susceptibility and progression (**Figure S18A**). We also derived how much a polygenic signal for susceptibility (e.g. PGS) can explain the variability in disease progression by calculating theoretical coefficient of determination (R^2^) and showing, as expected, that it increases with ρ (**Figure S18B**). These two results indicate that high correlation in the polygenic effect between disease susceptibility and disease progression can decrease the heritability of disease progression while making the polygenic signals for disease susceptibility a stronger predictor of progression. Finally, we derived the heritability of disease progression outside the within-patient population (**Supplementary material**). This is relevant if one believes that traits measured in the general population (e.g. longevity) can be a proxy of disease progression.

Third, we explored the impact of index event bias by introducing a shared non-genetic risk factor accounting for various proportions of the liability in disease susceptibility and progression. We compared the simulated effect of causal genetic variants on progression with the observed effect from the progression GWAS and found larger differences when the shared non-genetic component accounted for higher liability variance, indicating higher impact of index event bias (**Figure S19**). A correction approach similar to slope-hunter (Mahmoud et al., 2022) reduced the bias improving the concordance with the true simulated effects. However, in the scenario of a low progression heritability (*h*_*prog*_), which is consistent with our empirical findings, index event bias correction showed a limited impact as we observed no genetic variants significantly associated with disease progression before or after bias correction (**Table S19**). Furthermore, we do not see the impact of this correction on posterior variant classification (**Figure S20**).

## Discussion

In this study we systematically explored the overlap of genetic effects on disease susceptibility and a common measure of disease progression, disease-specific mortality, for 10 common diseases. By conducting the largest within-patient GWAS of disease-specific mortality to date we found: 1) leading variants affecting disease susceptibility do not have comparable effect sizes on disease mortality. Rather, they show little effect and no significant association with disease-specific mortality in GWAS ; 2) at a similar sample size, GWAS of disease-specific mortality identified fewer genome-wide significant loci than GWAS of disease susceptibility, suggesting that GWAS of disease progression might require larger sample size or more refined phenotypes than GWAS of disease susceptibility; 3) disease susceptibility PGSs do not transfer well on disease-specific mortality, suggesting that current PGSs are more suitable for identify individuals at high risk of developing a disease rather than those more likely to suffer from the worst consequences. Given the interest in using PGS for optimising clinical trials (Fahed et al., 2022), our results suggest PGS for disease susceptibility might not be the best choice if the trial main outcome is related to disease progression.

Why do we observe limited overlap in genetic effects on disease susceptibility and disease progression? There might be several explanations.

First, genetic influences on disease progression might be too small to detect. External environmental effects such as treatment choice, treatment response, quality and access to care might have a disproportionate impact on disease progression as compared to disease susceptibility, thus limiting the genetic influence. Heterogeneity in patients and their treatments plays a big role in progression for many diseases and we are not currently able to adjust for all that heterogeneity. Using data from clinical trials rather than observational studies and including finer measurements, such as disease relevant biomarkers, can obviate these shortcomings. We also notice that adjusting for age at disease diagnosis does reduce the overlap between susceptibility and progression because variants increasing disease susceptibility are often associated with earlier disease diagnosis (Feng et al., 2020). Previous studies have demonstrated impact of adjusting for age in disease progression analyses (Houlahan et al., 2023) and suggested that association between PGS and measurement of disease progression may be mediated by age.

Second, our definition of disease progression might be a poor proxy for the biological mechanisms impacting disease progression. Our approach aims to compare progression across multiple diseases and comes at expenses of a tailored definition of progression for each disease. Nonetheless, disease-specific mortality has been widely used as a measure of progression (Hernesniemi, 2022; Jabbari et al., 2021; Tan et al., 2022; Wu et al., 2014). In practice, biobank-based studies of disease progression often need to converge to simple definitions to maximise sample size, and disease-specific mortality is an information typically available across biobanks. However, a definition that permits the high data availability might not be one that best reflects the genetic aetiology of a specific disease.

Third, as a common concern for all studies on disease progression, we explored the impact of index event bias resulting from conditioning on diseased individuals. While this is not the main focus of this study, we found that index event bias by itself does not fully explain the lack of concordance between genetic effects on susceptibility and progression observed in our study. Our empirical observations in comparison to various simulations indicate a relatively low heritability for disease progression, defined as disease-specific mortality. Furthermore, heterogenous phenotypes like mortality, although constrained to be disease-specific, can be highly polygenic. In this case, even a perfect correction for index event bias will only be able to recover effect sizes that are not likely to be detected from a progression GWAS. The fact that only one genome-wide locus was picked up from our progression GWAS, indicating low signal-to-noise ratio in the progression GWAS, might be a bigger concern than index event bias. Furthermore, most methods to correct for index-event bias rely on fitting the relationships between variant effects on susceptibility and progression. In our case, this relationship is close to zero and thus the correction will be small and insignificant.

Apart from the impact of index event bias, our theoretical framework reveals some other interesting expectations. The heritability of disease progression is not monotonically increasing with the genetic similarity between susceptibility which implies that the variance in the progression phenotype decreases when disease susceptibility and progression get more genetically similar. This suggests that discovering genetic signals that are specific to disease progression requires a fine balance in the genetic similarity between disease susceptibility and progression and data availability. A simplified definition of disease progression might be useful to increase the sample size but might not be relevant to capture genetic variants specific to susceptibility. On the contrary, when disease susceptibility and progression highly overlap, we can expect homogeneity in patients’ progression, which reduces effective sample size. Once the similarity between disease susceptibility and progression reaches a certain level, it might not be necessary carrying out a GWAS of progression, since a susceptibility GWAS might already capture sufficient information to infer the genetic bases of progression.

Given the aforementioned challenges when conducting GWAS of disease progression among diseased individuals, one attractive alternative would be to study genetic signals for disease progression in a general population and subsequently adapted for within-patients prognostic prediction. For example, PGS for autoimmune conditions derived in the general population are correlated with immune-related adverse events among cancer patients treated with immune checkpoint inhibitors (Groha et al., 2022; Khan et al., 2020, 2021). In our analysis, a longevity PGS derived from GWAS of lifespan was significantly associated with patients’ survival for five out of ten diseases, suggesting patients’ survival could be more affected by general factors related to mortality than disease-specific factors. Methods for cross-trait PGS (Kember et al., 2021) might be leveraged to obtain progression PGS based on existing GWAS results in the general population.

The study has multiple limitations. First, while we explored the similarity in genetic effects between disease susceptibility and disease-specific mortality, we cannot decisively conclude the biological underpinnings to susceptibility and progression are distinct. For example, a phenotype that serves as poor proxy for disease progression will result in attenuated effect sizes, despite genetic variants being causally associated with both susceptibility and progression. Nonetheless the poor replication rate and opposite direction of effect observed for susceptibility signals on disease-specific mortality are consistent with a scenario where at least some variants have no shared effect on both susceptibility and progression. Second, our findings do not necessarily extend outside the diseases explored in this study and further work is needed to confirm the observed trends across more disease categories. Third, whether death certificates are accurately enough capturing primary or contributing causes of death depends on the biobank and healthcare system. We tried to address these concerns by restricting the follow-up duration in sensitivity analyses, reasoning that deaths occurring shortly after disease diagnosis were more likely to be caused by the disease. Fourth, our theoretical framework mostly focuses on the generic relationship between disease susceptibility and progression and does not take the impact of non-genetic factors into account. Also, for simplicity we did not use time-to-event model in this work, which might be more relatable to our empirical experiments.

In conclusion, our current results suggest there is a limited overlap in genetic effects on disease susceptibility and progression, defined as patients’ mortality. Further refinement in inclusion criteria among the patient population and in the definitions of disease progressions can be considered in future studies to robustly identify the genetic underpinning of disease progression.

## Methods

### Selection of diseases

We selected ten common complex diseases spanning various disease categories for the analyses. The diseases are selected to meet following criteria: 1. Have high epidemiological hazard ratio on mortality, so that mortality can be viewed as a reasonable prognosis; 2. Constitute high global disease burden in terms of disability adjusted life years (DALYs) (Abbafati et al., 2020); 3. Relatively common (> 1% prevalence) in population and have reasonable patient bodies in all biobanks; 4. Heritable and have large scale GWAS available to construct PGS. All disease endpoints were defined as a composition of ICD-10 codes curated by the clinical expert groups from FinnGen, Institute for Molecular Medicine Finland (FIMM) and Finnish Institute for Health and Welfare (THL) (Kurki et al., 2023). Same disease definitions, in terms of ICD-10 codes, were adopted by all participating biobanks to the maximum possible extent. See **Table** S2 for list of disease and relevant descriptive statistics.

### Progression definition

For all selected diseases, we defined mortality as our outcome. Precisely, we were interested in both *all-cause mortalities*, namely simple death status of the patient regardless of relevance to the disease, and *disease-specific mortalities*, meaning the death caused directly or indirectly by disease of interest specifically. Disease progression was evaluated as patients’ survival from each type of mortality after being diagnosed with the disease. For all mortality GWASs, we consider only disease-specific mortality whenever it is possible for each participating biobank. Whereas for the PGS analysis, both all-cause and disease-specific mortalities were evaluated. Same as the disease endpoints, cause of death linked to each disease was also curated by clinical expert groups and defined in terms of ICD 10 codes (World Health Organization, 2004). The same definitions were systematically applied to all biobanks to the possible extent. See **Table** S2 for definition of cause-specific mortality for each disease of interest and available sample sizes from each biobank.

### Within-patient mortality GWAS

To achieve variant level effect comparison, for each selected disease, within-patient mortality GWAS was carried out using GATE (Dey et al., 2022) for all biobanks but Generation Scotland, which used SPAcox (Bi et al., 2020) as an alternative. The event of interest in this GWAS was patients’ survival after disease diagnosis. For each disease of interest, GWAS was carried out separately within each ancestry group for biobanks that have a cause-specific mortality event count of 50 at minimum after quality control. Eligible individuals were restricted to patients having a follow-up time after diagnosis of three months (0.25 years) at minimum. We used model below to examine SNP association with patients’ survival:

surv(duration of follow up after diagnosis | disease-specific mortality) ∼ **SNP** + patient’s age of diagnosis + patient’s birth year + sex + PCs + study specific covariates,

where study specific covariates included other available non-heritable biobank specific covariates, such as genotyping chip or batch.

For analyses in the UK biobank, to minimise potential impact of survivor bias, only patients with disease diagnosed after enrollment were considered.

## Results quality control and meta-analysis

After mortality GWAS for selected diseases were carried out within each contributed biobank, we then filtered the resulting summary statistics by imputation INFO scores and minor allele counts. We kept only variants showing an imputation INFO score > 0.7 and having at least 20 minor allele counts for each summary statistics. For GWAS summary statistics with a different human genome build, we used the UCSC LiftOver tool (Kuhn et al., 2013) to convert their genome coordinates into hg38 assembly. Subsequently, for each disease, we meta-analysed GWAS results from each biobank using fixed-effect meta-analysis implemented in METAL (Willer et al., 2010). With which, we also scanned for heterogeneity in effect sizes across different biobanks using Cochran’s Q test. We applied an inverse variance weighted meta-analysis scheme whenever possible. However, since SPAcox does not have effect size or standard error output, in Generation Scotland, we estimated direction of effect under a logistic regression model using plink (Purcell et al., 2007), and subsequently proceeded with a sample-size weighted meta-analysis using the z-scores. This was done for four out of the ten diseases, for which Generation Scotland was one of the data sources: atrial fibrillation, breast cancer, coronary artery disease and type 2 diabetes.

### Variant level effect size comparison

We compared our mortality GWAS results for each disease of interest with large-scale published GWAS on diagnosis of the same disease. For disease diagnosis GWAS, we extracted SNP effects of reported genome-wide significant leading SNPs at independently associated loci from each study. For CKD, a large GWAS on estimated glomerular filtration rate (eGFR) was considered (Wuttke et al., 2019). Specifically, we looked at independent leading SNPs’ effect sizes on binary CKD diagnosis reported from the study so that the scale of measurement is more comparable. For our meta-analysed mortality GWAS, we identify independent genome-wide loci using summary statistics based conditional analysis implemented in GCTA-COJO (Yang et al., 2012). We merged 5,000 Finnish genomes, which is one of the largest GWAS cohorts in this study, with EUR from 1000 Genome as LD reference for this step. Subsequently, for each leading SNP from diagnosis or mortality GWAS, we classify them by linear relationships between their effect sizes in the diagnosis and mortality GWAS into three groups: disease diagnosis specific (slope = 0), disease mortality specific (slope = *inf*) and variants with effect on both diagnosis and mortality (slope = 1). The classification was carried out using a Bayesian framework implemented in R package linemodels (Pirinen, 2023).

### Comparison of genetic architectures for disease diagnosis and mortality

We compared genetic architectures between disease diagnosis and mortality in terms of SNP heritability estimated from the meta-analysed mortality GWAS summary statistics using LD score regression (Bulik-Sullivan et al., 2015). For eligible traits, i.e. traits with non-zero estimated SNP heritability, we further analysed genetic correlation across disease diagnosis, mortality, and general longevity GWAS using the same tool.

### Down-sampled GWAS on age of diagnosis

To ensure heritability comparison between disease susceptibility and progression endpoints not being subject to power issues resulted from difference in sample sizes and GWAS models, for each disease of interest, we also ran time-to-event GWAS to find SNP association with age of diagnosis using a randomly down-sampled cohort which had comparable number of total individuals and event counts as what was available for the within-patient mortality GWAS. The down-sampled GWAS was carried out under model below:

surv(follow-up from birth until diagnosis | disease diagnosis) ∼ **SNP** + patient’s birth year+ sex + PCs + study specific covariates.

This analysis was also carried out using GATE (Dey et al., 2022) but in Finngen and UKBB only, which are two of the largest participating biobanks in this study (See **Table** S2 for biobanks sample sizes).

### Computation of individual level PGS

For each selected disease, we derived variant weights for PGS from GWAS summary statistics listed in **Table** S2 using MegaPRS (Q. Zhang et al., 2021). Heritability contributed by each variant was estimated under the BLD-LDAK model as recommended. For weight estimation, we used the “mega” option which leaves it to the software to decide the most appropriate model given the data. Since we studied mortality, apart from the ten selected diseases, we also computed PGS weights for general longevity using the largest GWAS on lifespan (Timmers et al., 2019). Due to the heterogeneous and polygenic nature of lifespan, for this trait, we used the LDAK-Thin model for SNP level heritability estimation instead. Unlike the BLD-LDAK model used in variant weighting for other diseases, LDAK-Thin model does not take functional annotations into account but estimates SNP heritability only as functions of SNP allele frequencies and local linkage structures. Variant weights were derived for 1,330,820 common SNPs (minor allele frequency > 0.1) lying in the intersection of HapMap3 (International HapMap 3 Consortium et al., 2010) and 1000 Genome (1000 Genomes Project Consortium, 2015) that are available for each GWAS summary statistic.

Once the SNP weights were derived, individual level PGSs for each disease and general longevity were subsequently computed as a weighted sum of effect allele counts using plink (Purcell et al., 2007). Scores were standardised to have 0 mean and 1 as variance within each ancestry group.

### Association between PGS and disease of interest

As a baseline, we first examined if the disease PGSs were associated with their diagnoses. For each selected disease, the association was first tested using a general linear model on case-control status as below:

logit(Pr(Individual is diagnosed)) ∼ **disease PGS** + birth year + sex + PC1-10,

To achieve a fairer comparison with the other experiments, we also evaluated such relationship using a survival model on the age of diagnosis as below:

surv(follow-up from birth until diagnosis | disease diagnosis) ∼ **disease PGS**+ birth year + sex + PC1-10.

The two analyses above were carried out using all eligible individuals in the biobanks. Then for each selected disease, we extracted only the patient group to further conduct the following analyses. To reduce noise in measurements, we limited these within-patient analyses to individuals having a follow-up time of at least three months (0.25 year) after the diagnosis. We tested the association of disease PGSs with our defined prognosis, namely patient survival, using the model below:

surv(duration of follow up after diagnosis | mortality) ∼ **disease PGS** + birth year + sex + PC1-10 + age of diagnosis,

as well as the association of general longevity PGS with patient survival as below:

surv(duration of follow up after diagnosis | mortality) ∼ **general longevity PGS** + birth year + sex + PC1-10 + age of diagnosis.

For both associations, we examined both all-cause mortality and cause-specific mortality within the patient group. All analyses were corrected for gender, except in analyses for breast cancer and prostate cancer, where only female/male individuals were used.

These analyses were carried out independently for each ancestry group within each participating biobank. We only included biobanks where the count of events of interest in the analysed ancestry group was 50 or more. We subsequently meta-analysed effect sizes for the same ancestry group across biobanks using the inverse variance weighted approach.

### Construction of PGS from disease mortality GWAS and effect evaluation within FinnGen individuals

For diseases with sufficient power, we derived mortality PGS weights using meta-analysed mortality GWAS results of European populations from all available biobanks except for FinnGen or Generation Scotland. Apart from FinnGen which was used as a test cohort, we also left out results from Generation Scotland for this analysis because their summary statistics did not have effect size or standard error and therefore cannot be used for inverse-variance weighted meta-analysis, which returns necessary statistics for weight derivation. After deriving PGS weights using MegaPRS (Q. Zhang et al., 2021), we subsequently computed individual level disease mortality PGS for patients of each corresponding disease within FinnGen cohort. The weights and scores are computed in the same manner as mentioned in section **Computation of individual level PGS**. We evaluated effects of these scores on predicting patients’ disease mortality in FinnGen using the model below:

surv(duration of follow up after diagnosis | mortality) ∼ **disease-mortality PGS** + birth year+ sex + PC1-10 + age of diagnosis

### Sensitivity analyses for PGS experiments

We ran a series of sensitivity analyses in eligible biobanks to ensure our observations on the PGSs association were robust, under considerations listed below. Similarly, analyses were carried out per eligible ancestry within each biobank and then meta-analysed.

First, to demonstrate the impact of relevance between disease progression and susceptibility as shown in our theories, we examined the association between susceptibility PGS and all-cause mortality and compared the results with disease-specific mortality in FinnGen. See **Figure** S1 for this result.

We then consider other factors that may bias the results.

- Survivor bias

Depending on each biobank’s recruitment scheme, some patients were diagnosed before the start of their follow-up, which may lead to biassed results due to survivor effect. Therefore, we also ran these analyses for each disease using only samples enrolled before their first onset of the disease of interest. See **Figure** S12A for this result.

- Relevance between cause of mortality in death certificate and disease diagnosis

In this study, we aimed to define disease progression as accurately as possible by focusing our analysis on disease-caused mortality. However, some national death registries may not precisely capture the immediate cause of death, and some mortalities, while documented with the disease as one of causes, may not be truly relevant to the diagnosed disease. To address this concern, we ran the same analysis using only patients with a restricted maximum follow-up length, since death taking place reasonably sooner after being diagnosed might have more to do with the diagnosis, compared to death taking place decades after. Under this consideration, we varied the maximum duration of follow-up after diagnosis by 2, 5 or 10 years. The minimum is still 0.25 years for this analysis. See **Figure** S12B and **Table** S16 for this result. Also see **Table** S2 for sample size breakdown by duration of follow-up in each biobank. As a measurement for comparability between results, we reported the regression coefficients for PGS effect sizes on ten diseases between each sensitivity analysis and main results.

- The effect of diagnosed age

As shown above, we have included age of diagnosis as one of the covariates in all within-patient main analyses models in order to specifically investigate PGSs’ unique genetic effect on disease progression by correcting for the diagnosis. As one of our sensitivity analyses, we also analysed the role of these diagnosed ages in more detail. We repeated all the within-patient analyses for each disease by stratifying patients into early onset and late onset group using 50% age of diagnosis quantile as a cutoff and compared the PGS effects across the two groups. See **Figure** S13 and **Table** S17 for the result.

- Sample relatedness

We included all eligible individuals of each biobank in our main analysis, and one may argue that could impact our effect size estimates. Therefore, we ran the same analysis in Finngen with up to second degree relatives removed. See **Figure** S14 and **Table** S15 for this result.

- Results from non-European populations

Since only patients were considered for most of our analyses, although some of the biobanks, e.g. UK biobank and BioMe, were known to be rather diverse, we winded up with enough power for main results only for the European super population. Nevertheless, comparison of results with other less powered, but available populations can be found in **Figure** S15 for reference.

Forest plot for effects from each biobank is presented in **Figure** S16.

## Theoretical framework

### Setup and notations

We start by defining the liability of the endpoint of the disease susceptibility as the random variable *S* using a simple polygenic risk model, following the lines of (Hujoel et al., 2020):

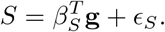

In the above expression, **g** is an *m ×* 1 random vector of standardized genotypes^1^ and *β*_**S**_ is a *sparse m ×* 1 zero-mean random vector of variant effect sizes on the liability of disease susceptibility. Additionally, *ϵ*_*S*_ is the zero mean *residual error* vector that is independent from *β*_*S*_**g** and includes non-genetic effects and environmental noise in disease susceptibility liability. (We use the subscript *S* in the above vectors as a reminder that the respective variables correspond to the liability of disease **s**usceptibility.) Therefore, *β*_*S*_**g** is a zero mean random variable, from which it follows that its expectation is zero, namely

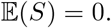

We now define the variance of *β*_*S*_**g** as

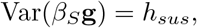

where 0 *≤ h*_*sus*_ *≤* 1. For simplicity, we normalize the variance of the random variable *S* to be equal to one, i.e., Var(*S*) = 1. Intuitively, this normalization implies that *h*_*sus*_ can be interpreted as the *heritablility* of the disease susceptibility endpoint.

Similarly, we can define the liability of the disease progression endpoint, which, in our work, is the mortality due to the disease, as the random variable *P* :

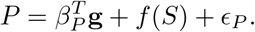

In the above expression, *β*_*P*_ is the zero-mean random vector of direct variant effect sizes that play a role *specifically* on the disease progression liability. Also, *ϵ*_*P*_ is the zero mean random vector that models residual error that is independent of 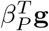 and any term of *f* (*S*) in the liability of the disease progression. Since disease progression is clinically considered as a continuation of the development of the same disease, we believe that it is reasonable to assume that the liability of disease susceptibility will also play a role on its progression. Therefore, we added the term *f* (*S*) in the liability of disease progression to introduce a *causal* contribution for the disease susceptibility liability. To the best of our knowledge, this function *f* (*S*) has not been studied or quantified in prior work. Therefore, for the sake of simplicity and in the absence of prior models, we assume that *f* (*S*) it as a simple linear function, namely

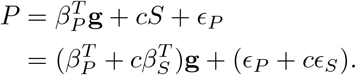

In the above, *c* is a constant and we assume that the effect of disease susceptibility liability on the progression is non-negative, from which it follows that *c* ≥ 0. Recall that *β*_*P*_, *ϵ*_*P*_, and *S* all have zero mean, implying that 𝔼 (*P*) = 0. Let the variance of the progression of the genetic component 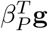 be defined as follows:

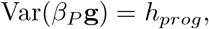

where 0 *≤ h*_*prog*_ *≤* 1. Again, we normalize the variance of the random variable *P* to be equal to one, i.e., Var(*P*) = 1, which can be achieved by placing constraints on the constant *c* and the error term *ϵ*_*P*_. In this case, *h*_*prog*_ can be interpreted as the *unique heritability* of the disease progression. Given our normalization, it follows that *c≤*1. As a corner case, when *c* = 1, *ϵ*_*P*_ and *h*_*prog*_ must both be equal to zero, which implies *P* = *S*. This special case does not merit any further consideration and in the upcoming section we will only focus on 0 *≤ c <* 1.

We note that the random variables *S* and *P* as defined in this section indicate the liability of the two endpoints of interest for a particular disease in *general* populations. However, in practice, the study of disease progression focuses only on the patient group. Such “within-patient” measurements on a continuous scale can be viewed, at least conceptually, as the liability of disease progression conditioned on the liability of disease susceptibility. Therefore, we regress the liability of the progression (random variable *P*) on the liability of disease susceptibility (random variable *S*) to get a continuous, within-patient, progression measurement *P*|*S*:

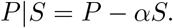

The regression coefficient *α* can be analytically derived as follows:

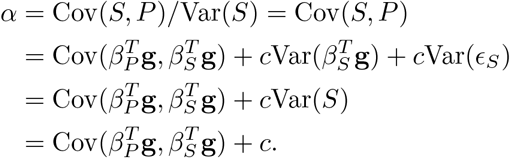

Therefore the within-patients progression liability can be expressed as follows:

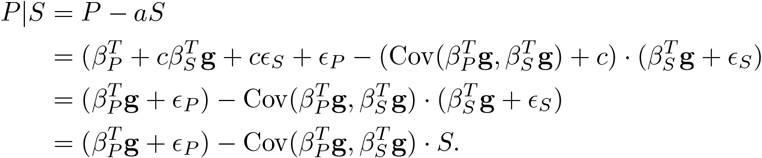

Let *ρ* denote the correlation coefficient between 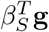 and 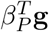. Then,

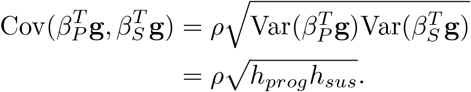

Finally, the within-patient disease progression liability *P*|*S* can be simplified as follows:

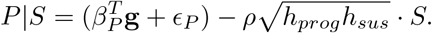

### The *ρ* = 0 case

In order to gain intuition for the more complicated analyses that will follow, we start by looking at the simple (yet admittedly unrealistic) case where there is no genetic correlation between the disease susceptibility and the progression due to the *unique* genetic factors. Mathematically, we assume that the covariance between (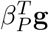 and 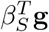 is equal to zero, i.e., *ρ* = 0. In this case, the regression parameter *α* is equal to *c* and

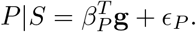

### Heritability of within-patient disease progression

In this case, *P*|*S* has a simple genetic component given by 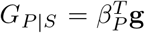. Therefore, its heritability can be

expressed as

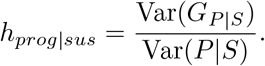

In the above,

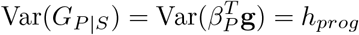

and

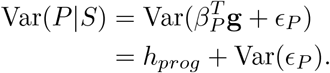

Recall that *P* is normalized so that Var(*P*) = 1. Therefore, 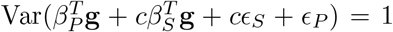. Since we assumed that 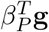 and 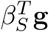 are independent, all terms of *P* are pairwise independent and

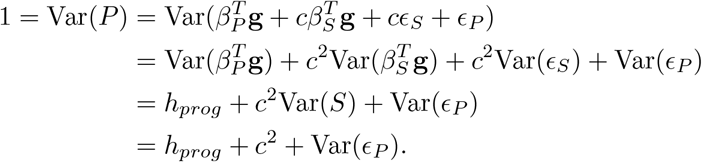

Thus,

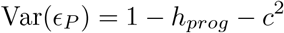

and

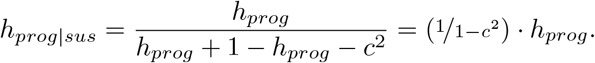

Recall that 0 *≤ c <* 1, which implies that 1*/*(1 *− c*^2^) *≥* 1. Therefore, when *c* approaches one, the constant before *h*_*prog*_ increases rapidly. Intuitively, in this case, the progression endpoint liability depends heavily on susceptibility and little phenotypic variability among patients remains. However, the constraint on *h*_*prog*_ (1 *− h*_*prog*_ *− c*^2^ *>* 0) implies that *h*_*prog*_ will also have to be small in this case.

### Using disease susceptibility genetics to understand “within-patients” progression

In our empirical evaluations using simulated data, we try to explore the association between between Polygenic Scores (PGS) derived from the respective disease susceptibility GWAS and the patient disease progression, as characterized by mortality. Equivalently, using the parlance of the previous sections, we are trying to explore the extent to which the “within-patients” progression can be explained by the genetics of the disease susceptibility endpoint.

Within our framework, we can theoretically answer this question. Let the genetic component of disease susceptibility liability be denoted by 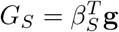. We can then look at the correlation coefficient *R*^2^(*G*_*S*_, *P*|*S*) using the variance-covariance ratio:

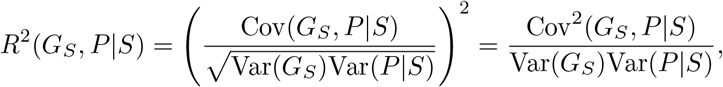

where 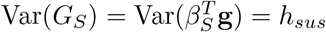 and Var(*P*|*S*) = 1 *− c*^2^. Assuming *ρ* = 0, we get

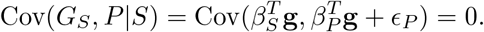

As expected, in this setting, the genetics of disease susceptibility cannot be used to explain the “within-patient” disease progression.

### Using population-level disease progression genetics to understand “within-patients” progression

In our empirical evaluations using simulated data, we also tried to look at the behavior of general longevity PGS derived from lifespan GWAS (Timmers et al., 2019). Recall that in our work we defined mortality as the event of interest, which we will use as a proxy for genetics of the disease progression measure in the general population instead of only within the patient group. We are aware that getting a real “liability of disease progression measure in the general population” is usually impossible in practice: in most cases, a disease progression is not defined for individuals who do not have a disease diagnosis in the first place. This is only possible in special cases where the progression is defined in a general manner. For example, if instead of disease specific mortality, mortality due to *any* cause is defined as disease progression^2^, we can get the genetic determinants of this progression from a longevity GWAS for the general population instead of just the patient group.

In our proposed framework, we can simulate such measurements by defining the liability of the disease progression in the general population as *P*. Then, the within-patient progression liability is defined as *P*|*S*. Therefore, the genetics of the disease progression measure in the general population can be analyzed via *P*, which includes both the unique component of the disease progression as well as the contribution of the disease susceptibility, namely

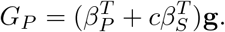

Association between the longevity PRS and patients’ survival is akin to asking about the amount of variance of the within-patients disease progression that is explained by the genetics of the progression liability assessed in the general population. We can theoretically estimate this value using the correlation coefficient *R*^2^(*G*_*P*_, *P*|*S*) as follows:

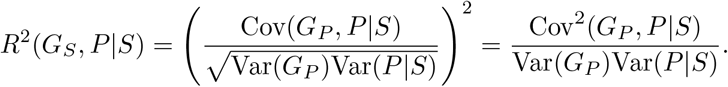

Assuming that 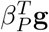 and 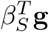 are independent, we get

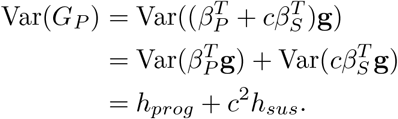

Similarly,

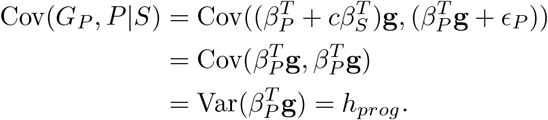

Using the above, we can now express the correlation coefficient *R*^2^(*G*_*P*_, *P*|*S*) as follows:

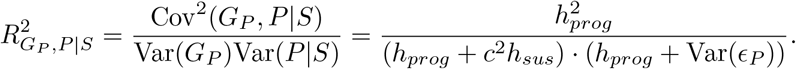

In the extreme corner case *c* = 0, i.e., when disease susceptibility and progression are two completely independent endpoints, we get

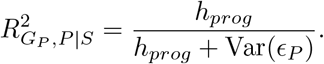

In this case, we get perfect correlation 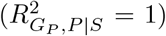 if Var(*ϵ*_*P*_) = 0, because assuming *c* = 0 and *ρ* = 0, disease susceptibility and progression are completely orthogonal events. Therefore, population-level disease progression genetics is equivalent to within-patient progression genetics, which predicts the progression status perfectly in the absence of noise.

### Genetic correlation across within-patient disease progression, progression in general population and disease susceptibility

Finally, we can estimate the correlation between the genetic component of the “within-patient” disease progression 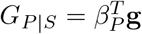, the population disease progression liability 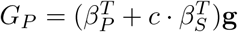, and the genetics of the disease susceptibility 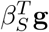. In the case where *ρ* = 0, it is easy to show that the genetic correlation *ρ*(*P*|*S, S*) between *P*|*S* and *S* is

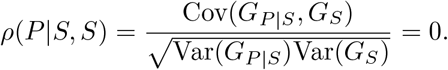

We compute the genetic correlation *ρ*(*P*|*S, P*) between *P*|*S* and *S* as follows:

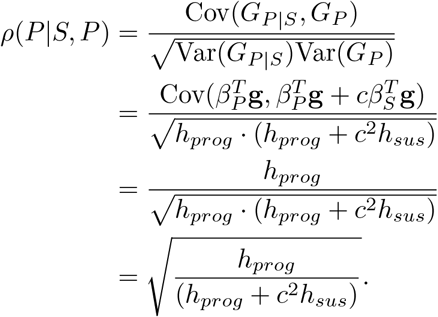

The genetic correlation *ρ*(*P, S*) between *P* and *S* can be computed as:

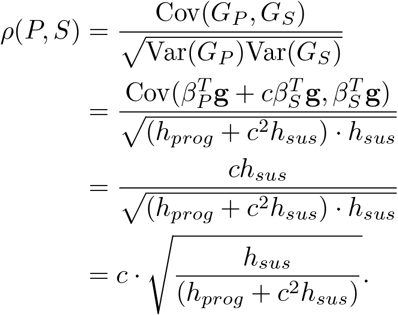

We note that when *c* = 0, i.e., the disease susceptibility is completely independent of the progression, we get *ρ*(*P, S*) = 0, as expected. Similarly, when *c* = 1, it follows that 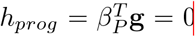^3^ and we get *ρ*(*P, S*) = 1, again as expected.

### The *ρ ≠* 0 case

We now proceed to discuss the more interesting *ρ ≠*0 case. The resulting formulas are analogs of the formulas in Section, albeit more complicated to account for non-zero *ρ*.

### Heritability of within-patient disease progression

Consider the case where 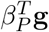 and 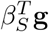 are not independent. In this setting, the genetic component for the within-patient disease progression will be

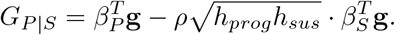

Its variance can be computed as follows:

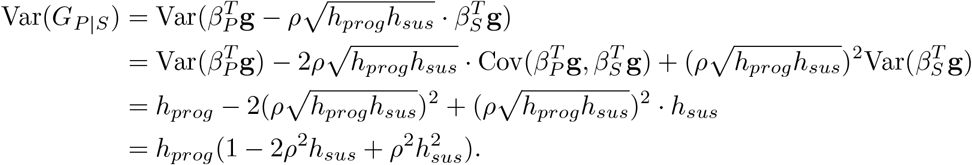

Similarly, its phenotypic variance will be:

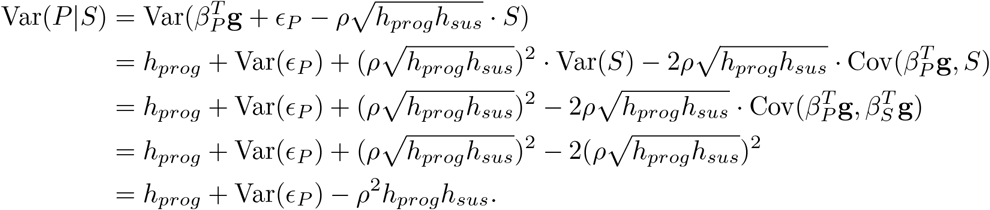

Next, in order to compute the variance of *ϵ*_*P*_, i.e., Var(*ϵ*_*P*_), we need to look at the variance of the disease progression liability defined in the population. Recall that, by our normalization assumptions, Var(*P*) = 1. Therefore,

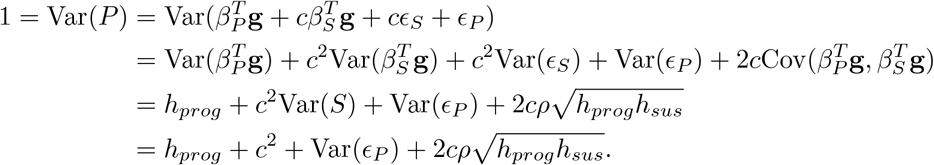

We can now conclude that

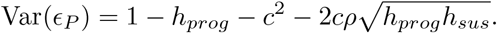

Using the above equation, the phenotypic variance of within-patient disease progression can then be expressed as:

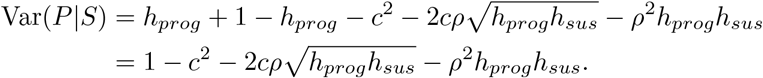

Finally, its heritability can be expressed as

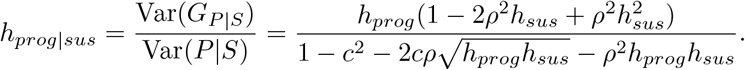

### Using disease susceptibility genetics to understand “within-patients” progression

We again consider the correlation coefficient *R*^2^(*G*_*S*_, *P*|*S*):

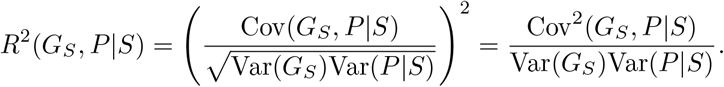

In the above, 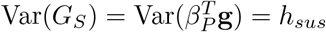 and

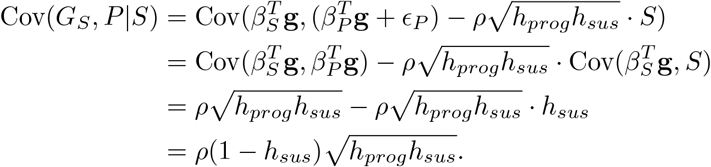

We can now use the expression for Var(*P*|*S*) derived in the previous section to get:

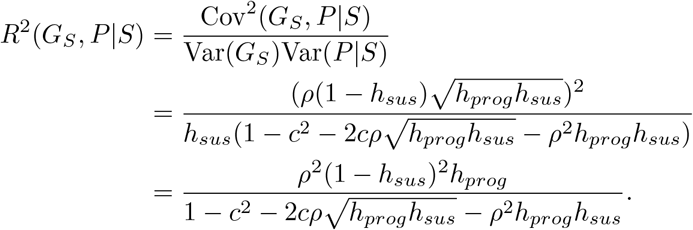

### Using population-level disease progression genetics to understand “within-patients” progression

We now express the “within-patient” group disease progression variance that is explained by the genetics of the progression liability as measured in the population. Formally, *R*^2^(*G*_*P*_, *P*|*S*) can be expressed as:

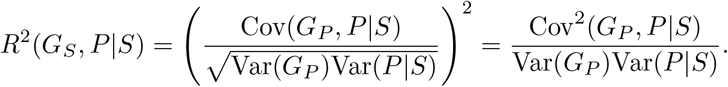

The variance of the genetic component for the “within-patient” group disease progression is:

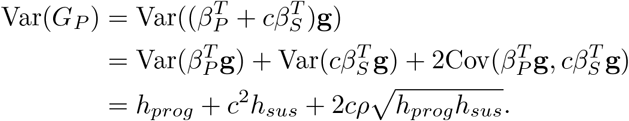

Similarly, the covariance Cov(*G*_*P*_, *P*|*I*) is

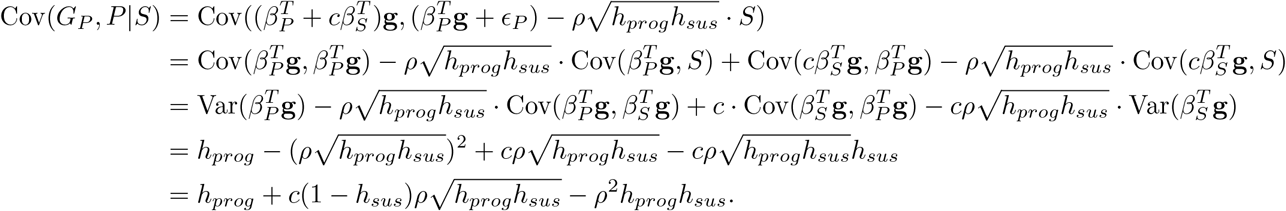

Finally, the correlation coefficient *R*^2^(*G*_*P*_, *P*|*S*) can be expressed as follows:

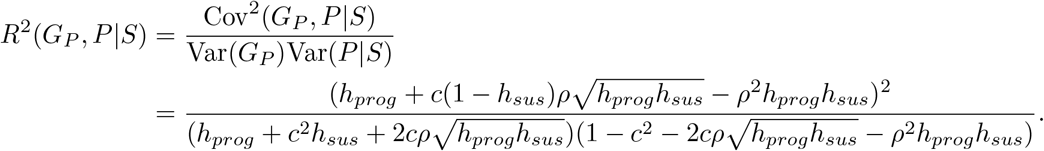

Note that if *c* = 0 and *ρ* = 0, i.e., the liability of disease progression and the susceptibility are uncorrelated, the correlation coefficient *R*^2^(*G*_*P*_, *P*|*S*) = *h*_*prog*_, which is exactly equal to the heritability of the progression liability.

### Genetic correlation across in-patient progression, progression endpoint in population, and susceptibility

Similar to the *ρ* = 0 case, we can estimate the correlation coefficient between the genetic component of the “within-patient” group disease progression (in our notation, this component is 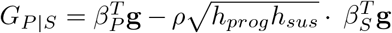, the genetics for progression liability in the population (in our notation, 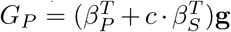, and the disease susceptibility genetics (in our notation, 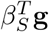).

Recall that, by definition, *G*_*P* |*S*_ has a variance equal to

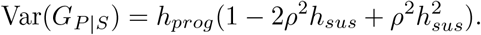

Also,

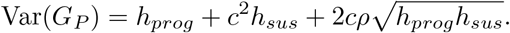

Also, by definition, Var(*β*_**gi**_**g**) = *h*_*sus*_. We can now compute the genetic correlation *R*(*P*|*S, S*)) between *P*|*S* and *S* as follows:

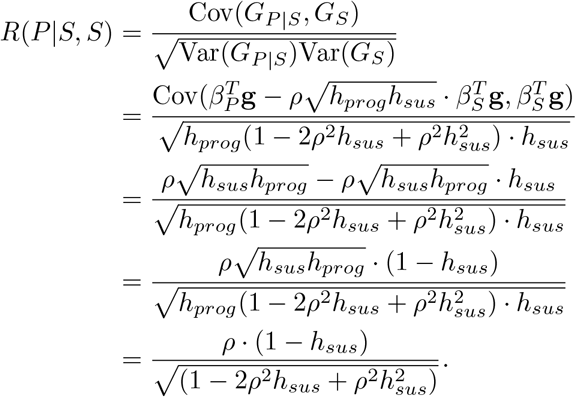

The genetic correlation *R*(*P*|*S, P*) between *P*|*S* and *P* is:

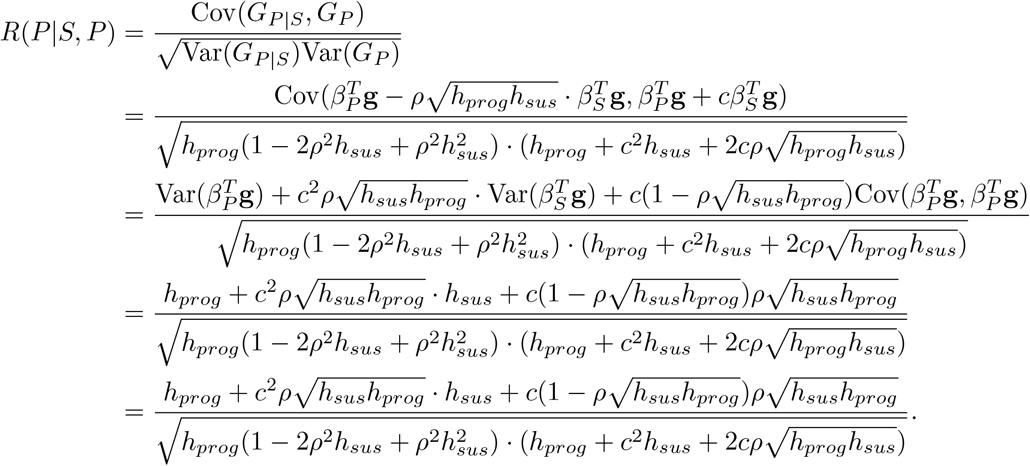

Finally, the genetic correlation *R*(*P, S*) between *P* and *S* is:

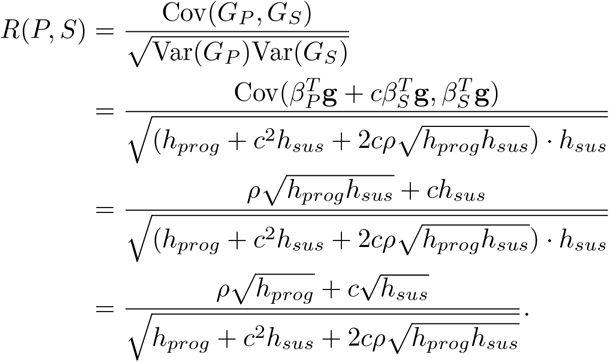

We note that if *c* = 0, then *R*(*P, S*) = *ρ*; if, in addition, *ρ* = 1, then *R*(*P, S*) = 1 as well.

### Estimating variant-level effect sizes for in-patient progression

As part of our empirical assessment using real and simulated data, we ran disease mortality GWAS within the patient groups in order to compare variant level effects between disease susceptibility and within-patient disease progression. In our empirical evaluations, we found it difficult to replicate SNPs that are strongly associated with disease susceptibility in the mortality GWAS. A naive explanation for such lack of concordance is the poor definition of the disease progression endpoint as well as the low genetic similarity between disease susceptibility and disease-specific mortality. We believe that there are deeper and more significant reasons for the aforementioned issue. Indeed, in this section, we will show that beyond real genetic distinctions between the two endpoints, the so called *index event bias* (Yaghootkar et al., 2017) can also play an important role. Intuitively, this bias arises naturally from the design of prognostic research which focuses on within-patient assessments. This bias can result in attenuation of effects even for variables that have exactly the same underlying effect size on disease susceptibility and progression in the population.

We start by discussing two, somewhat minor, notational modifications that we will use in the remainder of this section compared to our work in the previous sections. *First*, our analysis will focus on a *single* variant instead of additive polygenic effects. Therefore, *g* is no longer a vector variable, and neither are *β*_*gp*_ and *β*_*gi*_. Similarly to the previous sections, *g* is still standardized to satisfy E(*g*) = 0 and Var(*g*) = 1. *Second*,, we introduce the standardized variable *u* with E(*u*) = 0 and Var(*u*) = 1 that is independent from *g*, as well as from the error terms *ϵ*_*i*_ and *ϵ*_*p*_; this variable *u* accounts for all other causal factors that could be shared by the disease susceptibility and the progression in the population. This new variable *u* has direct effects denoted by *β*_*ui*_ and *β*_*up*_ on the disease susceptibility and progression liability, respectively. As a composite variable, *u* can contain genetic effects from other variants beyond the one captured by *g* as well as common non-genetic effects.

In light of the above discussion and notations, we now model the random variables *S, P* as:

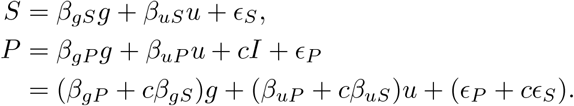

In a disease progression GWAS, we are usually interested in the unique genetic effect on disease progression, namely the quantity *β*_*gP*_. However, in a within-patient GWAS, we are interested in measuring *g*’s observed genetic effect on disease progression, conditioned on disease susceptibility. We denote this within-patient observed genetic effect as 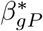. Then, the expectation of the within-patient disease progression can be expressed as

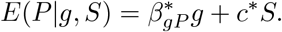

In the above equation, *c*^*\**^ is the observed effect size from the disease susceptibility liability as modelled by the variable *S*. Using least-squares to estimate 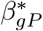 and *c*^*\**^, we get:

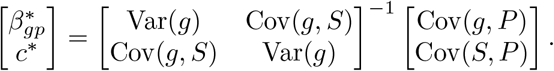

Recall that *g, u, S*, and *P* are all standardized random variables and that *ϵ*_*S*_ and *ϵ*_*P*_ satisfy E(*ϵ*_*S*_) = E(*ϵ*_*P*_) =0.Moreover, *g, u, ϵ*_*S*_, and *ϵ*_*P*_ are all pairwise independent. We now analytically compute each term in the above equation, starting with Cov(*g, S*):

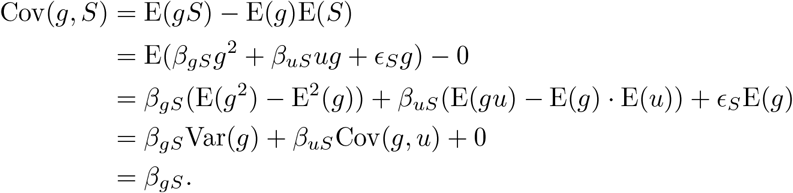

Similarly,

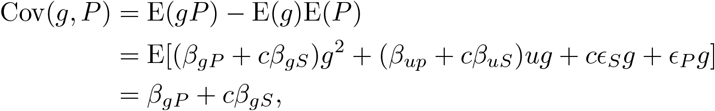

and

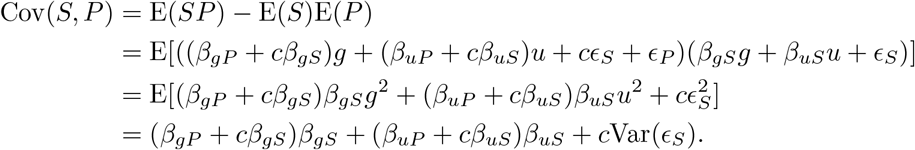

We can now rewrite the least square solution for the observed effect sizes as follows:

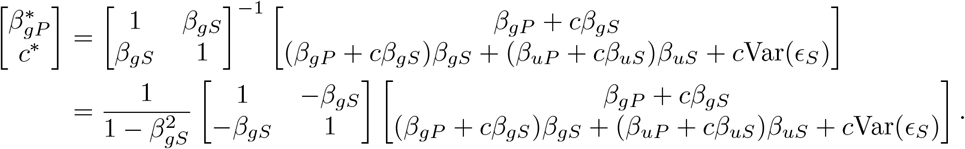

Notice that in the above we analytically computed the inverse of the susceptibility matrix that appeared in the least-squares formulation. Focusing on 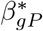, we get

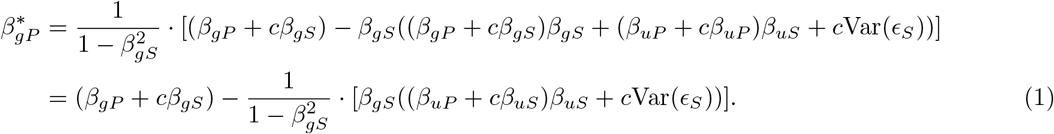

We can further compute the variance of *ϵ*_*i*_, i.e., Var(*ϵ*_*S*_), by looking at the variance of *S*, which is a sum of three independent components (recall that the variance of *S* is normalized and equal to one):

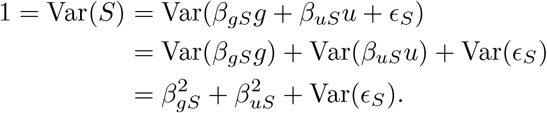

Therefore,

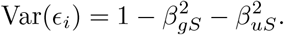

Combining with eqn. (1), we get

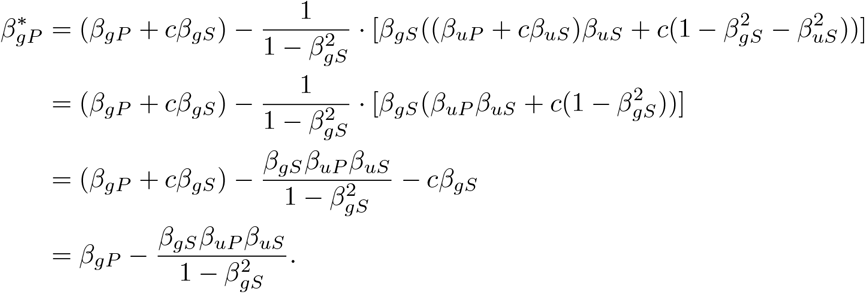

In order to further simplify and provide intuition for the above expression, recall that *u* is a composite variable accounting for all other causal factors shared by disease susceptibility and progression, including shared polygenic effects, with the exception of the variant of interest *g*. In order to proceed with our theoretical analysis, we will focus on the extreme case where *u*’s components only include the shared polygenic effect and there are no environmental risk factors. In other words, in our theoretical analysis, we care more about how the genetic relationship between disease susceptibility and progression affects downstream analysis.

Due to the polygenic nature of most complex diseases, a single variant usually contributes little to the total endpoint heritability. A reasonable way to model this assumption is to approximate *β*_*up*_ as follows:

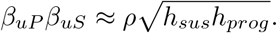

Using this approximation, it follows that

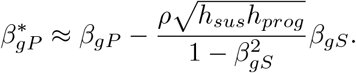

The above relationship indicates that when there is no shared non-genetic risk factors, GWAS results could suffer from index event bias depending on the genetic similarity between variants underlying disease susceptibility and the progression of the unique genetic components, and also the heritability of progression endpoint. Thus, with fixed susceptibility genetics, when the progression endpoint has really low heritability, the absolute biased effect size can also be relatively negligible.

### Simulation under proposed framework

We carried out a simulation based on the genome of chromosome 21 (containing 111,212 HM3 SNPs) for 10,000 synthetic European individuals created using Hapgen2 (Su et al., 2011). In the simulation, we fixed heritability of disease susceptibility at *h*_*sus*_ = 0.2, and impact of susceptibility liability on disease progression liability at *c* = 0.3. We further fixed the compositions of causal SNPs for each of these two endpoints so that 0.001 of total SNPs (ie. around 110 in this case) have direct effect on disease susceptibility and 0.001 have direct effect on progression. We changed the proportion of overlap between these two genetic components so that 25%, 50%, 75% of the causal SNPs were shared between susceptibility and progression. To decide effect size for each causal SNP *i*, we first drew a base effect independently from a standard univariate normal distribution

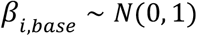

and multiply it to the square root of heritability this SNP accounts for to get its final effect size. Causal SNPs shared by susceptibility and progression were simulated to have same base effect on susceptibility and progression so that expected correlation of overall polygenic effects between two endpoints ρ will approximately correspond to the proportion of shared causal SNPs, in this case ρ = 0.25, 0.5 and 0.75. We further vary heritability of disease progression *h*_*prog*_ = 0.005, 0.1, and 0.2.

Under each simulation setup, we run standard GWAS correcting for top 10 PCs for both susceptibility and progression liability. Note that just like we added age of diagnosis as a covariate in our empirical mortality GWAS, in the progression GWAS, we also correct for susceptibility liability. Subsequently, we clump the GWAS results using plink (Purcell et al., 2007) under parameters --clump-p1 5e-8 --clump-r2 0.5 --clump-kb 250 to extract independent genome-wide significant loci from each GWAS, ran linemodels (Pirinen, 2023) and plot the results just like we did in our empirical experiments.

### Impact of index event bias and Slope-Hunter-like adjustment

To investigate the impact of index event bias, we think the most direct way would be to compare the underlying simulated SNP effects to observed effects from GWAS for disease progression. Recall that effect size for each causal SNP *i* is a standard normal variable multiplied by square root of its heritability, which can be expressed as

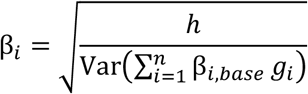

where *βi* is the underlying causal effect simulated, *h* is the endpoint heritability, *g*_*i*_ is the genotype of causal SNP *i*, and *n* is the total number of causal SNPs for the endpoint. The same equation applies to both susceptibility and progression causal genetic effects.

In this experiment as we are investigating the impact of index event bias, on top of shared polygenic effects, we introduced another component *u* to account for any other shared non-genetic risk factor between the two. Same as previous experiment, heritability of disease susceptibility and impact of susceptibility liability on disease progression liability were still fixed as *h*_*sus*_= 0.2 and c = 0.3. For this experiment, we further fixed the heritability of disease progression at *h*_*prog*_ = 0.005. We vary ρ = 0.25, 0.5 and 0.75, and contribution of the non-genetic component on variance of susceptibility and progression liability among *Var*_*u*_ = 0, 10%, or 20%.

Under each setup, we ran GWAS on disease susceptibility and progression as described before, and for all progression causal variants, we plotted simulated SNP effects against SNP effects observed from the progression GWAS. We examined the residual sum of squares (rss) for the points around function y = x. Furthermore, based on the theory behind of Slope-Hunter, we applied adjustment on SNPs that suffer from index event bias through a procedure described as below:

1. Extract all susceptibility specific causal SNPs and regress their observed effect sizes from susceptibility GWAS against progression GWAS to obtain the correction factor *b*.
2. For each causal SNP *i* shared between susceptibility and progression, compute the corrected progression genetic effect 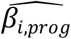 as below

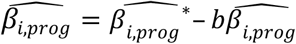

where 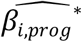 is the observed effect for SNP *i* from the conditioned progression GWAS, and 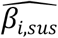 is the observed effect for SNP *i* from the susceptibility GWAS. Note that this experiment may demonstrate the utility of Slope-Hunter-like correction in a nearly “perfect” scenario, where the classification of SNPs (susceptibility specific, progression specific, shared or no effect in either) is given. In practice, a Bayesian or comparable approach needs to be applied for posterior variant group assignment, which can result in worse performance than shown in this manuscript. As a comparison, we show in the same plot the corrected variants effect sizes against the simulated underlying effects and examined rss. See **Figure** S19 for results. Note the previous experiment shows impact of index event bias and correction on all underlying causal variants for the progression, whereas in practice, such information is not a given. Therefore, subsequently we tried to examine the real impact of Slope-Hunter-like correction on observed results from GWAS under one of the conditions where the most severe index event bias could be observed (*h*_*prog*_ = 0.005, *Var*_*u*_ = 20%, ρ = 0.5). We chose ρ = 0.5 rather than ρ = 0.75, where more causal variants are shared, so that more susceptibility specific SNPs were available for correction factor (b) estimation and a more accurate estimate could be achieved. For this experiment, we first ran progression and susceptibility GWAS respectively as mentioned before and clumped their results to identify independent signals for each. Then using susceptibility specific SNPs, we fitted the correction factor and applied correction on all SNP effects in the progression GWAS. We also corrected for the standard errors as mentioned in (Mahmoud et al., 2022) and recomputed the p-values. Mindful of the difference from the previous experiment, that correction was not only applied on shared causal SNPs but all variants, as that would be what is done empirically. Note that here we still provided causal information for susceptibility specific variants to fit the correction factor, which was again a rather ideal use case for the method. Then, as what was done empirically, we ran linemodels (Pirinen, 2023) on SNP effect before and after the correction (**Figure** S20). None of the SNPs became genome-wide significant in progression GWAS after correction (**Table** S19), and there was no change in Bayesian classification of SNPs.

## Supporting information

All tables

## Data Availability

All data produced in the present study are available upon reasonable request to the authors

## Funding

This study has received funding from the European Union’s Horizon 2020 research and innovation programme under grant agreement No 101016775, from the European Research Council (ERC) under the European Union’s Horizon 2020 research and innovation program (grant number 945733) and from Academy of Finland fellowship grant N. 323116.

## Acknowledgements

We would like to thank Prof. George Davey Smith for the invaluable comments on this manuscript.

## FinnGen

We thank all those who contributed samples and data for the FinnGen scientific project; and P. VandeHaar for technical consultation on PheWeb. The FinnGen project is funded by two grants from Business Finland (HUS 4685/31/2016 and UH 4386/31/2016) and the following industry partners: AbbVie, AstraZeneca UK, Biogen, Bristol Myers Squibb (and Celgene Corporation & Celgene International II), Genentech, Merck Sharp & Dohme LLC, a subsidiary of Merck & Co., Inc., Rahway, NJ, USA, Pfizer, GlaxoSmithKline Intellectual Property Development, Sanofi US Services, Maze Therapeutics, Janssen Biotech, Novartis, and Boehringer Ingelheim. The following biobanks are acknowledged for delivering samples to FinnGen: Auria Biobank (https://www.auria.fi/biopankki/), THL Biobank (https://www.thl.fi/biobank), Helsinki Biobank (https://www.helsinginbiopankki.fi), Biobank Borealis of Northern Finland (https://www.ppshp.fi/Tutkimus-ja-opetus/Biopankki/Pages/Biobank-Borealis-briefly-in-English.aspx), Finnish Clinical Biobank Tampere (https://www.tays.fi/en-US/Research_and_development/Finnish_Clinical_Biobank_Tampere), Biobank of Eastern Finland (https://www.ita-suomenbiopankki.fi/en), Central Finland Biobank (https://www.ksshp.fi/fi-FI/Potilaalle/Biopankki), Finnish Red Cross Blood Service Biobank (www.veripalvelu.fi/verenluovutus/biopankkitoiminta) and Terveystalo Biobank (https://www.terveystalo.com/fi/Yritystietoa/Terveystalo-Biopankki/Biopankki/). All Finnish biobanks are members of the BBMRI.fi infrastructure (https://www.bbmri.fi). The FINBB (https://finbb.fi/) is the coordinator of BBMRI-ERIC operations in Finland. The Finnish biobank data can be accessed through the Fingenious services (https://site.fingenious.fi/en/) managed by FINBB.

## UK biobank

Ethics approval for the UK Biobank study was obtained from the North West Centre for Research Ethics Committee (11/NW/0382). UK Biobank data used in this study were obtained under approved application 78537.

## Estonia biobank

The authors would also like to acknowledge all the recruiters and the participants of the EGCUT, the University of Tartu, the Ministry of Social Affairs, the Ministry of Science and Education, the Ministry of Economic Affairs and Communications, the Archimedes Foundation, the Estonian Biocentre, the Institute of Molecular and Cell Biology and the Centre for Ethics of the University of Tartu. The authors would also like to acknowledge the EGCUT technical personnel.

Other contributors: Annely Allik, Tarmo Annilo, Merli Hass, Atso-Heinar Jõks, Aidula-Taie Kaasik, Aime Keis, Erkki Leego, Merike Leego, Kadri Lilienthal, Kristjan Metsalu, Evelin Mihailov, Kairit Mikkel, Ene Mölder, Helja Niinemäe, Tiit Nikopensius, Mairo Puusepp, Steven Smit, Viljo Soo, Riin Tamm, Maris Teder-Laving andMaris Väli-Täht.

## Genomics England

This research was made possible through access to data in the National Genomic Research Library, which is managed by Genomics England Limited (a wholly owned company of the Department of Health and Social Care). The National Genomic Research Library holds data provided by patients and collected by the NHS as part of their care and data collected as part of their participation in research. The National Genomic Research Library is funded by the National Institute for Health Research and NHS England. The Wellcome Trust, Cancer Research UK and the Medical Research Council have also funded research infrastructure.We acknowledge the contribution of the Genomics England Research Consortium. The members of this consortium are: John C. Ambrose1, Prabhu Arumugam1, Roel Bevers1, Marta Bleda1, Freya Boardman-Pretty1,2, Christopher R. Boustred1, Helen Brittain1, Matt J. Brown1, Mark J. Caulfield1,2, Georgia C. Chan1, Adam Giess1, Angela Hamblin1, Shirley Henderson1,2, Tim J. P. Hubbard1, Rob Jackson1, Louise J. Jones1,2, Dalia Kasperaviciute1,2, Melis Kayikci1, Athanasios Kousathanas1, Lea Lahnstein1, Sarah E.A. Leigh1, Ivonne U. S. Leong1, Javier F. Lopez1, Fiona Maleady-Crowe1, Meriel McEntagart1, Federico Minneci1, Jonathan Mitchell1, Loukas Moutsianas1,2, Michael Mueller1,2, Nirupa Murugaesu1, Anna C. Need1,2, Peter O’Donovan1, Chris A.Odhams1, Christine Patch1,2, Daniel Perez-Gil1, Mariana Buongermino Pereira1, John Pullinger1, Tahrima Rahim1, Augusto Rendon1, Tim Rogers1, Kevin Savage1, Kushmita Sawant1, Richard H.Scott1, Afshan Siddiq1, Alexander Sieghart1, Samuel C. Smith1, Alona Sosinsky1,2, Alexander Stuckey1, Mélanie Tanguy1, Ana Lisa Taylor Tavares1, Ellen R. A. Thomas1,2, Simon R.Thompson1, Arianna Tucci1,2, Matthew J. Welland1, Eleanor Williams1, Katarzyna Witkowska1,2, Suzanne M. Wood1,2, Magdalena Zarowiecki1.

1. Genomics England, London, UK
2. William Harvey Research Institute, Queen Mary University of London, London, EC1M 6BQ, UK

## Generation Scotland

Generation Scotland received core support from the Chief Scientist Office of the Scottish Government Health Directorates [CZD/16/6] and the Scottish Funding Council [HR03006]. Genotyping of the GS:SFHS samples was carried out by the Genetics Core Laboratory at the Wellcome Trust Clinical Research Facility, Edinburgh, Scotland and was funded by the Medical Research Council UK and the Wellcome Trust (Wellcome Trust Strategic Award “STratifying Resilience and Depression Longitudinally” (STRADL) Reference 104036/Z/14/Z).

We are grateful to all the families who took part, the general practitioners and the Scottish School of Primary Care for their help in recruiting them, and the whole Generation Scotland team, which includes interviewers, computer and laboratory technicians, clerical workers, research scientists, volunteers, managers, receptionists, healthcare assistants and nurses.

Ethical approval for the GS:SFHS study was obtained from the Tayside Committee on Medical Research Ethics (on behalf of the National Health Service).

Ethical approval for the GS:3D study was obtained from the Tayside Committee on Medical Research Ethics (on behalf of the National Health Service).

Ethical approval for the GS:21CGH study was obtained from the Scotland A Research Ethics Committee.

## Genes & Health

Genes & Health is/has recently been core-funded by Wellcome (WT102627, WT210561), the Medical Research Council (UK) (M009017, MR/X009777/1, MR/X009920/1), Higher Education Funding Council for England Catalyst, Barts Charity (845/1796), Health Data Research UK (for London substantive site), and research delivery support from the NHS National Institute for Health Research Clinical Research Network (North Thames). Genes & Health is/has recently been funded by Alnylam Pharmaceuticals, Genomics PLC; and a Life Sciences Industry Consortium of Astra Zeneca PLC, Bristol-Myers Squibb Company, GlaxoSmithKline Research and Development Limited, Maze Therapeutics Inc, Merck Sharp & Dohme LLC, Novo Nordisk A/S, Pfizer Inc, Takeda Development Centre Americas Inc.

We thank Social Action for Health, Centre of The Cell, members of our Community Advisory Group, and staff who have recruited and collected data from volunteers. We thank the NIHR National Biosample Centre (UK Biocentre), the Social Genetic & Developmental Psychiatry Centre (King’s College London), Wellcome Sanger Institute, and Broad Institute for sample processing, genotyping, sequencing and variant annotation.

We thank: Barts Health NHS Trust, NHS Clinical Commissioning Groups (City and Hackney, Waltham Forest, Tower Hamlets, Newham, Redbridge, Havering, Barking and Dagenham), East London NHS Foundation Trust, Bradford Teaching Hospitals NHS Foundation Trust, Public Health England (especially David Wyllie), Discovery Data Service/Endeavour Health Charitable Trust (especially David Stables), Voror Health Technologies Ltd (especially Sophie Don), NHS England (for what was NHS Digital) - for GDPR-compliant data sharing backed by individual written informed consent.

## BioMe Biobank

This work was supported in part through the computational and data resources and staff expertise provided by Scientific Computing and Data at the Icahn School of Medicine at Mount Sinai and supported by the Clinical and Translational Science Awards (CTSA) grant UL1TR004419 from the National Center for Advancing Translational Sciences. Additionally, this work was supported by the Office of Research Infrastructure of the National Institutes of Health under award number S10OD026880, which allowed us to use Mount Sinai Data Warehouse (MSDW) data. Regarding HPI.MS resources, funding was provided by the Hasso Plattner Foundation (HPF). The Mount Sinai BioMe Biobank has been supported by The Andrea and Charles Bronfman Philanthropies and in part by Federal funds from the NHLBI and NHGRI (U01HG00638001; U01HG007417; X01HL134588). We thank all participants in the Mount Sinai BioMe Biobank. We also thank all of our recruiters who have assisted in data collection and management and are grateful for the computational resources and staff expertise provided by Scientific Computing at the Icahn School of Medicine at Mount Sinai.

## Supplementary Method

### Data and resources

#### FinnGen

##### Genotyping and quality control

FinnGen consists of prospectively recruited samples and a series of legacy cohorts with genotypes already available (*FinnGen Project* | *FinnGen*, n.d.). Prospective samples were genotyped using the ThermoFisher Axiom custom array which tags a total of 655,973 variants. Genotype calling was performed using the Array Power Tools software. Legacy cohorts were genotyped using various Illumina arrays and genotype calling was performed using either GenCall or zCall algorithms.

For both prospective and legacy cohorts the following quality control metrics were used. Samples were removed if:

- Pihat was > 0.9 and the samples were not monozygotic or replicates
- There was a discrepancy between reported sex and genetically determined sex (F-value ≤ 0.3 for females and ≥ 0.8 for males)
- Missingness was ≥ 5%
- Heterozygosity was ±4 standard deviations from the population average
- Pihat was > 0.1 with 14 or more samples
- Samples were ±4 standard deviations away from the population average according to the first two genetic principal components.

Samples were tagged should there be evidence of a mendelian error or contain replicate samples with over 50,000 discrepancies.

Variants were removed if:

- The variant failed the Hardy-Weinberg Equilibrium test (p-value < 10-6)
- The variant had a call rate < 98%

##### Imputation

Pre-phasing was performed using Eagle 2.3.5 (Loh et al., 2016) and samples were imputed using the SiSu v3 imputation reference panel. This reference panel is specific to the Finnish population, containing high-coverage (25-30x) whole-genome sequencing data from 3,775 Finns and 16,962,023 variants with minor allele count ≥ 3. After imputation, 16,387,711 variants were imputed with high quality (INFO > 0.6).

##### Ancestry assignment

Firstly, the FinnGen samples were combined with the 1000 genomes phase 3 dataset. Genetic principal components were calculated using a subset of 49,451 pruned SNPs. Aberrant (Bellenguez et al., 2012) was used to identify and remove samples that deviated from the main cluster. A probability of belonging to either a North-Western European or Finnish population was calculated by firstly performing PCA with individuals belonging to these ancestries from 1000 genomes data. FinnGen samples were then projected onto this PCA space and Mahalanobis distances calculated for each sample against each of the two ancestries. Samples were retained if there was ≥ 95% probability of belonging to the Finnish ancestry cluster.

##### Ethics statement

Patients and control subjects in FinnGen provided informed consent for biobank research, based on the Finnish Biobank Act. Alternatively, separate research cohorts, collected prior the Finnish Biobank Act came into effect (in September 2013) and start of FinnGen (August 2017), were collected based on study-specific consents and later transferred to the Finnish biobanks after approval by Fimea (Finnish Medicines Agency), the National Supervisory Authority for Welfare and Health. Recruitment protocols followed the biobank protocols approved by Fimea. The Coordinating Ethics Committee of the Hospital District of Helsinki and Uusimaa (HUS) statement number for the FinnGen study is Nr HUS/990/2017.

The FinnGen study is approved by Finnish Institute for Health and Welfare (permit numbers: THL/2031/6.02.00/2017, THL/1101/5.05.00/2017, THL/341/6.02.00/2018, THL/2222/6.02.00/2018,

THL/283/6.02.00/2019, THL/1721/5.05.00/2019 and THL/1524/5.05.00/2020), Digital and population data service agency (permit numbers: VRK43431/2017-3, VRK/6909/2018-3, VRK/4415/2019-3), the Social Insurance Institution (permit numbers: KELA 58/522/2017, KELA 131/522/2018, KELA 70/522/2019, KELA 98/522/2019, KELA 134/522/2019, KELA 138/522/2019, KELA 2/522/2020, KELA16/522/2020), Findata permit numbers THL/2364/14.02/2020, THL/4055/14.06.00/2020, THL/3433/14.06.00/2020, THL/4432/14.06/2020, THL/5189/14.06/2020, THL/5894/14.06.00/2020, THL/6619/14.06.00/2020, THL/209/14.06.00/2021, THL/688/14.06.00/2021, THL/1284/14.06.00/2021, THL/1965/14.06.00/2021,THL/5546/14.02.00/2020, THL/2658/14.06.00/2021, THL/4235/14.06.00/2021,Statistics Finland (permit numbers: TK-53-1041-17 and TK/143/07.03.00/2020 (earlier TK-53-90-20) TK/1735/07.03.00/2021, TK/3112/07.03.00/2021) and Finnish Registry for Kidney Diseases permission/extract from the meeting minutes on 4th July 2019.

The Biobank Access Decisions for FinnGen samples and data utilized in FinnGen Data Freeze 10 include: THL Biobank BB2017_55, BB2017_111, BB2018_19, BB_2018_34, BB_2018_67, BB2018_71, BB2019_7, BB2019_8, BB2019_26, BB2020_1, BB2021_65, Finnish Red Cross Blood Service Biobank 7.12.2017, Helsinki Biobank HUS/359/2017, HUS/248/2020, HUS/150/2022 § 12, §13, §14, §15, §16, §17, §18, and §23, Auria Biobank AB17-5154 and amendment #1 (August 17 2020) and amendments BB_2021-0140, BB_2021-0156 (August 26 2021, Feb 2 2022), BB_2021-0169, BB_2021-0179, BB_2021-0161, AB20-5926 and amendment #1 (April 23 2020)and it’s modification (Sep 22 2021), Biobank Borealis of Northern Finland_2017_1013, 2021_5010, 2021_5018, 2021_5015, 2021_5023, 2021_5017, 2022_6001, Biobank of Eastern Finland 1186/2018 and amendment 22 § /2020, 53§/2021, 13§/2022, 14§/2022, 15§/2022, Finnish Clinical Biobank Tampere MH0004 and amendments (21.02.2020 & 06.10.2020),§8/2021, §9/2022, §10/2022, §12/2022, §20/2022, §21/2022, §22/2022, §23/2022, Central Finland Biobank 1-2017, and Terveystalo Biobank STB 2018001 and amendment 25th Aug 2020, Finnish Hematological Registry and Clinical Biobank decision 18th June 2021, Arctic biobank P0844: ARC_2021_1001.

#### UK Biobank

##### Genotyping and quality control

UK Biobank participants were genotyped by two genotyping arrays: The UK Biobank Lung Exome Variant Evaluation (UKBiLEVE) Axiom array was used to genotype 49,950 participants. The remaining 438,427 participants were genotypes using the Applied Biosystems UK Biobank Axiom Array. Principal Component Analysis (PCA) was performed on the genetic data and centralised quality control (QC) on variants was performed on individuals identified to belong to the largest cluster (N=463,844) according to Aberrant - an unsupervised clustering algorithm (Bellenguez et al., 2012). Variants were assessed for evidence of allele frequency variation across batch, plate, sex or array and that genotypes were largely consistent with Hardy-Weinberg Equilibrium expectations (all p-value thresholds < 10^−12^). If a variant failed one or more tests within a given batch it was set to missing. See (UK Biobank, 2015) for more detailed information on testing.

##### Imputation

For 487,442 individuals, imputation was performed using the IMPUTE4 (Howie et al., 2009) software. Genetic variation from the Haplotype Reference Consortium (HRC) (McCarthy et al., 2016) and merged UK10K+1000 Genomes (1000 Genomes Project Consortium, 2015) were used as a reference panel. Single Nucleotide Polymorphisms (SNPs) were only included in the final imputation if they were present in both reference panels, giving a total of 96,959,328 SNPs.

##### Ancestry assignment

Ancestry assignment uses methodology and scripts from GenoPred (*Prediction within Ancestral Diversity*, n.d.). Individuals were stratified into one of five super populations African (AFR), American (AMR), South Asian (SAS), East Asian (EAS) and European (EUR). The 1000 Genomes data (1000 Genomes Project Consortium, 2015) acted as a reference given the individuals are known to belong to one of the 5 super populations. Only unambiguous SNPs also present in both the HapMap3 consortium (Gibbs et al., 2003) and the imputed UK Biobank data were retained for PCA. SNPs within both the reference (1000 Genomes) and target (UK Biobank) samples underwent quality control such that the minor allele frequency (MAF) > 5%, variant missingness > 2% and Hardy-Weinberg Equilibrium p-value > 1e^-6^.467,970 autosomal SNPs remained following QC and were in the intersection of the reference and target samples. Regions with long range linkage disequilibrium were excluded and independent SNPs (SNPs greater than 1000kb apart and r^2^ < 0.2) retained. PCA was then performed in the reference sample using PLINK v2 (Purcell et al., 2007) and a multinomial elastic-net regression was trained using 5-fold cross validation, super population as the outcome and the first 10 PCs as covariates. PCs from the target sample were then projected into the reference space and prediction on super population made. Classifications were made according to the super population with the greatest probability. To be classified the max probability must be over 0.5, otherwise it was set to missing.

PCA was performed using a random subset of 1000 individuals per super population and PC’s from the rest of the super population sample projected onto this space. Distances from the centroid were calculated and outliers removed. Outliers were classified as having a distance > 75 percentile + 30*Interquartile Range. Following within-ancestry QC, 8,381, 1,063, 2,393, 447,332 and 9,435 individuals were allocated to AFR, AMR, EAS, EUR and SAS super populations respectively.

#### Estonian Biobank

##### Genotyping and quality control

Estonian BioBank (EstBB) samples were genotyped with 4 sub-versions of Infinium Global Screening Array-24. Samples with less than 95% call-rate were excluded. Sample sex recorded in the EstBB database was compared with genetic sex. Samples with sex mismatch were further inspected for sex chromosome abnormalities (X0, XXY, etc.), and samples with confirmed database vs genetic sex mismatch were excluded. In total, 202 910 individuals passed sample quality control. SNP quality control was performed by excluding: (a) all SNPs with less than 95% call-rate, (b) SNPs showing more than 5% AF difference from the AF mean estimated using all genotyping batches with more than 10 000 samples per batch, (c) SNPs with Illumina GenTrain score < 0.6 or cluster separation score < 0.4 in any genotyping batch, (d) autosomal SNPs with HWE exact test p-value < 1e-4. In total, approximately 328K autosomal and X-chromosome SNPs with MAF > 1% passed quality control and were used in the imputation. All the variants were processed on the human genome assembly GRCh37.

##### Imputation

Imputation was performed using a local Estonian imputation reference panel made of 2056 WGS samples. Genotypes were pre-phased with Eagle v2.4.1 and imputed with Beagle 5.1 using default parameters. Multiallelic positions were excluded from imputation output. In total, 39 546 641 variants were used in the study.

##### Ancestry assignment

EstBB samples were combined with the 1000 genomes phase 3 dataset for ancestry analysis. Genetic principal components were calculated using a subset of quality controlled and pruned genotyped SNPs. This was further used to identify and remove samples that deviated from the main cluster via visual inspection. In total, 481 non-european ancestry individuals based on principal components were excluded from the analysis.

#### Genomics England

##### Genotyping and quality control

Genome sequencing was performed in DNA samples from 78,195 individuals using Illumina HiSeq X systems (150bp paired-end format). Reads were aligned using the iSAAC Aligner (version 03.16.02.19) and small variants were called using Starling Small Variant Caller (version 2.4.7). Samples were aligned to the Homo Sapiens NCBI GRCh38 assembly with decoys.

Aggregation of single-sample gVCFs was performed using the Illumina software gVCF genotyper (version 2019). Variant normalisation and decomposition were implemented by vt (version 0.57721). Genomic annotation and calculation of allele statistics were performed using Ensembl VEP and bcftools respectively. The multi-sample VCF dataset (aggV2) was then split into 1,371 roughly equal chunks to allow faster processing. Only variants that passed all provided site quality control criteria were processed.

##### Imputation

The WGS genotypes (∼722M variants) were filtered to a variant base list used for PGS model generation, which includes 18,421,839 variants. (For further information on how the variant list was derived see: https://research-help.genomicsengland.co.uk/pages/viewpage.action?pageId=72351761)

Genotypes were phased and imputed using the 1000G reference panel (v5a) which was lifted-over from GRCh37 to GRCh38 using cross-map.

##### Ancestry assignment

The genetic ancestry of the patients was estimated using a random forest classifier and data from 1000 genomes project phase 3 (1KGP3) dataset. Firstly, all unrelated samples from the 1KGP3 were selected and 188,382 HQ SNPs were subsetted. After filtering for MAF > 0.05 in 1KGP3 (and GE data), the first 20 PCs were calculated using GCTA and the aggV2 data were projected onto the 1KGP3 PC loadings. The random forest model to predict ancestries was trained based on:

A. First 8 1KGP3 PCs
B. set Ntrees = 400
C. Train and predict on 1KGP3 Admixed American, African, East Asian, European, and South Asian super-populations.

Individuals were assigned for any one ancestry with a probability of > 0.8.

#### Genes and Health

##### Genotyping and quality control

We used the latest 2021 July GNH data release including 44,190 individuals (26,537 British-Bangladeshi, 17,653 British-Pakistani). Genotyping was performed on DNA samples from saliva, using the Illumina Infinium Global Screening Array v3, which contained 730,059 variants. GenomeStudio from Illumina was used to perform clustering and initial quality control on the genotype data. Variants were removed if they had low call rate, or were tagging structural variants, a positive HetExcess > 0.03, Hardy-Weinberg equilibrium P-value < 1.0 × 10^−6^, cluster sep <0.57, or automated clustering (GenTrain) score <= 0.7. A total of 637,829 variants remained with call rates of > 0.992 for female samples and > 0.995 for male samples (including X and Y chromosomes). Sample exclusion criteria included duplicate GSA genotypes that should not be sample duplicates, samples that should be duplicated but have not matching GSA genotypes, and a few late withdrawals of consent. Only chip genotyped samples with valid NHS numbers were preserved. When two chip genotype samples with the same NHS number were found, the samples with the highest call rate were retained.

##### Imputation

Monomorphic SNPs, non-ACGT, palindromic (A/T, T/A, C/G, G/C), and chr Y variants were excluded. Variants were evaluated by TOPMed QC to obtain SNPs that required strand flipping (performed in plink). Furthermore, variants with MAF<0.0001 were excluded. The TOPMed-r2 Minimac4 Imputation Server (version 1.5.7, https://imputation.biodatacatalyst.nhlbi.nih.gov/#!pages/home), created by the University of Michigan, was subsequently used to impute the genotypes. Rsq filter (imputation quality) of 0.3 was applied within the Imputation Server.

##### Ancestry assignment

A total of 44,396 individuals and 355,862 directly genotyped variants (retaining only autosomal variants, MAF>0.01, call rate >99% and those passing HWE in declared Bangladeshi individuals) were used with the KING software to estimate pairwise relationship up to 4 degrees. PCA was performed on GNH unrelated individuals, projecting related individuals into the PC, to obtain 50 PCs for all GNH samples. For the ancestry assignment, we used a reference cohort consisting of 3,433 individuals from 1000G and HGDP. A PCA up to 50 PCs was performed on the reference set (3,433 individuals and 104,552 variants) and subsequently the GNH samples were projected into the reference PCA. Using UMAP with 7 PCs, we genetically inferred Bangladeshi and Pakistani individuals and excluded 76 non South Asian outliers and 130 South Asian outliers (not falling into the main clusters).

#### Generation Scotland

##### Genotyping and quality control

Generation Scotland (GS) consists of ∼24,000 individuals from across Scotland aged between 18-99 years. Phenotypic data were obtained at baseline along with whole blood samples for DNA quantification. Disease outcomes were ascertained through linkage to primary (GP) and secondary (hospital) healthcare records. Genotype data was assayed for 20,195 participants in two batches with 9,863 participants in the first batch and the remainder in the second. The genotyping was performed using the Illumina HumanOmniExpressExome-8 v1.0 BeadChip and the Illumina HumanOmniExpressExome-8 v1.2 BeadChip, respectively. Individuals or SNPs with a low call rate (<98%) and SNPs with Hardy-Weinberg p-value<1x10^−6^ were removed. Mendelian errors were removed by setting the individual-level genotypes at erroneous SNPs to missing.

##### Imputation

Genotyped data were imputed using the HRC panel v1.1 (McCarthy et al., 2016). Autosomal haplotypes were checked to ensure consistency with the reference panel (strand orientation, reference allele, position. Pre-phasing was performed using Shapeit2 v2r837 (O’Connell et al., 2014) using the Shapeit2 duohmm option11 (O’Connell et al., 2014) and cohort family structure in order to improve imputation quality (O’Connell et al., 2014). Variants with low imputation quality (INFO<0.4) as well as monogenic variants were removed from the imputed set resulting in 24,111,857 variants for downstream analysis.

##### Ancestry assignment

Ancestry outliers were removed from the dataset. These were defined as individuals who were more than six standard deviations away from the mean in a principal component analysis of GS merged with 1092 participants from the 1000 Genomes Project (1000 Genomes Project Consortium, 2015).

#### Dana Farber

##### Genotyping and quality control

DNA samples were processed from the whole blood and genotyped on either the Illumina Multi-Ethnic Genotyping Array (MEGA), the Expanded Multi-Ethnic Genotyping Array (MEGA Ex) array, or the Multi-Ethnic Global (MEG) BeadChip (Bien et al., 2016). All germline samples were imputed to the Haplotype Reference Consortium (HRC) reference panel (McCarthy et al., 2016) and then restricted to ∼ 1.1 million HapMap3 variants that typically exhibit high imputation accuracy across genotyping platforms and uniformly tag common SNP variation (Finucane et al., 2015). Small indels were not available in the HRC reference panel due to sequencing ambiguity, and we additionally imputed small indels into the germline genotyped data using the 1000 Genomes Phase 3 reference panel (1000 Genomes Project Consortium, 2015) and restricted to high-quality indels with INFO score (imputation confidence score) > 0.9.

##### Imputation

We assessed three imputation algorithms intended for low-coverage data: STITCH v1.5.3 (Davies et al., 2016), GLIMPSE v1.0.0 (Davies et al., 2021; Rubinacci et al., 2021), and QUILT v0.1.9 (Davies et al., 2021). For all analyses, OncoPanel data was aligned to hg19 using bwa and processed with the GATK IndelRealigner. The 1000 Genomes Phase 3 release was used as a haplotype reference, targeting variants with > 1% frequency in the European population. Tumor imputation was performed using the 1000 Genomes reference (rather than the HRC reference) because the HRC panel is not publicly available and the HRC imputation server does not support raw sequencing data. We thus sought to use the best reference panels that were accessible for the two data types. We note that HRC largely improves imputation accuracy for low-frequency variants (McCarthy et al., 2016), which were not the target of our analysis.

Imputation with STITCH was carried out on all samples using aligned reads in 5-MB batches (see the “Availability of data and materials” section for the detailed parameters and code). The potential influence of target cohort size was evaluated by randomly downsampling to a lower number of sequenced tumors. Imputation with QUILT was carried out using the same input and batching procedure, with default parameters. Imputation with GLIMPSE was carried out on all samples with default parameters as recommended in the documentation: calling genotype likelihoods from each raw BAM file, splitting the genome into chunks, performing imputation and phasing, and ligating the chunks. An alternative, reference-only version of GLIMPSE was kindly provided to us by the authors but could not be compiled in our computing environment. Lastly, we considered two other imputation approaches: GeneImp (Spiliopoulou et al., 2017) and BEAGLE (Browning et al., 2021), but found that their computational requirements were infeasible for sample sizes in the thousands. Identical reference panel data was used for all methods except small indels, structural variants, and multi-allelic polymorphisms were excluded from the STITCH and GLIMPSE analysis (which only allows biallelic single nucleotides). After imputation, variants were considered “filtered” if they had a minor allele frequency > 1% and an INFO score (imputation confidence score) > 0.4 (similar to parameters used previously (S. Liu et al., 2018)).

##### Ancestry assignment

Samples were projected into genetic ancestry principal components using the weights previously derived by the SNPWEIGHTS software (Chen et al., 2013) for the continental populations. Weights were constructed from the 1000 Genomes reference groups with ancestry from Northern/Western Europe (CEU), Western Africa (YRI), and China (CHB+CHD). In our data, each component was projected independently as a linear combination of the weights and individual sample dosages (using the plink2 “--score” command). Components were then linearly recalibrated by fitting to self-reported race as an outcome (note this linear recalibration is for interpretation purposes only and does not influence the significance of any downstream associations). To estimate ancestry fractions, we uniformly rescaled the African and Asian components to be between 0 and 1 and additionally uniformly scaled the ancestry of each individual to be between 0 and 1.

#### BioMe

##### Genotyping and quality control

BioMe participants have been genotyped using Illumina’s Global Screening Array (GSA-24 v1). Samples flagged as being contaminated, possibly duplicated, having low coverage, a call rate < 95%, or showing genotype-exome discordance were removed. Sex discordant samples were etiher reconciled after a plate swap resolution or removed. Sample missingness and depth of coverage were calculated using vcftools: mean missingness was 1.24 x 10^−3^, mean depth of coverage for all samples was 36.4x. Variant missingness and depth of coverage were calculated using vcftools (Danecek et al., 2011): mean missingness rate of 1.24 x 10^−3^, mean depth of coverage for all coding sites was 36.4x. Sites with HWE P-values < 1e-6 were retained but flagged.

##### Imputation

Imputation was performed using the 1000G (1000 Genomes Project Consortium, 2015) and TOPMed (Taliun et al., 2021) reference panel, and the software packages Beagle (Browning et al., 2021) and Impute2 (Howie et al., 2009). A filter of r2 > 0.7 was applied. Approximately 31,700 samples and 7,8M variants passed QC and were used in downstream analyses.

##### Ancestry assignment

We inferred the genetic ancestry following the guidelines of the Pan UKBB (*Quality Control (QC)* | *Pan UKBB*, n.d.). We performed a PCA using PLINK (Purcell et al., 2007), excluding relatives above 2nd-degree (kinship method, estimated using KING (Manichaikul et al., 2010)) and variants with MAF < 0.05. We trained a random forest classifier to infer the cohort’s genetic ancestry using the 1000G labels as reference, removed outliers (by only including the quantiles 0.25-0.90) and participants with mixed ancestry (random forest probability ≤ 0.5). Inferred ancestry: AMR (n=5,336), AFR (n=5,660), EUR (n=7,447), SAS (n=613), and EAS (n=728).

## Supplementary figures

**Figure S1.**
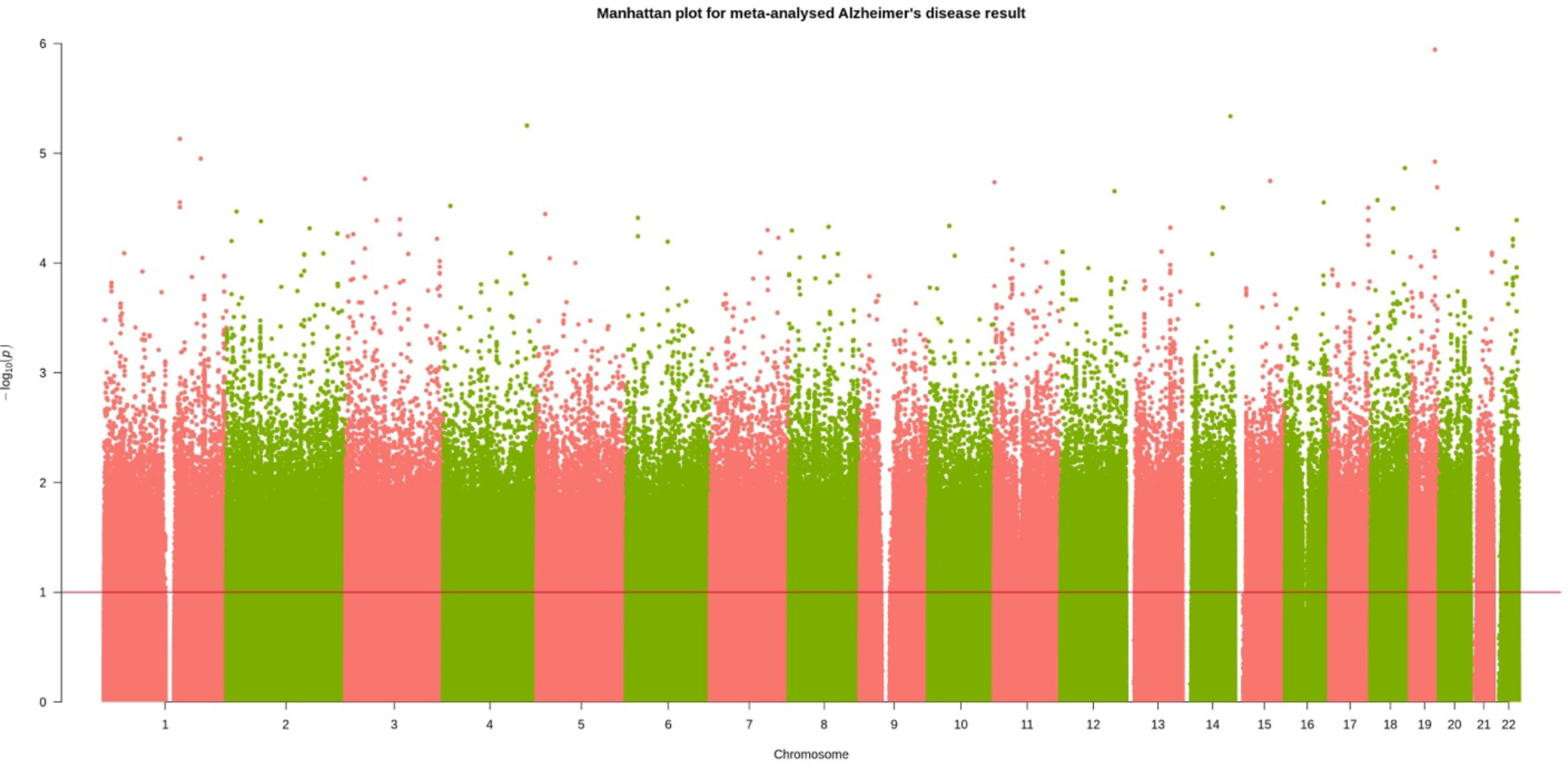
Manhattan plot for meta-analysed Alzheimer’s disease mortality GWAS.

**Figure S2.**
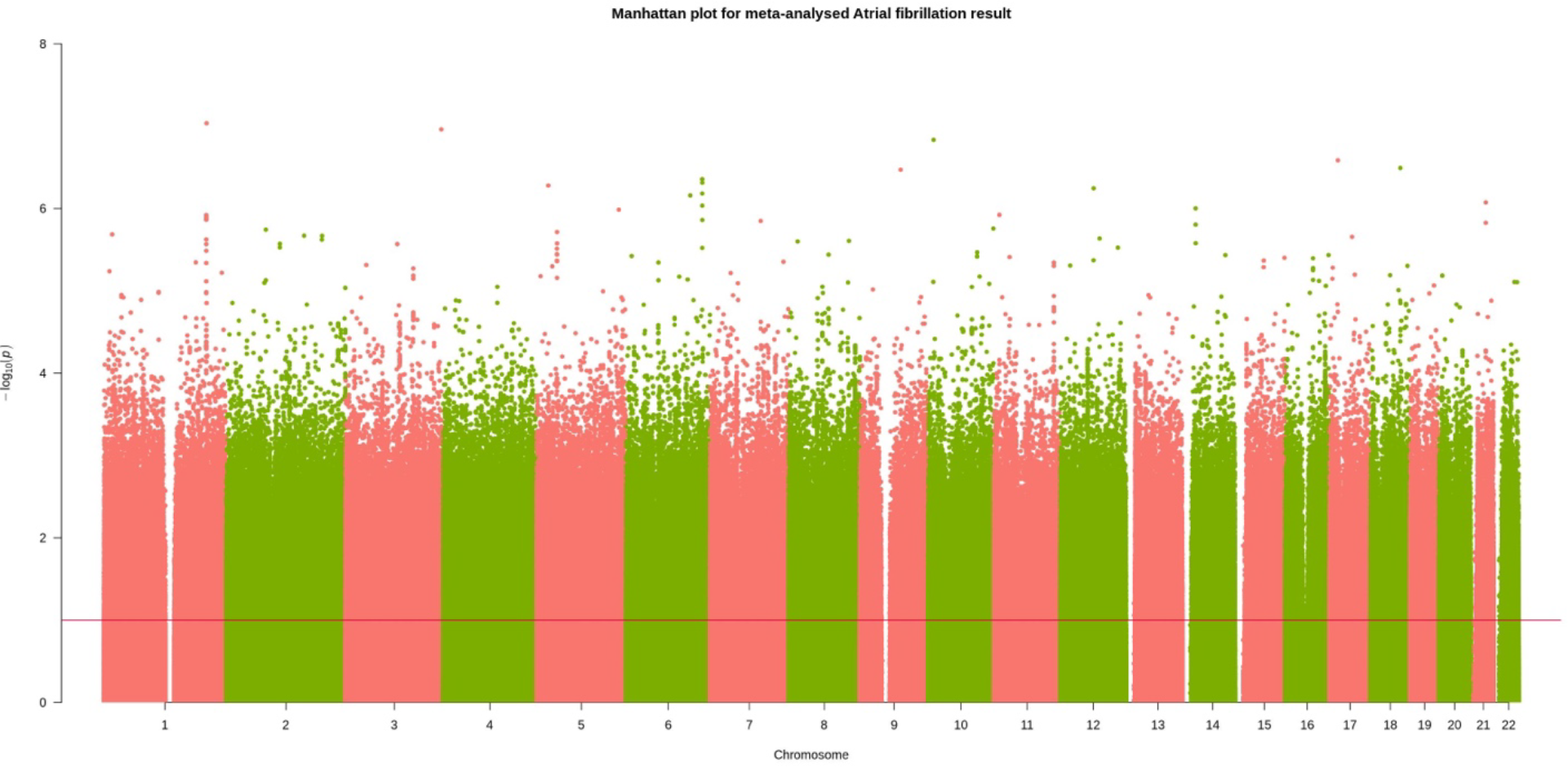
Manhattan plot for meta-analysed atrial fibrillation mortality GWAS.

**Figure S3.**
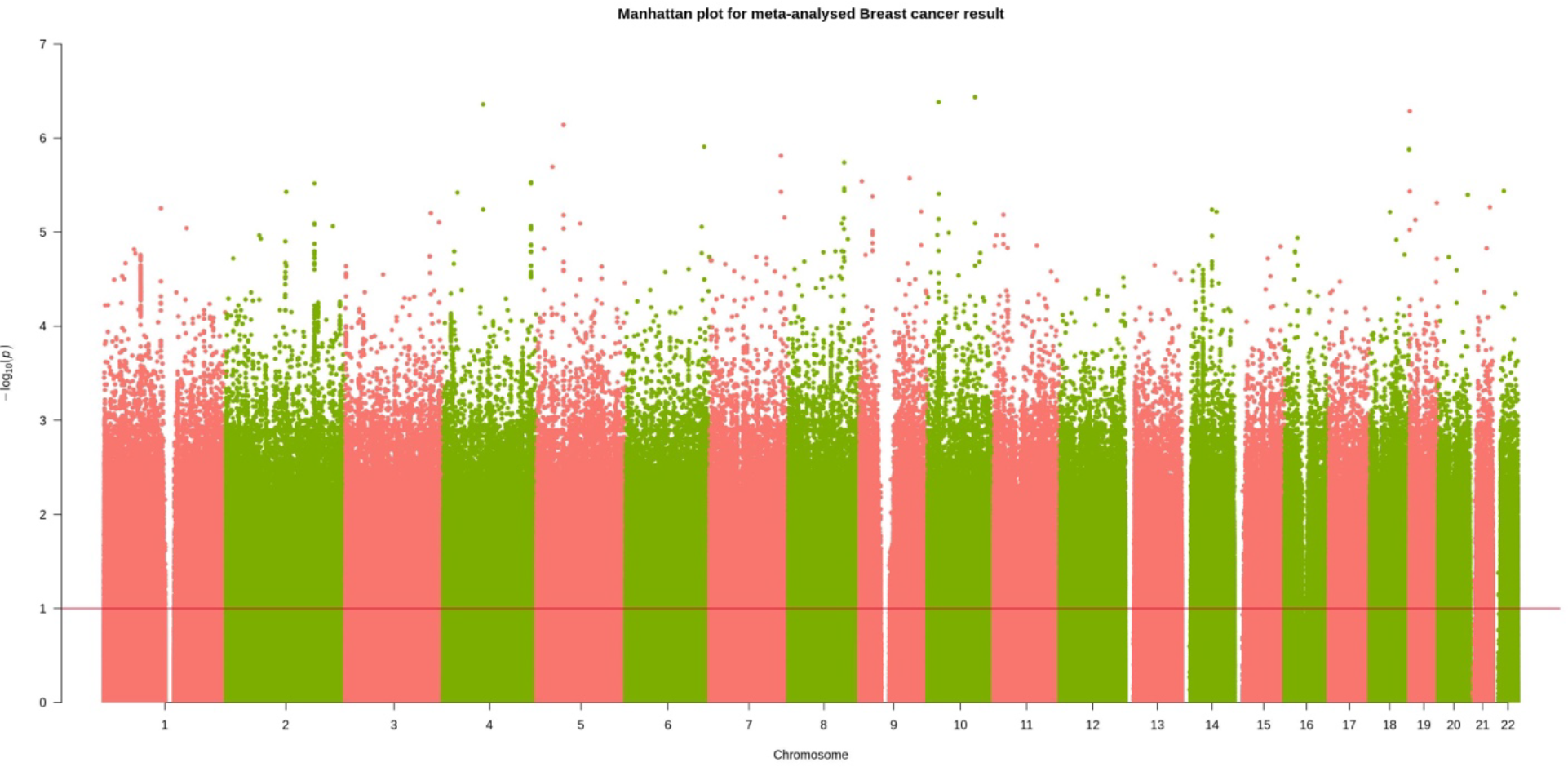
Manhattan plot for meta-analysed breast cancer mortality GWAS.

**Figure S4.**
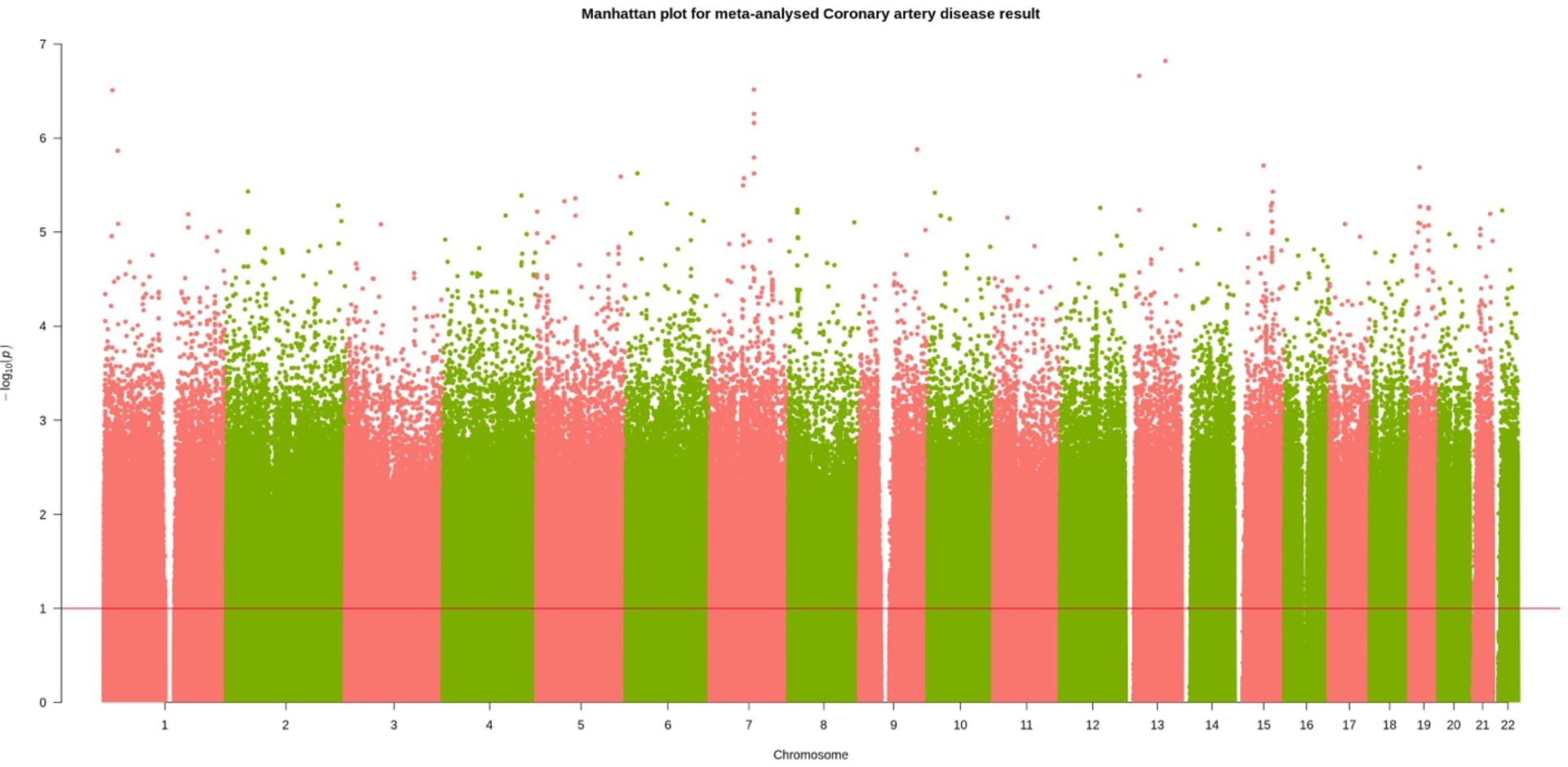
Manhattan plot for meta-analysed coronary artery disease mortality GWAS.

**Figure S5.**
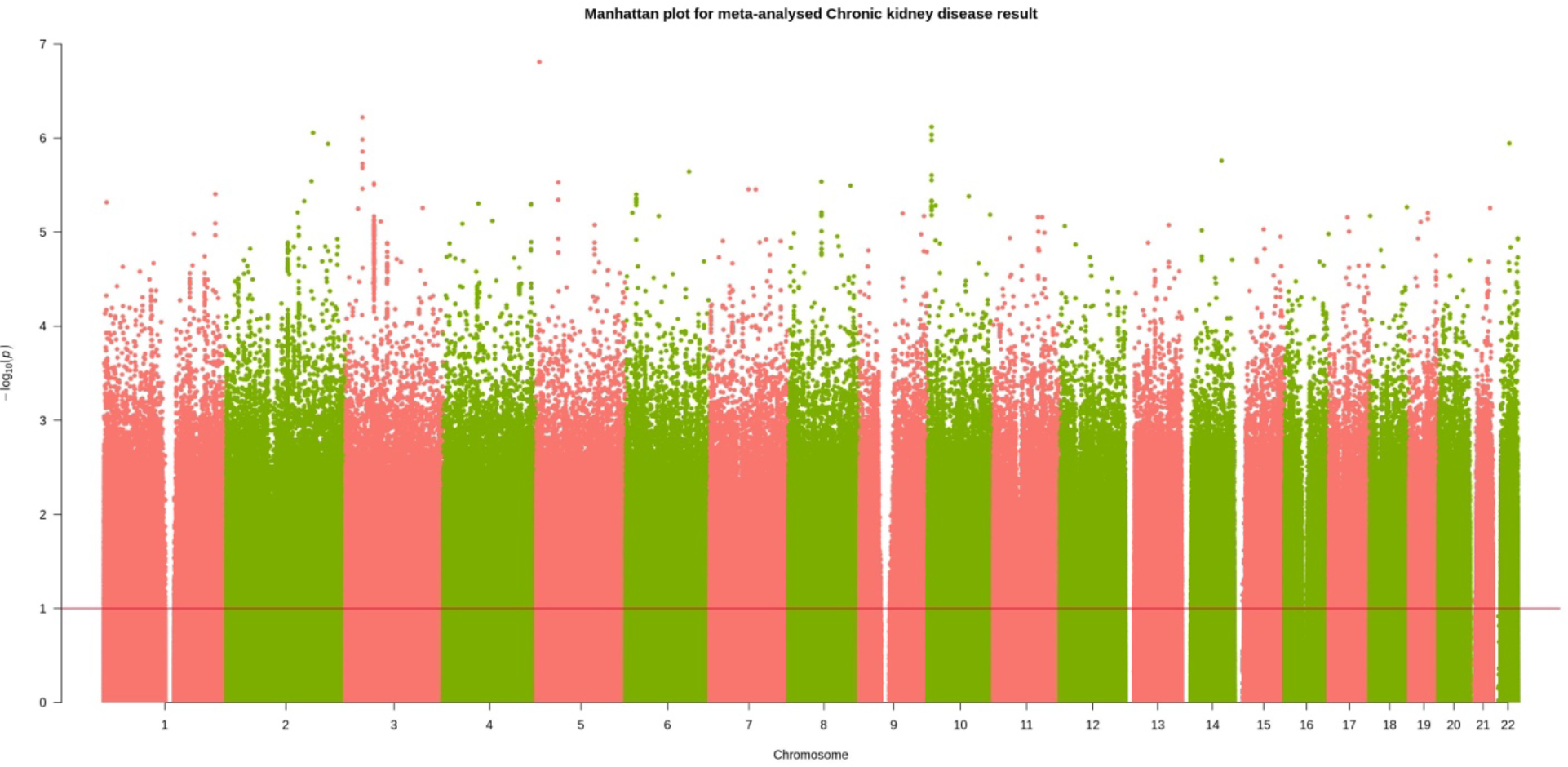
Manhattan plot for meta-analysed chronic kidney disease mortality GWAS.

**Figure S6.**
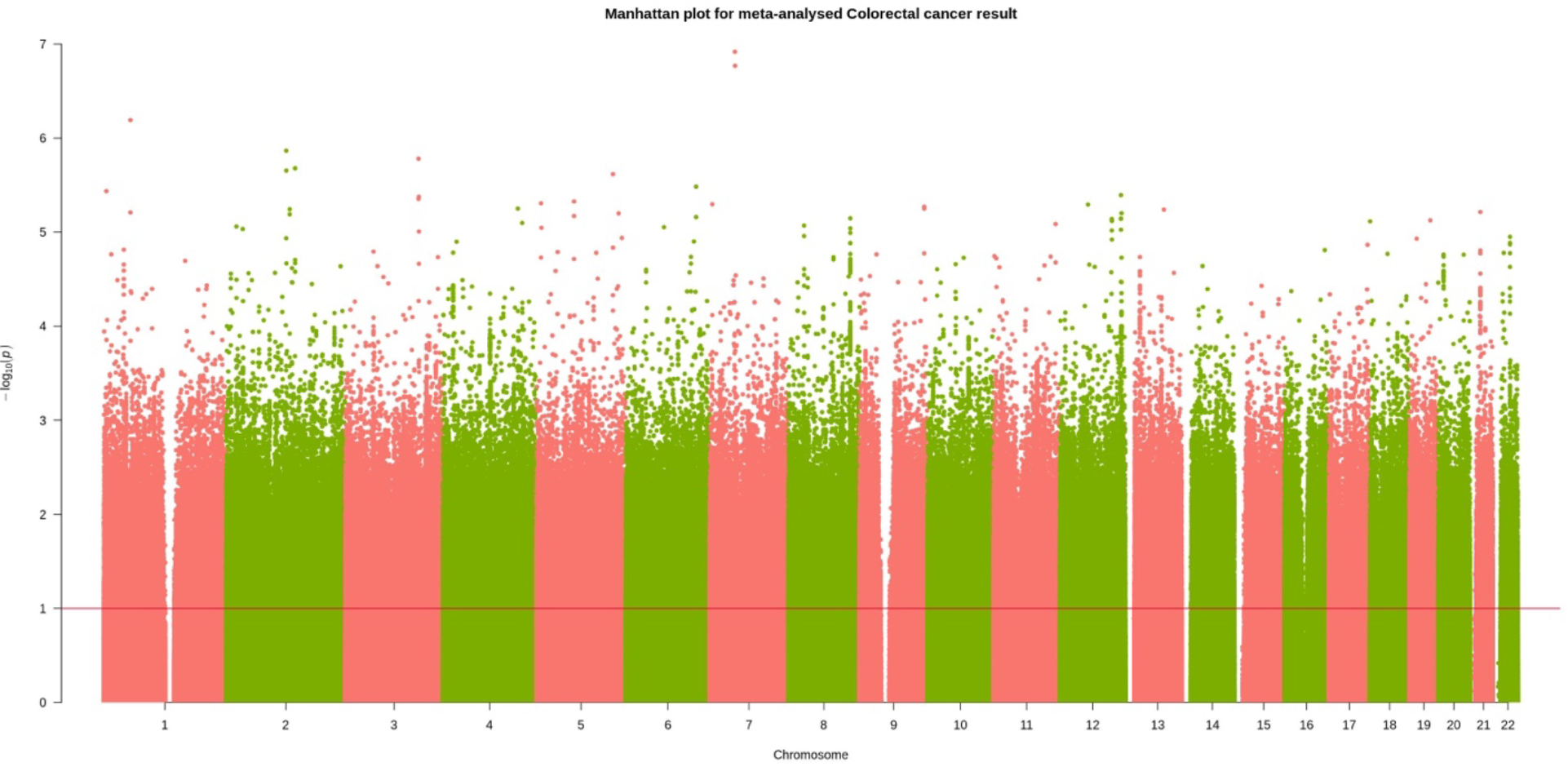
Manhattan plot for meta-analysed colorectal cancer mortality GWAS.

**Figure S7.**
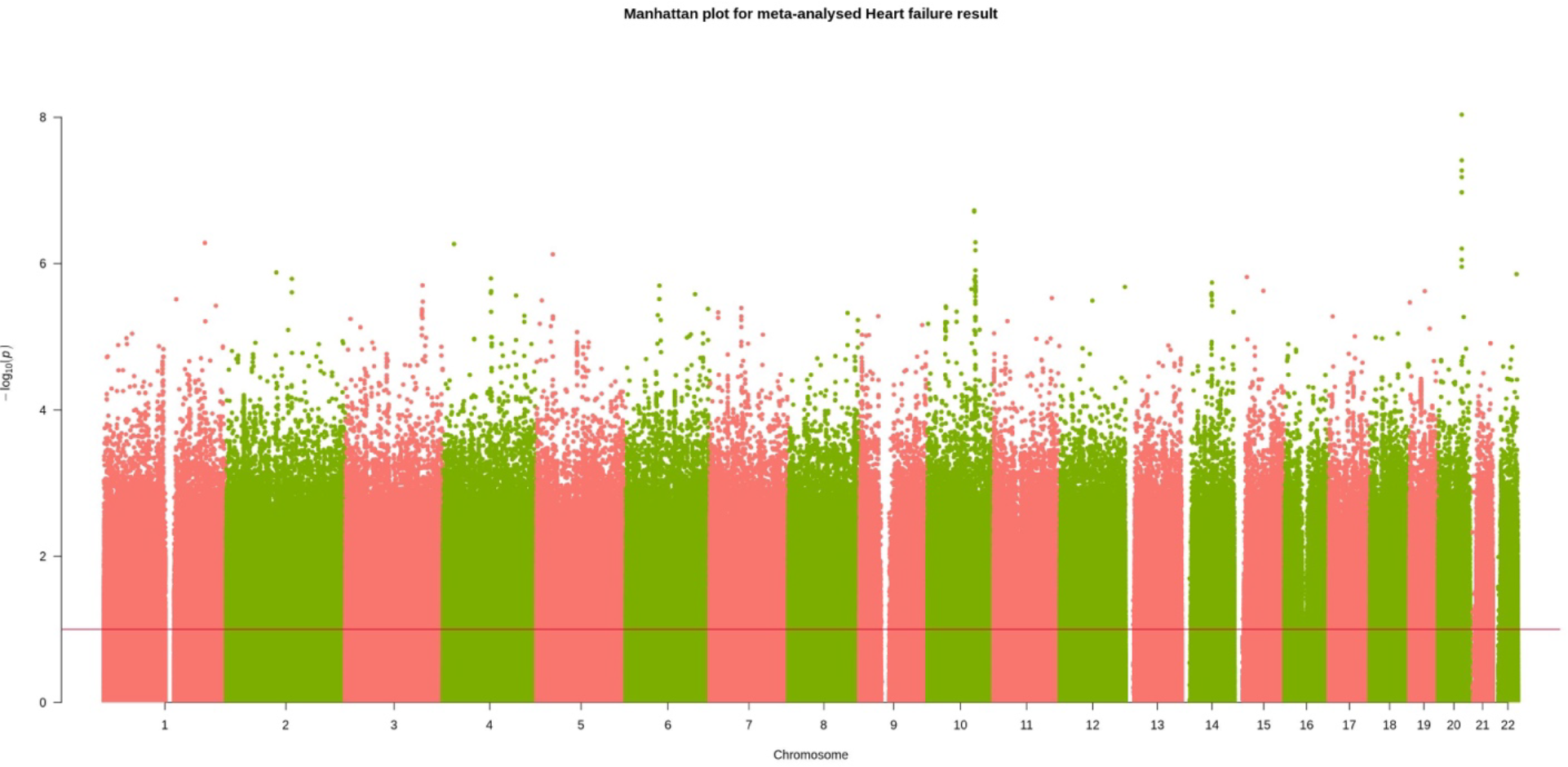
Manhattan plot for meta-analysed heart failure mortality GWAS.

**Figure S8.**
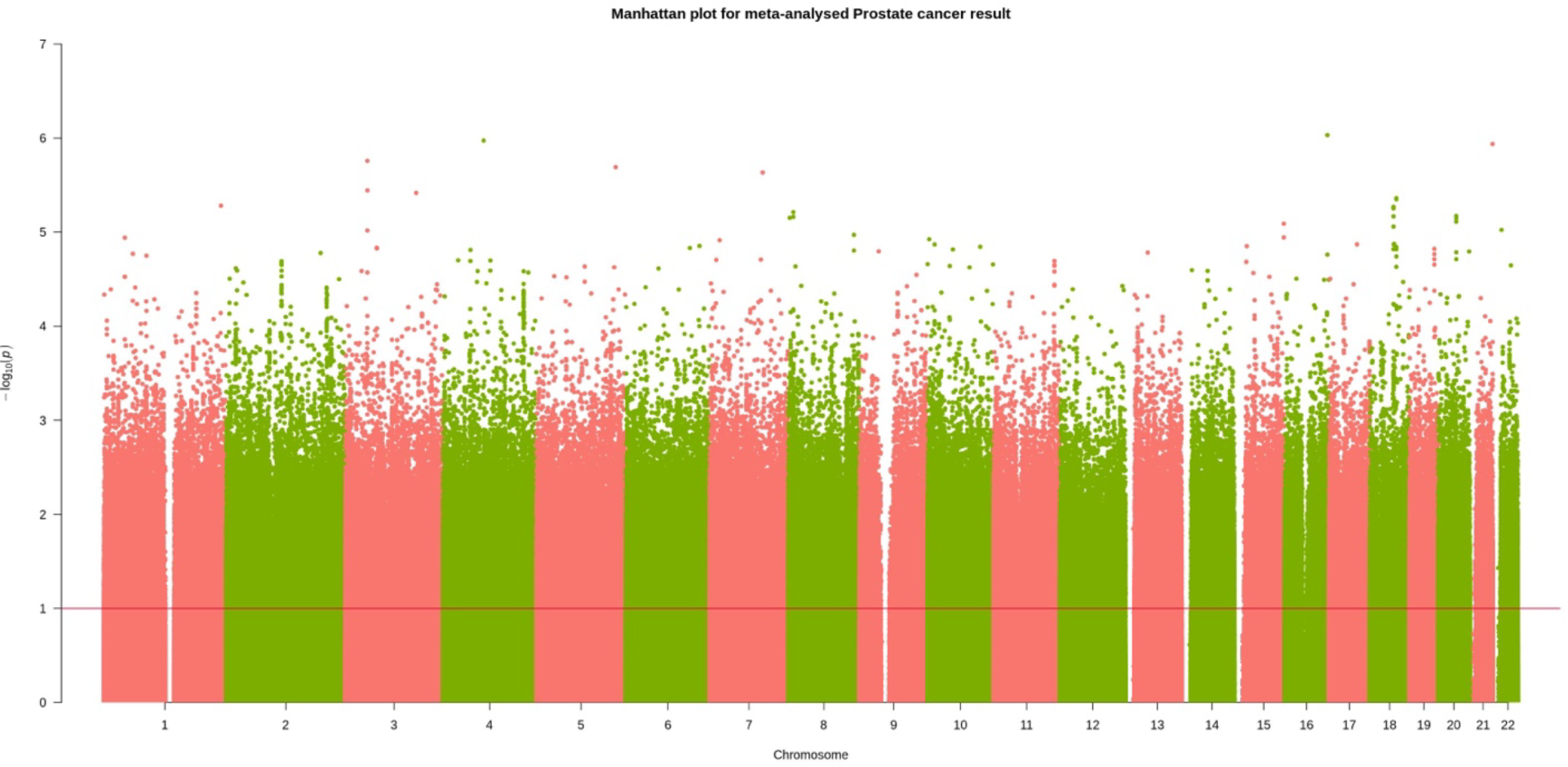
Manhattan plot for meta-analysed prostate cancer mortality GWAS.

**Figure S9.**
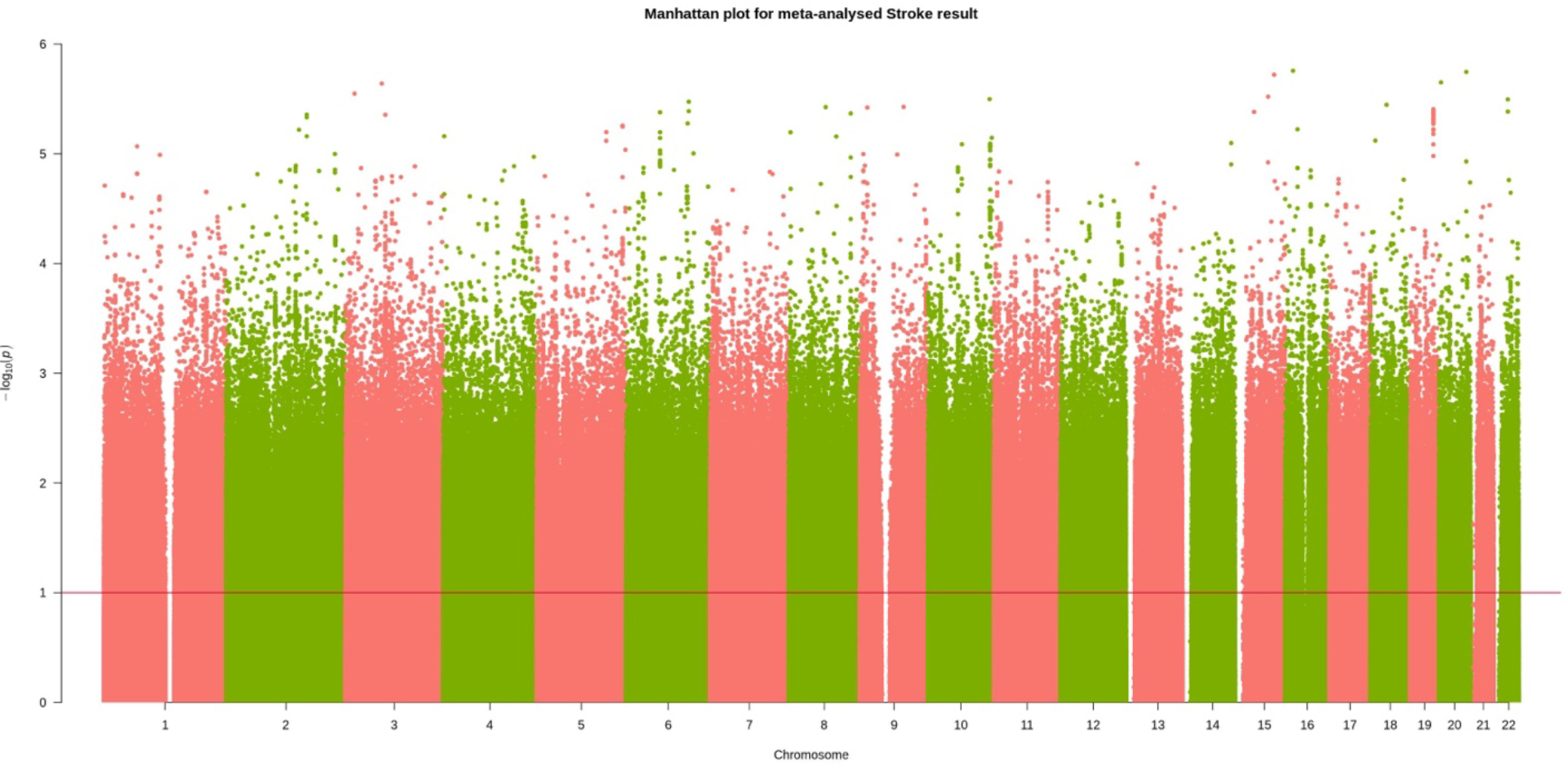
Manhattan plot for meta-analysed stroke mortality GWAS.

**Figure S10.**
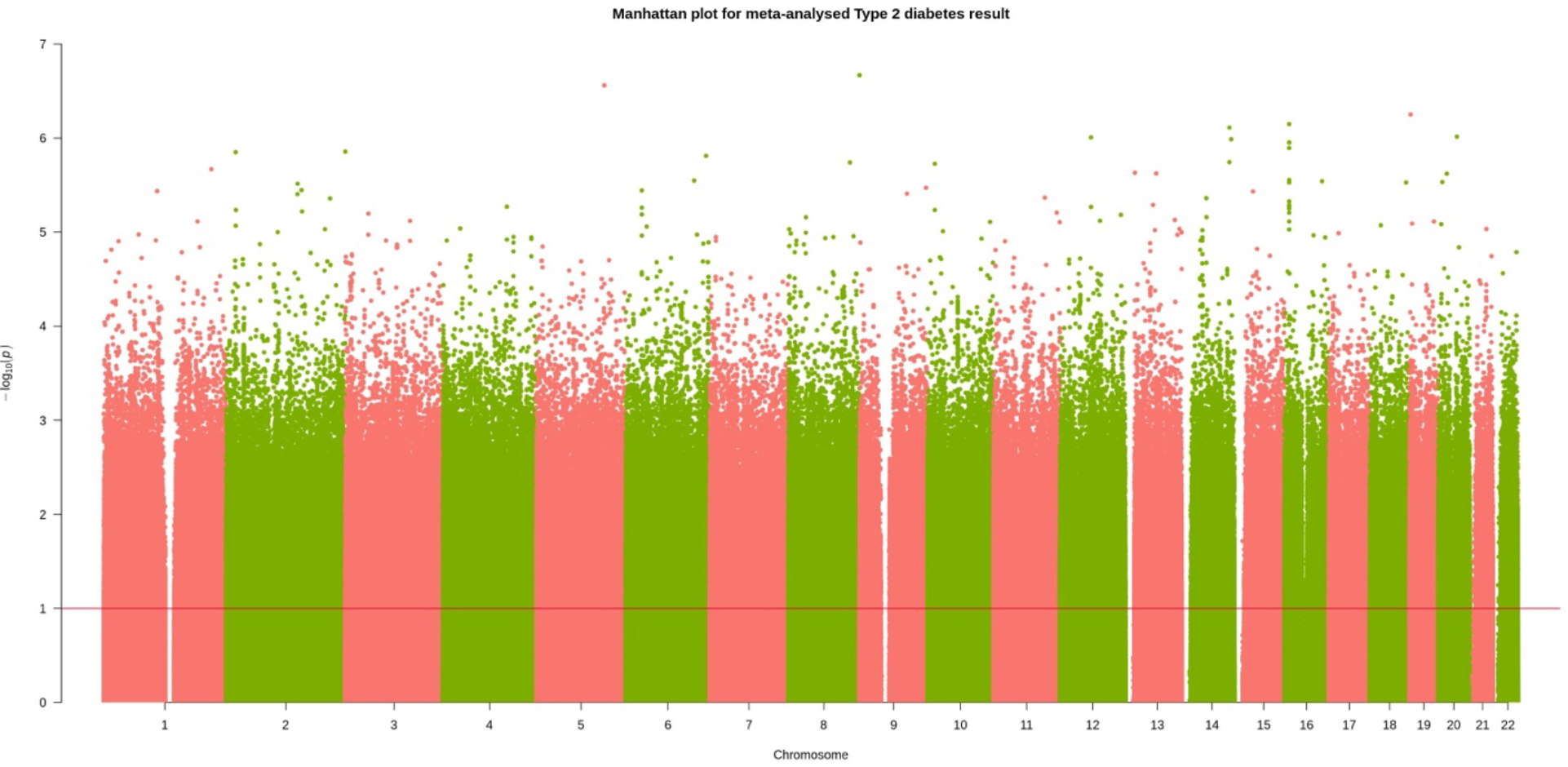
Manhattan plot for meta-analysed type ii diabetes mortality GWAS.

**Figure S11.**
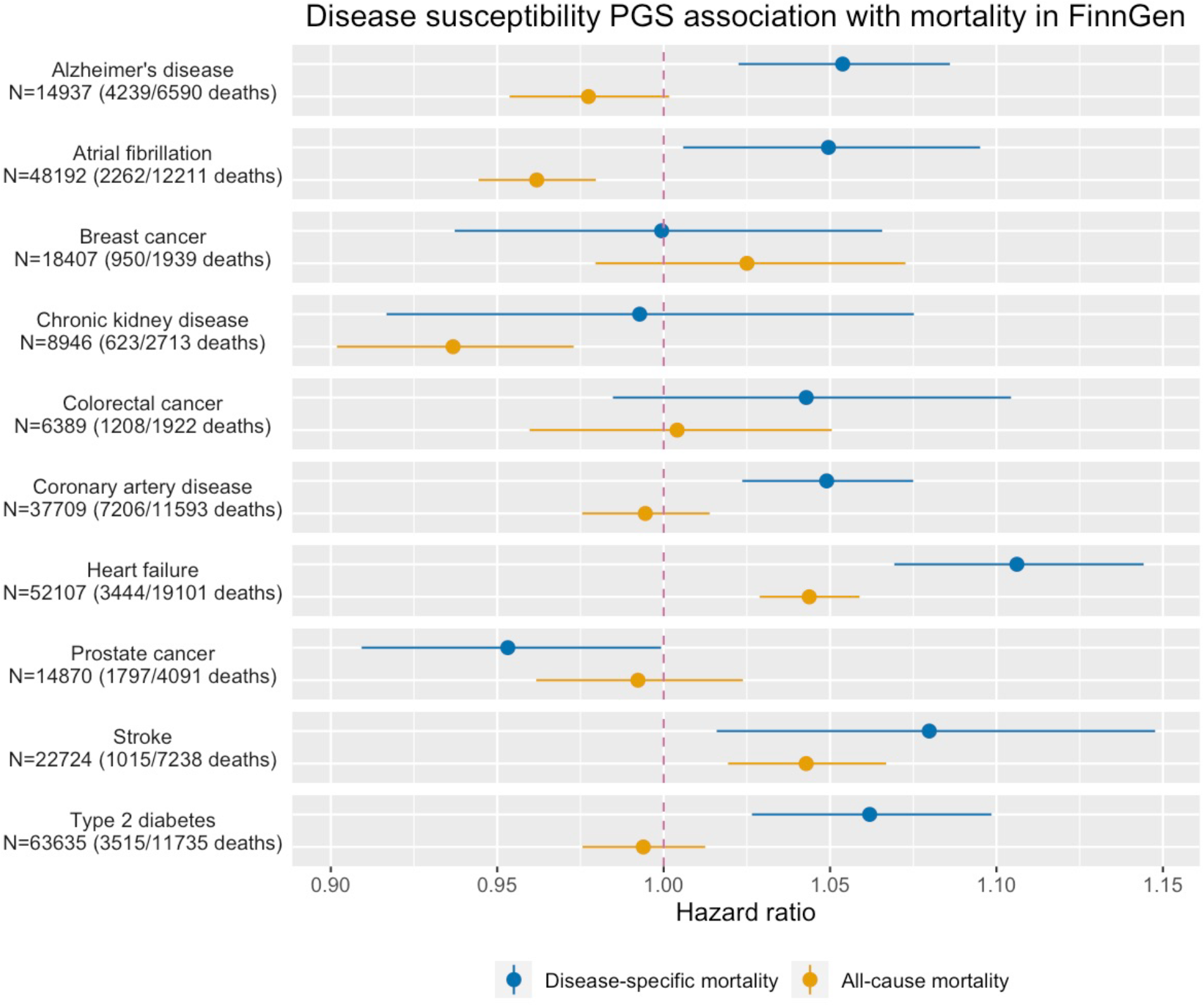
Comparison of disease susceptibility PGS association with disease-specific mortality and all-cause mortality in Finngen. In parenthesis stated number of disease-specific mortalities/number of all-cause mortality within the total number of patients (N). Horizontal solid lines represent 95% CI. Also see Table S15 for quantitative results.

**Figure S12.**
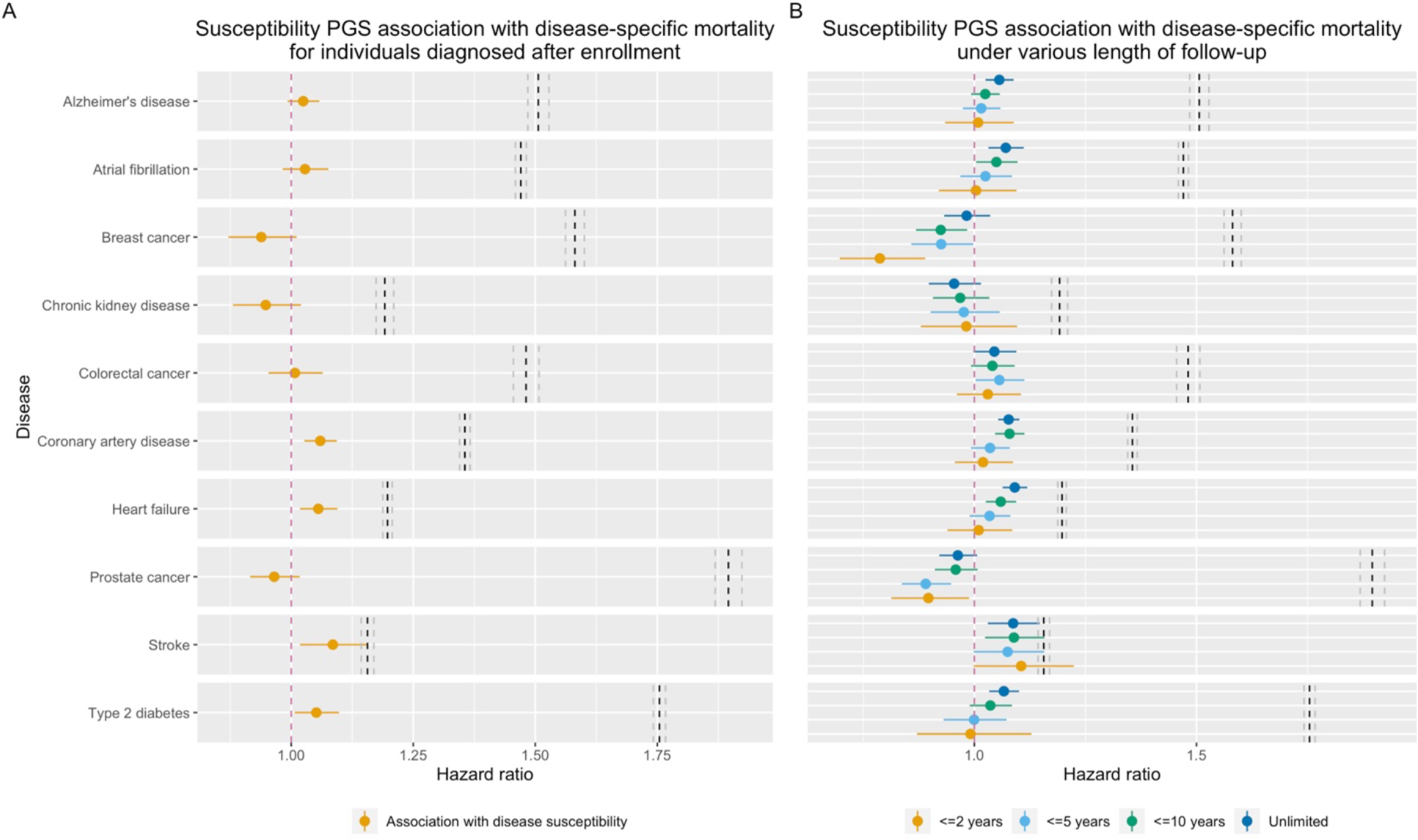
Sensitivity analyses. Left: Association between disease susceptibility PGS with disease specific mortality among only patients diagnosed after enrollment; Right: association between disease susceptibility PGS with disease specific mortality among patients with various lengths of follow-up after diagnosis. Horizontal solid lines represent 95% CI. Also see Table S16 for quantitative results.

**Figure S13.**
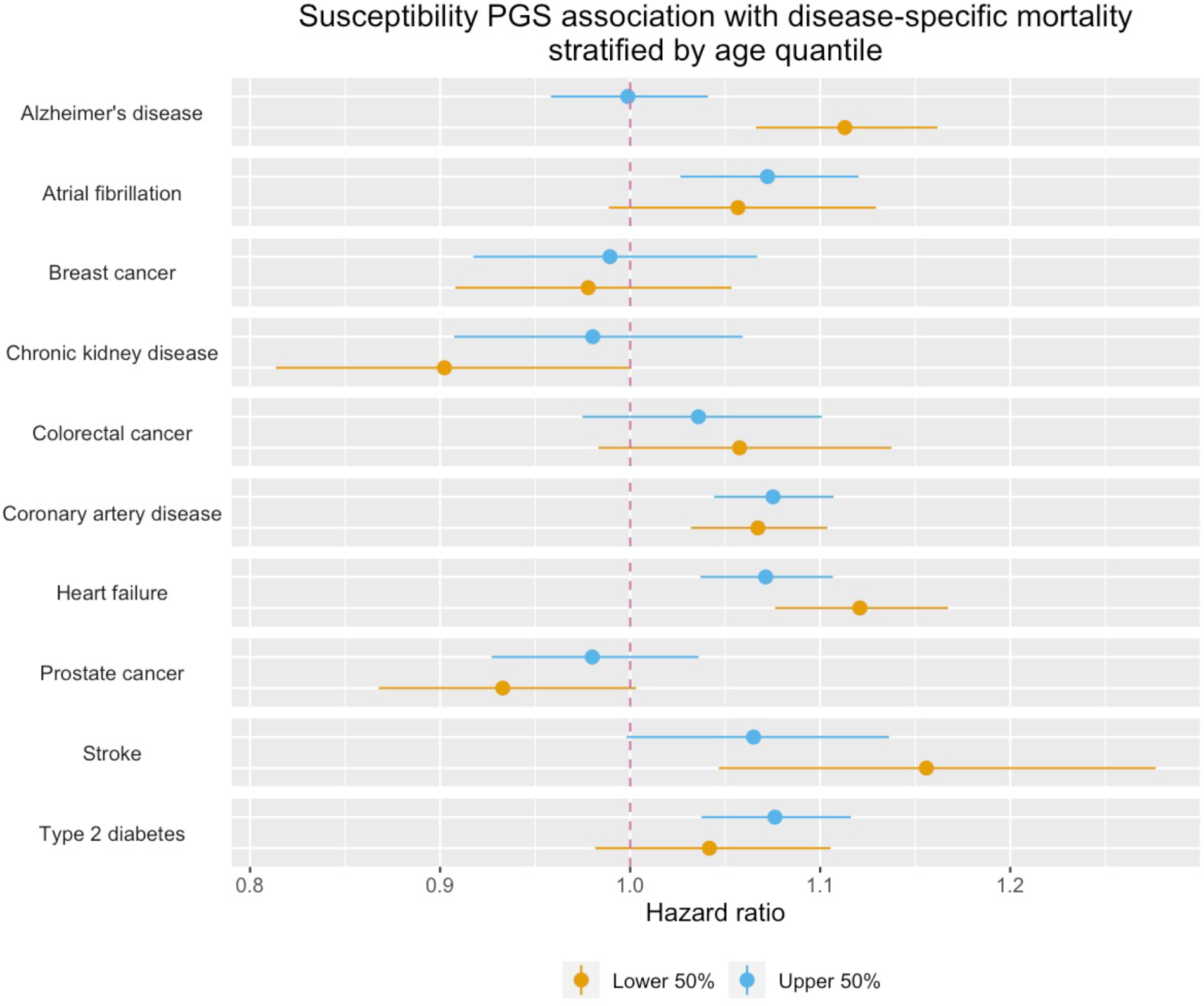
Sensitivity analyses. Susceptibility PGS association with disease-specific mortality stratified by age quantile. Horizontal solid lines represent 95% CI. Also see Table S17 for quantitative results.

**Figure S14.**
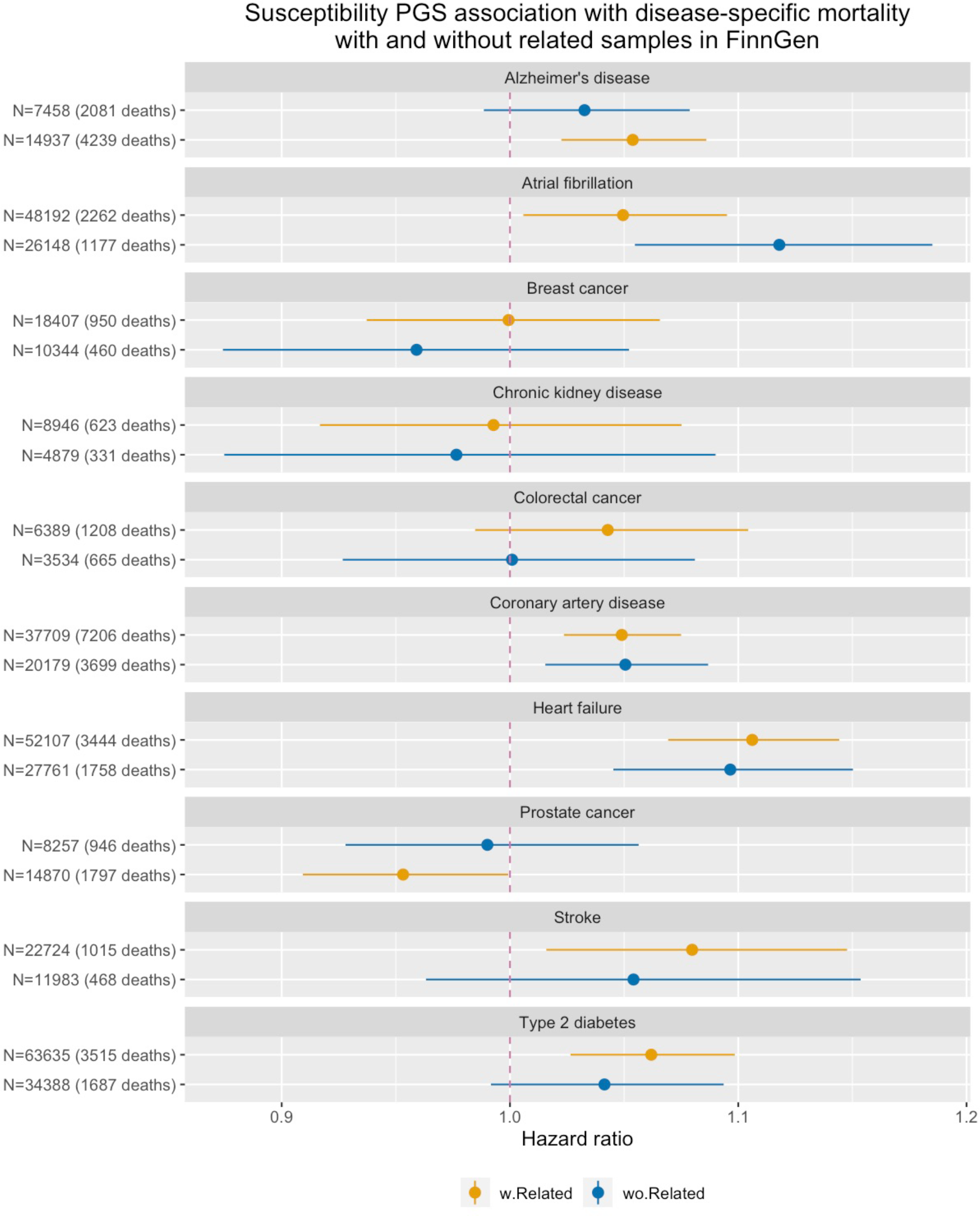
Sensitivity analyses. Susceptibility PGS association with disease-specific mortality in FinnGen with and without related individuals. For the without relatedness group (wo.Related), we removed up until second degree relatedness in the analyses. Horizontal solid lines represent 95% CI. Also see Table S15 for quantitative results.

**Figure S15.**
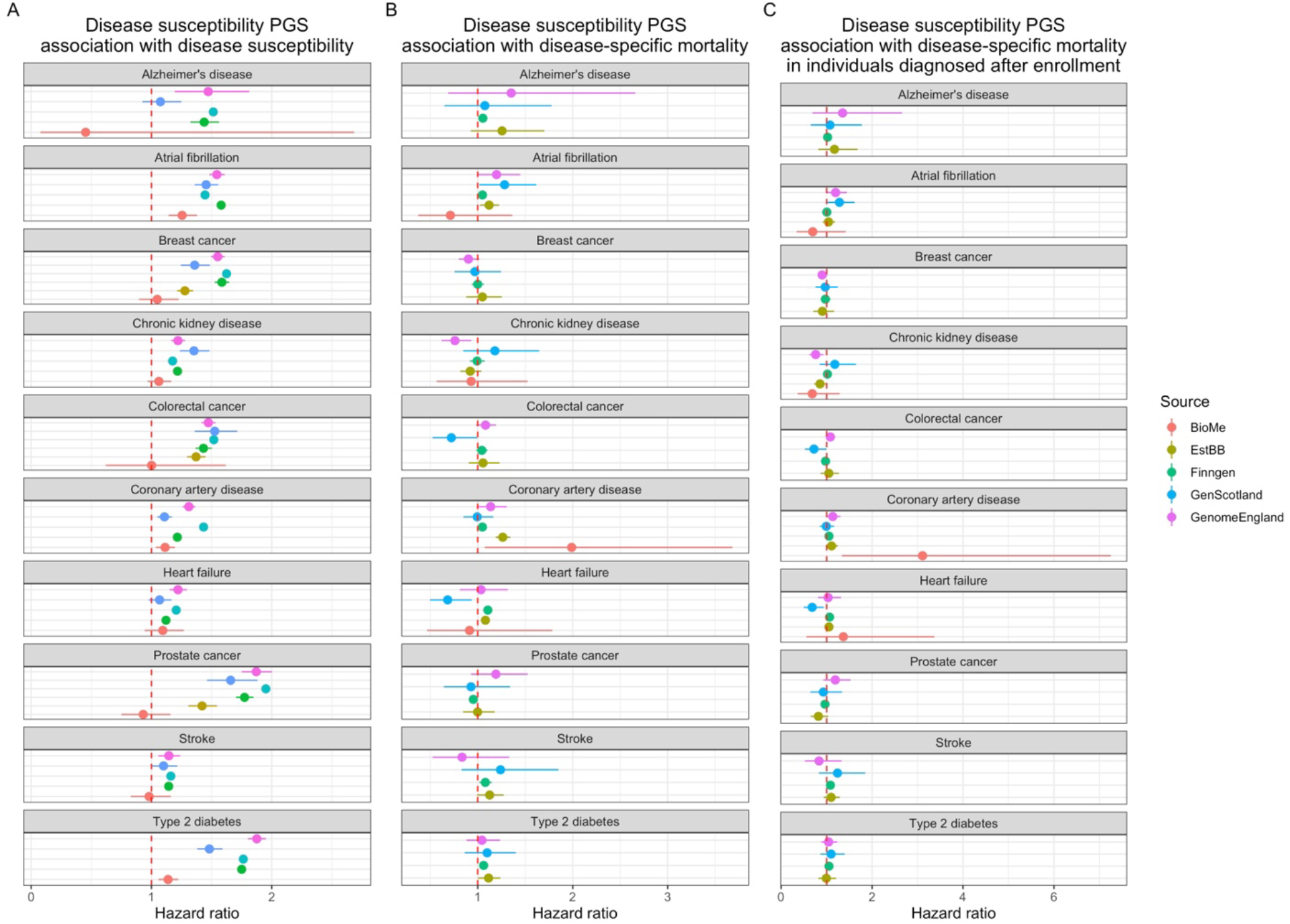
Forest plot for effect sizes from each participant biobank. Horizontal solid lines represent 95% CI.

**Figure S16.**
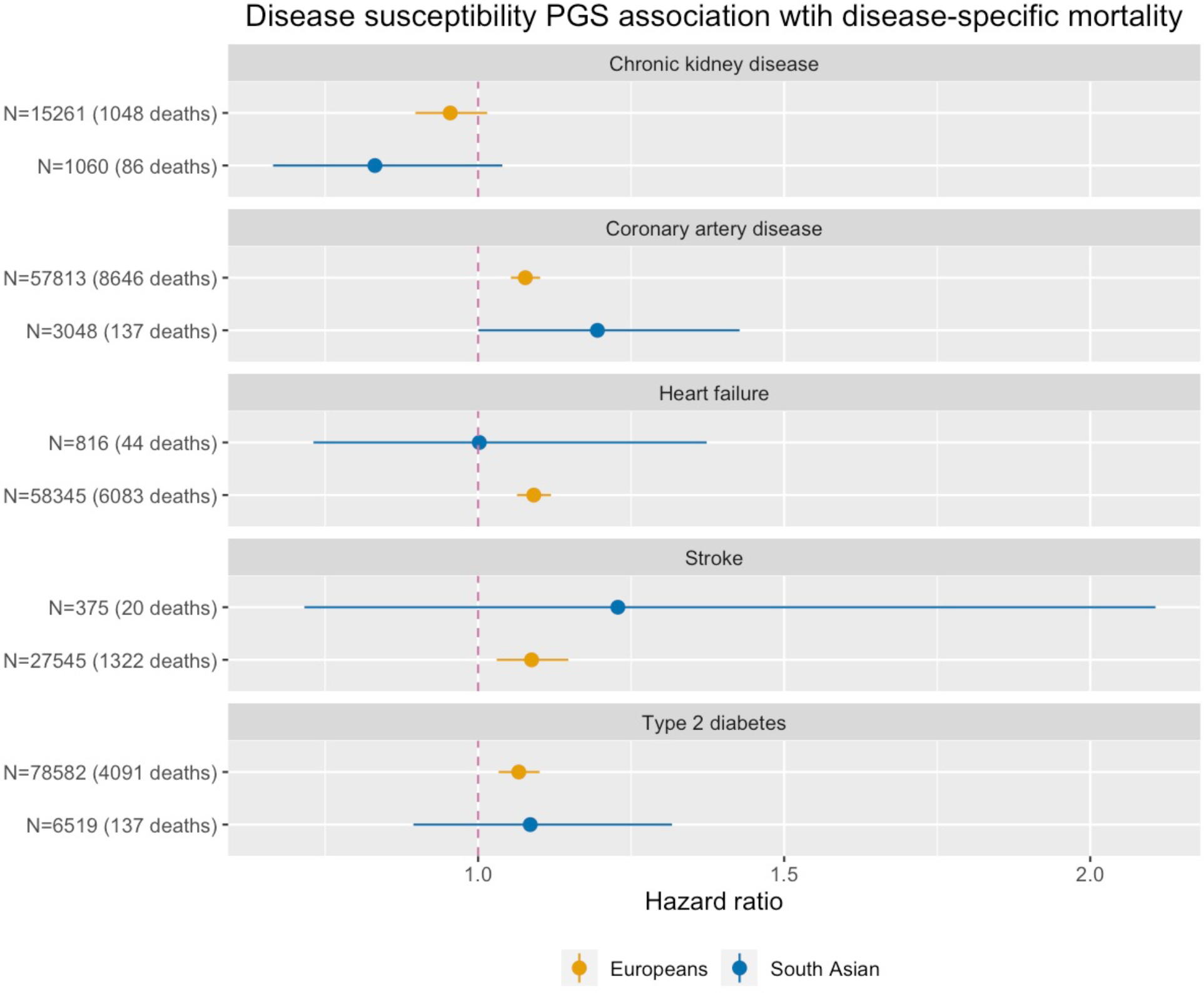
Disease susceptibility PGS association with disease-specific mortality in non-European population. As only patient cohorts are of interest in this study, for the non-European population, the only relatively powered results we had were associations for South Asians from biobank Genes & Health in a subset of diseases. Horizontal solid lines represent 95% CI.

**Figure S17.**
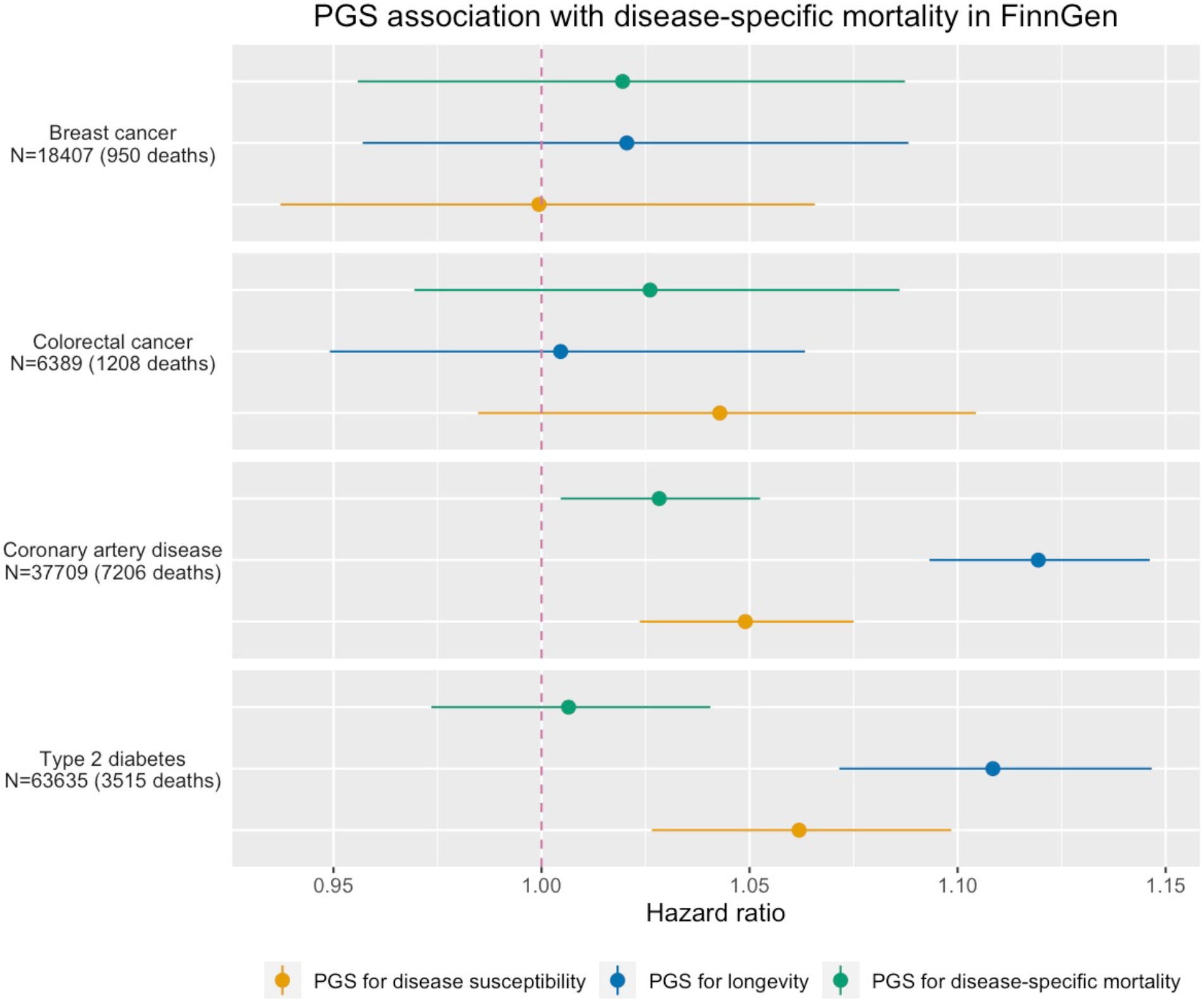
Association between PGS and disease-specific mortality in Finngen for eligible diseases. We constructed disease mortality PGS using meta-analysed mortality GWAS results with Finngen left out and evaluated its association with disease specific mortality in Finngen, comparing with disease diagnosis PGS and longevity PGS. Horizontal solid lines represent 95% CI. Also see Table S18 for quantitative results.

**Figure S18.**
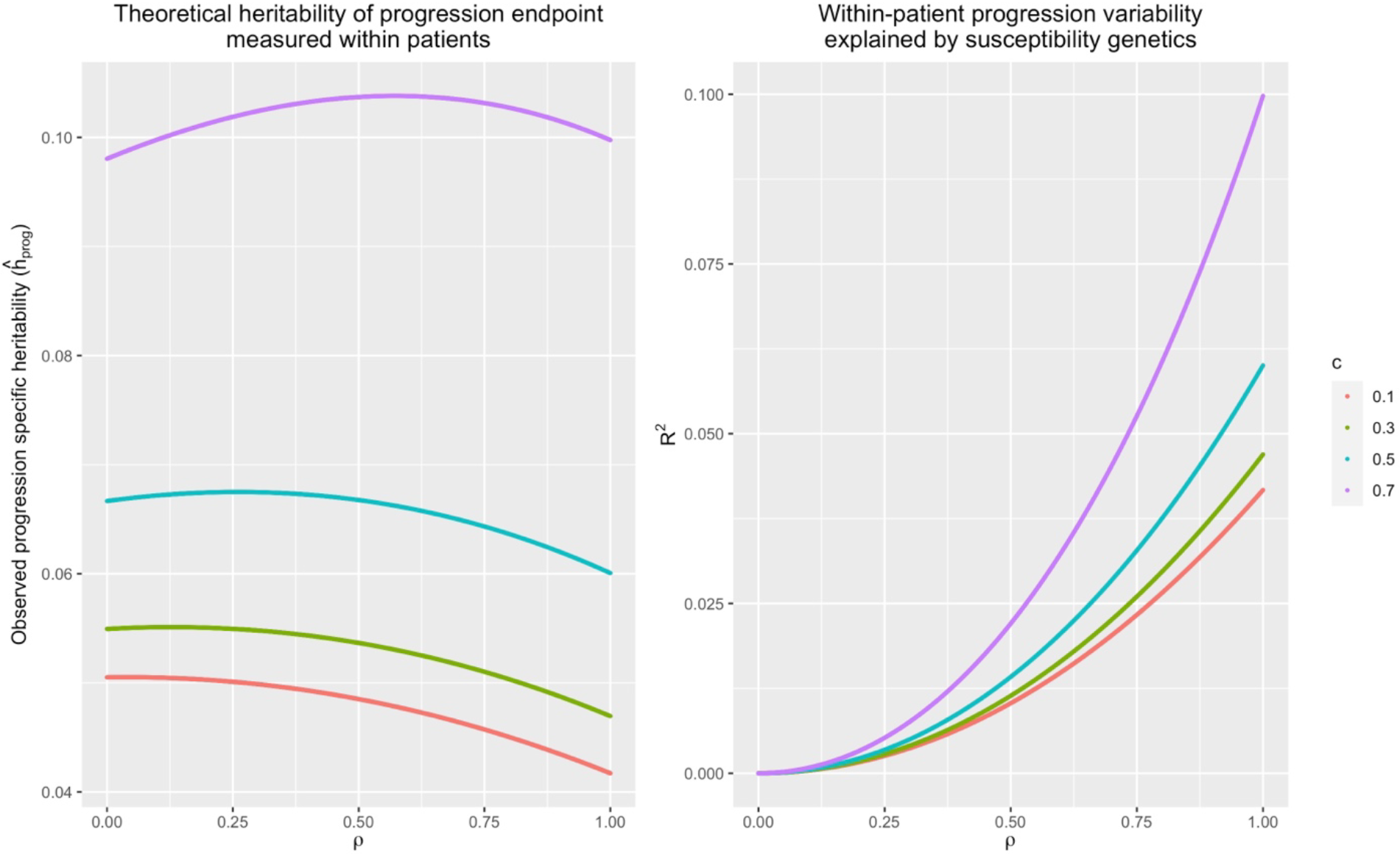
Theoretical derivation of expected genetic measurements. For both derivations, heritability for the genetic component of disease susceptibility (*h*_*sus*_) has been set to fixed value 0.3 and *unique genetic components* of progression endpoint in population (*h*_*prog*_) has been set to fixed value 0.1. Under varying contribution of disease susceptibility on progression liability (c) and correlation of the susceptibility and progression specific genetic component (ρ), we derive A. Theoretical within-patient heritability of disease progression, corresponding to the expected heritability can be observed from a within-patient progression GWAS; B. Theoretical patient progression variance explained by susceptibility genetics, corresponding to the expected *R*^*2*^ can be observed from disease susceptibility association with patients’ progression.

**Figure S19.**
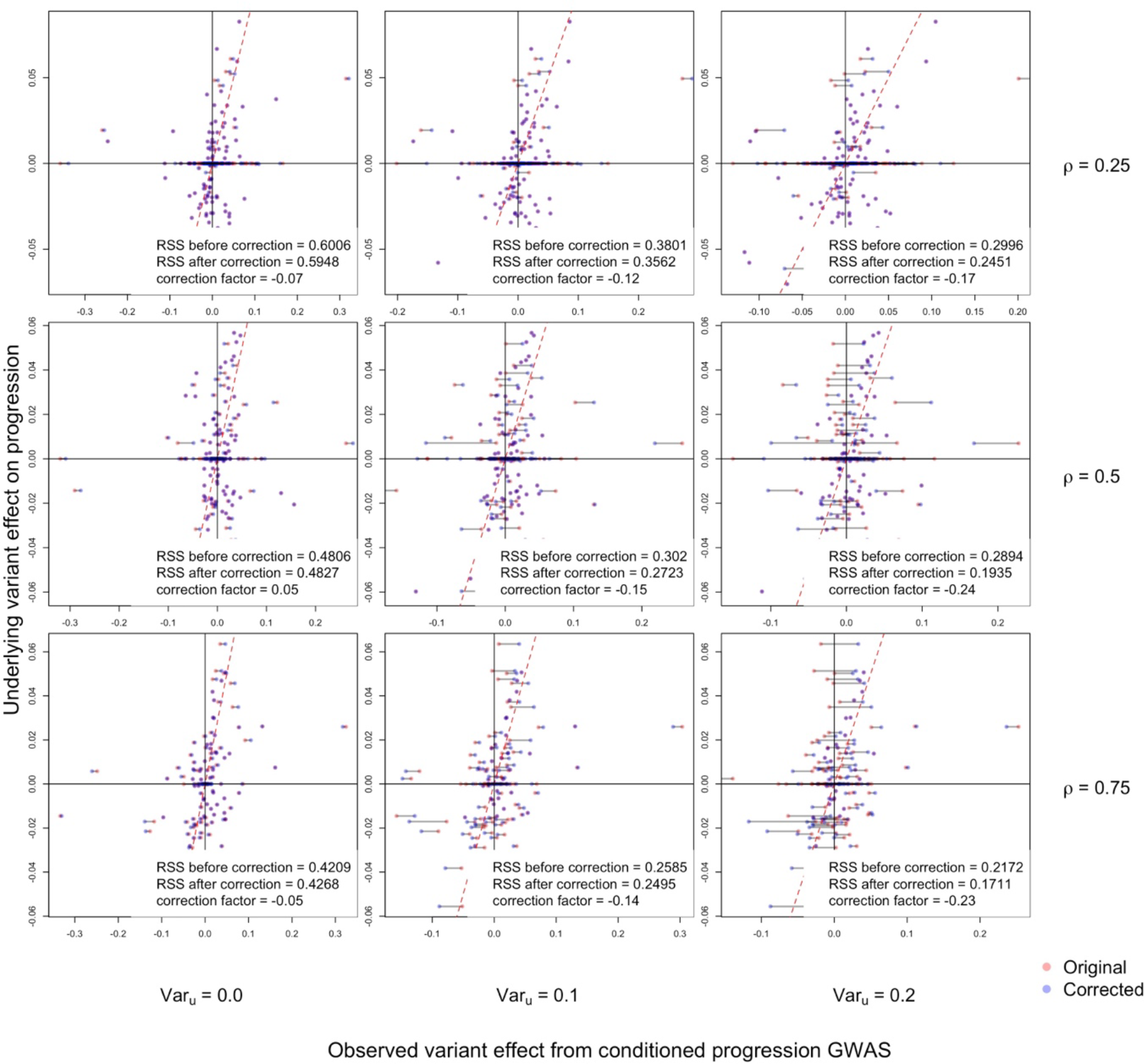
Impact of index event bias and slope-hunter-like correction under various conditions. In this experiment, we fixed heritability of disease susceptibility (*h*_*sus*_ = 0.2) and progression (*h*_*out*_ = 0.005). Impact of susceptibility liability on disease progression liability was also fixed at *c* = 0.3. Each panel corresponds to a scenario under certain amount of shared causal variants (ρ) and amount of shared non-genetic factor between the two endpoints (*Var*_*u*_). Plot shows alignment of GWAS observed variant effects (x-axis) with underlying causal effects (y-axis) on disease progression for all causal SNPs before and after slope-hunter-like correction on shared and susceptibility specific causal variants (note susceptibility specific causal variants are on y = 0 axis since their underlying effects on progression are 0). Also see Table S19 for quantitative results.

**Figure S20.**
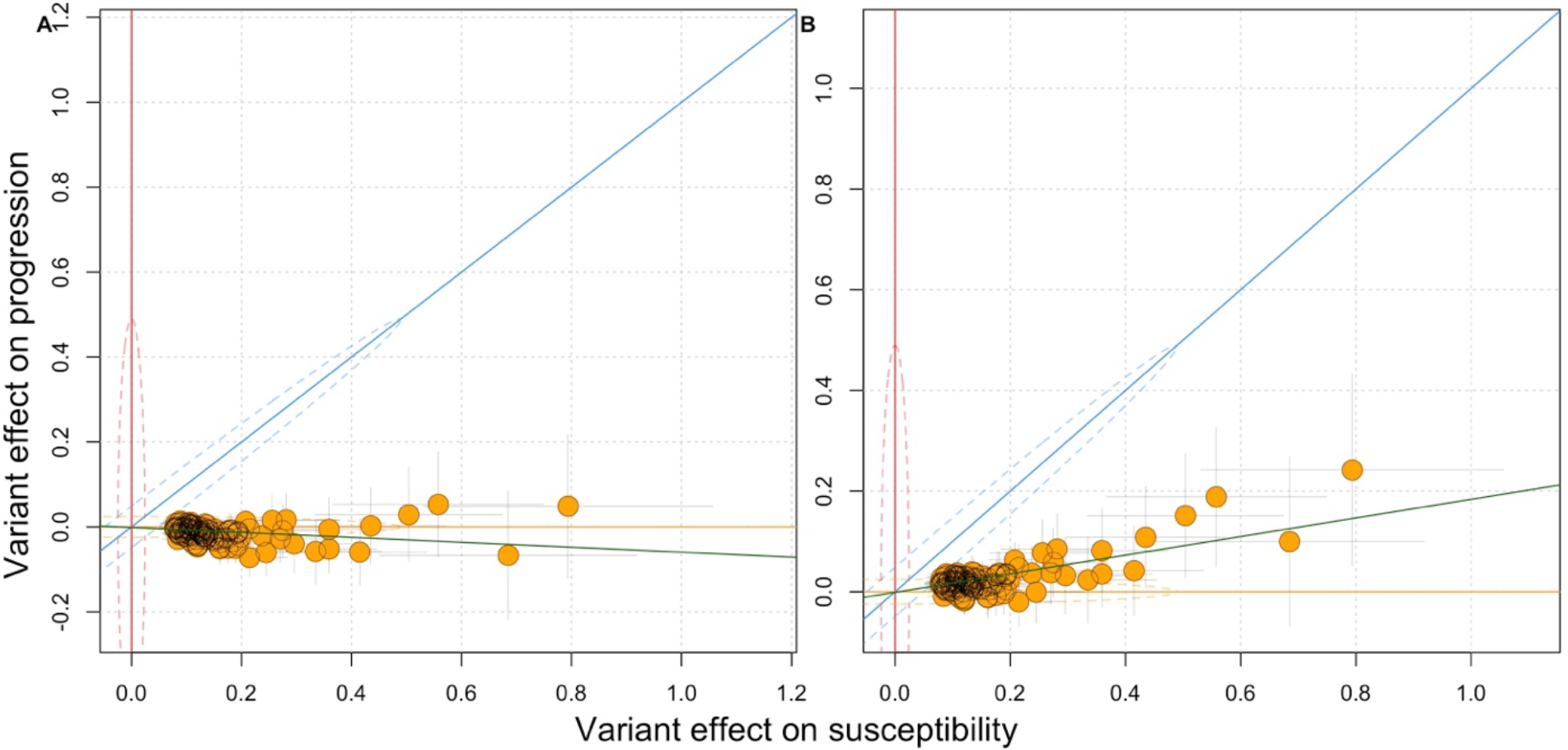
Impact of index event bias and slope-hunter-like correction on GWAS observation and SNP classification. From all configurations demonstrated in Figure S7, we chose one of the settings where we see most severe impact of index event bias (*Var*_*u*_ = 0.2, ρ = 0.5) and compared the linemodel (Pirinen, 2023) classification on GWAS results before (left) and after (right) correction. We plot variant effects before and after correction on disease progression (y-axis) against their effect on susceptibility (x-axis). The regression line (green line) shifted after correction, whereas there was no change in variant classification. Also see Table S19 for quantitative results.

## Footnotes

1 Here *m* could be the total number of variants in the human genome.

2 This measure has been widely used in prior work.

3 I.e., the disease susceptibility and the progression are exactly the same.

## Notes

### Competing Interest Statement

The authors have declared no competing interest.

### Funding Statement

This study has was funded by the European Union Horizon 2020 research innovation programme under grant agreement No 101016775, European Research Council (ERC) under the European Union Horizon 2020 research and innovation program (grant number 945733), and Academy of Finland fellowship grant N. 323116

### Author Declarations

Ethics Committee of the Helsinki and Uusimaa Hospital District, North West Centre for Research Ethics Committee, Tayside Committee on Medical Research Ethics, Scotland A Research Ethics Committee and Centre for Ethics of the University of Tartu gave ethical approval for this work.

## Reference

1000 Genomes Project Consortium. (2015). A global reference for human genetic variation. Nature. http://www.nature.com/nature/journal/v526/n7571/abs/nature15393.html

Abbafati, C., Abbas, K. M., Abbasi-Kangevari, M., Abd-Allah, F., Abdelalim, A., Abdollahi, M., Abdollahpour, I., Abegaz, K. H., Abolhassani, H., Aboyans, V., Abreu, L. G., Abrigo, M. R. M., Abualhasan, A., Abu-Raddad, L. J., Abushouk, A. I., Adabi, M., Adekanmbi, V., Adeoye, A. M., Adetokunboh, O. O., … Amini, S. (2020). Global burden of 369 diseases and injuries in 204 countries and territories, 1990–2019: a systematic analysis for the Global Burden of Disease Study 2019. The Lancet, 396(10258), 1204–1222. 10.1016/S0140-6736(20)30925-9/ATTACHMENT/1802C2B8-7CCC-467E-B4DD-92F466CF5E15/MMC2E.PDF

Barbieux, P., György, B., Gand, E., Saulnier, P. J., Ducrocq, G., Halimi, J. M., Feigerlova, E., Hulin-Delmotte, C., Llaty, P., Montaigne, D., Rigalleau, V., Roussel, R., Sosner, P., Zaoui, P., Ragot, S., Marre, M., Tregouët, D. A., & Hadjadj, S. (2019). No prognostic role of a GWAS-derived genetic risk score in renal outcomes for patients from French cohorts with type 1 and type 2 diabetes.Diabetes & Metabolism, 45(5), 494–497. 10.1016/J.DIABET.2018.01.016

Bellenguez, C., Strange, A., Freeman, C., Donnelly, P., & Spencer, C. C. A. (2012). A robust clustering algorithm for identifying problematic samples in genome-wide association studies. Bioinformatics, 28(1), 134–135. 10.1093/BIOINFORMATICS/BTR599

Bi, W., Fritsche, L. G., Mukherjee, B., Kim, S., & Lee, S. (2020). A Fast and Accurate Method for Genome-Wide Time-to-Event Data Analysis and Its Application to UK Biobank. American Journal of Human Genetics, 107(2). 10.1016/J.AJHG.2020.06.003

Bien, S. A., Wojcik, G. L., Zubair, N., Gignoux, C. R., Martin, A. R., Kocarnik, J. M., Martin, L. W., Buyske, S., Haessler, J., Walker, R. W., Cheng, I., Graff, M., Xia, L., Franceschini, N., Matise, T., James, R., Hindorff, L., Marchand, L. Le North, K. E., … Carlson, C. S. (2016). Strategies for Enriching Variant Coverage in Candidate Disease Loci on a Multiethnic Genotyping Array. PLOS ONE, 11(12), e0167758. 10.1371/JOURNAL.PONE.0167758

Browning, B. L., Tian, X., Zhou, Y., & Browning, S. R. (2021). Fast two-stage phasing of large-scale sequence data. American Journal of Human Genetics, 108(10), 1880–1890. 10.1016/J.AJHG.2021.08.005/ATTACHMENT/3B2363C6-FB5A-41B8-90D7-E8E171727BF0/MMC1.PDF

Bulik-Sullivan, B. K., Loh, P.-R., Finucane, H. K., Ripke, S., Yang, J., Patterson, N., Daly, M. J., Price, A. L., & Neale, B. M. (2015). LD Score regression distinguishes confounding from polygenicity in genome-wide association studies. Nature Genetics, 47(3), 291–295. 10.1038/ng.3211

Bycroft, C., Freeman, C., Petkova, D., Band, G., Elliott, L. T., Sharp, K., Motyer, A., Vukcevic, D., Delaneau, O., O’Connell, J., Cortes, A., Welsh, S., Young, A., Effingham, M., McVean, G., Leslie, S., Allen, N., Donnelly, P., & Marchini, J. (2018). The UK Biobank resource with deep phenotyping and genomic data. Nature, 562(7726), 203–209. 10.1038/s41586-018-0579-z

Chen, C. Y., Pollack, S., Hunter, D. J., Hirschhorn, J. N., Kraft, P., & Price, A. L. (2013). Improved ancestry inference using weights from external reference panels. Bioinformatics (Oxford, England), 29(11), 1399–1406. 10.1093/BIOINFORMATICS/BTT144

Cho, S. M. J., Koyama, S., Honigberg, M. C., Surakka, I., Haidermota, S., Ganesh, S., Patel, A. P., Bhattacharya, R., Lee, H., Kim, H. C., & Natarajan, P. (2023). Genetic, sociodemographic, lifestyle, and clinical risk factors of recurrent coronary artery disease events: a population-based cohort study. European Heart Journal. 10.1093/EURHEARTJ/EHAD380

Danecek, P., Auton, A., Abecasis, G., Albers, C. A., Banks, E., DePristo, M. A., Handsaker, R. E., Lunter, G., Marth, G. T., Sherry, S. T., McVean, G., & Durbin, R. (2011). The variant call format and VCFtools. Bioinformatics, 27(15), 2156–2158. 10.1093/BIOINFORMATICS/BTR330

Davies, R. W., Flint, J., Myers, S., & Mott, R. (2016). Rapid genotype imputation from sequence without reference panels. Nature Genetics 2016 48:8, 48(8), 965–969. 10.1038/ng.3594

Davies, R. W., Kucka, M., Su, D., Shi, S., Flanagan, M., Cunniff, C. M., Chan, Y. F., & Myers, S. (2021). Rapid genotype imputation from sequence with reference panels. Nature Genetics 2021 53:7, 53(7), 1104–1111. 10.1038/s41588-021-00877-0

Dey, R., Zhou, W., Kiiskinen, T., Havulinna, A., Elliott, A., Karjalainen, J., Kurki, M., Qin, A., Lee, S., Palotie, A., Neale, B., Daly, M., & Lin, X. (2022). Efficient and accurate frailty model approach for genome-wide survival association analysis in large-scale biobanks. Nature Communications 2022 13:1, 13(1), 1–13. 10.1038/s41467-022-32885-x

Dudbridge, F., Allen, R. J., Sheehan, N. A., Schmidt, A. F., Lee, J. C., Jenkins, R. G., Wain, L. V., Hingorani, A. D., & Patel, R. S. (2019). Adjustment for index event bias in genome-wide association studies of subsequent events. Nature Communications 2019 10:1, 10(1), 1–10. 10.1038/s41467-019-09381-w

Escala-Garcia, M., Guo, Q., Dörk, T., Canisius, S., Keeman, R., Dennis, J., Beesley, J., Lecarpentier, J., Bolla, M. K., Wang, Q., Abraham, J., Andrulis, I. L., Anton-Culver, H., Arndt, V., Auer, P. L., Beckmann, M. W., Behrens, S., Benitez, J., Bermisheva, M., … Schmidt, M. K. (2019). Genomewide association study of germline variants and breast cancer-specific mortality. British Journal of Cancer 2019 120:6, 120(6), 647–657. 10.1038/s41416-019-0393-x

Fahed, A. C., Philippakis, A. A., & Khera, A. V. (2022). The potential of polygenic scores to improve cost and efficiency of clinical trials. Nature Communications, 13(1). 10.1038/S41467-022-30675-Z

Feng, Y.-C. A., Ge, T., Cordioli, M., FinnGen Ganna, A., Smoller, J. W., & Neale, B. M. (2020). Findings and insights from the genetic investigation of age of first reported occurrence for complex disorders in the UK Biobank and FinnGen. MedRxiv, 2020.11.20.20234302. 10.1101/2020.11.20.20234302

Finer, S., Martin, H. C., Khan, A., Hunt, K. A., Maclaughlin, B., Ahmed, Z., Ashcroft, R., Durham, C., Macarthur, D. G., McCarthy, M. I., Robson, J., Trivedi, B., Griffiths, C., Wright, J., Trembath, R. C., & Van Heel, D. A. (2020). Cohort Profile: East London Genes & Health (ELGH), a communitybased population genomics and health study in British Bangladeshi and British Pakistani people. International Journal of Epidemiology, 49(1), 20–21i. 10.1093/IJE/DYZ174

FinnGen project | FinnGen. (n.d.). Retrieved September 13, 2023, from https://www.finngen.fi/en/for_researchers

Finucane, H. K., Bulik-Sullivan, B., Gusev, A., Trynka, G., Reshef, Y., Loh, P.-R., Anttila, V., Xu, H., Zang, C., Farh, K., Ripke, S., Day, F. R., Purcell, S., Stahl, E., Lindstrom, S., Perry, J. R. B., Okada, Y., Raychaudhuri, S., Daly, M. J., … Price, A. L. (2015). Partitioning heritability by functional annotation using genome-wide association summary statistics. Nature Genetics, 47(11), 1228–1235. 10.1038/ng.3404

Gibbs, R. A., Belmont, J. W., Hardenbol, P., Willis, T. D., Yu, F., Zhang, H., Zeng, C., Matsuda, I., Fukushima, Y., Macer, D. R., Suda, E., Stein, L. D., Cunningham, F., Kanani, A., Thorisson, G. A., Chakravarti, A., Chen, P. E., Cutler, D. J., Kashuk, C. S., … Tanaka, T. (2003). The International HapMap Project. Nature, 426(6968), 789–796. 10.1038/nature02168

Groha, S., Alaiwi, S. A., Xu, W., Naranbhai, V., Nassar, A. H., Bakouny, Z., El Zarif, T., Saliby, R. M., Wan, G., Rajeh, A., Adib, E., Nuzzo, P. V., Schmidt, A. L., Labaki, C., Ricciuti, B., Alessi, J. V., Braun, D. A., Shukla, S. A., Keenan, T. E., … Gusev, A. (2022). Germline variants associated with toxicity to immune checkpoint blockade. Nature Medicine, 28(12), 2584–2591. 10.1038/S41591-022-02094-6

Guo, Q., Schmidt, M. K., Kraft, P., Canisius, S., Chen, C., Khan, S., Tyrer, J., Bolla, M. K., Wang, Q., Dennis, J., Michailidou, K., Lush, M., Kar, S., Beesley, J., Dunning, A. M., Shah, M., Czene, K., Darabi, H., Eriksson, M., … Pharoah, P. D. P. (2015). Identification of Novel Genetic Markers of Breast Cancer Survival. JNCI: Journal of the National Cancer Institute, 107(5), 81. 10.1093/JNCI/DJV081

Gusev, A., Groha, S., Taraszka, K., Semenov, Y. R., & Zaitlen, N. (2021). Constructing germline research cohorts from the discarded reads of clinical tumor sequences. Genome Medicine, 13(1), 1–14. 10.1186/S13073-021-00999-4/FIGURES/6

Harroud, A., Stridh, P., McCauley, J. L., Saarela, J. R van den Bosch, A. M., Engelenburg, H. J., Beecham, A. H., Alfredsson, L., Alikhani, K., Amezcua, L. M Andlauer, T. F., Ban, M., Barcellos, L. F., Barizzone, N., Berge, T., Berthele, A., Bittner, S., Bos, S. D. S Briggs, F. B., … Stefamp, ri. (2023). Locus for severity implicates CNS resilience in progression of multiple sclerosis. Nature 2023, 1–9. 10.1038/s41586-023-06250-x

Hernesniemi, J. A. (2022). Dawn of the Era of Individualized Genetic Profiling in the Prevention of Sudden Cardiac Death. Journal of the American College of Cardiology, 80(9), 884–886. 10.1016/J.JACC.2022.06.016

Houlahan, K. E., Livingstone, J., Fox, N. S., Kurganovs, N., Zhu, H., Sietsma Penington, J., Jung, C.-H., Yamaguchi, T. N., Heisler, L. E., Jovelin, R., Costello, A. J., Pope, B. J., Kishan, A. U., Corcoran, N. M., Bristow, R. G., Waszak, S. M., Weischenfeldt, J., He, H. H., Hung, R. J., … Boutros, P. C. (2023). A polygenic two-hit hypothesis for prostate cancer. JNCI: Journal of the National Cancer Institute, 115(4), 468–472. 10.1093/JNCI/DJAD001

Howie, B. N., Donnelly, P., & Marchini, J. (2009). A Flexible and Accurate Genotype Imputation Method for the Next Generation of Genome-Wide Association Studies. PLOS Genetics, 5(6), e1000529. 10.1371/JOURNAL.PGEN.1000529

Hujoel, M. L. A., Gazal, S., Loh, P. R., Patterson, N., & Price, A. L. (2020). Liability threshold modeling of case–control status and family history of disease increases association power. Nature Genetics 2020 52:5, 52(5), 541–547. 10.1038/s41588-020-0613-6

International HapMap 3 Consortium, D. M., Altshuler, D. M., Gibbs, R. A., Peltonen, L., Altshuler, D. M., Gibbs, R. A., Peltonen, L., Dermitzakis, E., Schaffner, S. F., Yu, F., Peltonen, L., Dermitzakis, E., Bonnen, P. E., Altshuler, D. M., Gibbs, R. A., de Bakker, P. I. W., Deloukas, P., Gabriel, S. B., Gwilliam, R., … McEwen, J. E. (2010). Integrating common and rare genetic variation in diverse human populations. Nature, 467(7311), 52–58. 10.1038/nature09298

Jabbari, E., Koga, S., Valentino, R. R., Reynolds, R. H., Ferrari, R., Tan, M. M. X., Rowe, J. B., Dalgard, C. L., Scholz, S. W., Dickson, D. W., Warner, T. T., Revesz, T., Höglinger, G. U., Ross, O. A., Ryten, M., Hardy, J., Shoai, M., Morris, H. R., Mok, K. Y., … T.M. Hu, M. (2021). Genetic determinants of survival in progressive supranuclear palsy: a genome-wide association study. The Lancet. Neurology, 20(2), 107–116. 10.1016/S1474-4422(20)30394-X

Kember, R. L., Merikangas, A. K., Verma, S. S., Verma, A., Judy, R., Abecasis, G., Baras, A., Cantor, M., Coppola, G., Economides, A., Lotta, L., Overton, J. D., Reid, J. G., Shuldiner, A., Beechert, C., Forsythe, C., Fuller, E. D., Gu, Z., Lattari, M., … Bućan, M. (2021). Polygenic Risk of Psychiatric Disorders Exhibits Cross-trait Associations in Electronic Health Record Data From European Ancestry Individuals. Biological Psychiatry, 89(3), 236–245. 10.1016/J.BIOPSYCH.2020.06.026

Khan, Z., Di Nucci, F., Kwan, A., Hammer, C., Mariathasan, S., Rouilly, V., Carroll, J., Fontes, M., Acosta, S. L., Guardino, E., Chen-Harris, H., Bhangale, T., Mellman, I., Rosenberg, J., Powles, T., Hunkapiller, J., Chandler, G. S., & Albert, M. L. (2020). Polygenic risk for skin autoimmunity impacts immune checkpoint blockade in bladder cancer. Proceedings of the National Academy of Sciences of the United States of America, 117(22), 12288–12294. 10.1073/PNAS.1922867117/-/DCSUPPLEMENTAL

Khan, Z., Hammer, C., Carroll, J., Di Nucci, F., Acosta, S. L., Maiya, V., Bhangale, T., Hunkapiller, J., Mellman, I., Albert, M. L., McCarthy, M. I., & Chandler, G. S. (2021). Genetic variation associated with thyroid autoimmunity shapes the systemic immune response to PD-1 checkpoint blockade. Nature Communications, 12(1). 10.1038/S41467-021-23661-4

Kuhn, R. M., Haussler, D., & James Kent, W. (2013). The UCSC genome browser and associated tools. Briefings in Bioinformatics, 14(2), 144–161. 10.1093/BIB/BBS038

Kunkle, B. W., Grenier-Boley, B., Sims, R., Bis, J. C., Damotte, V., Naj, A. C., Boland, A., Vronskaya, M., van der Lee, S. J., Amlie-Wolf, A., Bellenguez, C., Frizatti, A., Chouraki, V., Martin, E. R., Sleegers, K., Badarinarayan, N., Jakobsdottir, J., Hamilton-Nelson, K. L., Moreno-Grau, S., … Pericak-Vance, M. A. (2019). Genetic meta-analysis of diagnosed Alzheimer’s disease identifies new risk loci and implicates A*β*, tau, immunity and lipid processing. Nature Genetics 2019 51:3, 51(3), 414–430. 10.1038/s41588-019-0358-2

Kurki, M. I., Karjalainen, J., Palta, P., Sipilä, T. P., Kristiansson, K., Donner, K. M., Reeve, M. P., Laivuori, H., Aavikko, M., Kaunisto, M. A., Loukola, A., Lahtela, E., Mattsson, H., Laiho, P., Della Briotta Parolo, P., Lehisto, A. A., Kanai, M., Mars, N., Rämö, J., … Palotie, A. (2023). FinnGen provides genetic insights from a well-phenotyped isolated population. Nature 2023 613:7944, 613(7944), 508–518. 10.1038/s41586-022-05473-8

Law, P. J., Timofeeva, M., Fernandez-Rozadilla, C., Broderick, P., Studd, J., Fernandez-Tajes, J., Farrington, S., Svinti, V., Palles, C., Orlando, G., Sud, A., Holroyd, A., Penegar, S., Theodoratou, E., Vaughan-Shaw, P., Campbell, H., Zgaga, L., Hayward, C., Campbell, A., … Dunlop, M. G. (2019). Association analyses identify 31 new risk loci for colorectal cancer susceptibility. Nature Communications 2019 10:1, 10(1), 1–15. 10.1038/s41467-019-09775-w

Lee, J. C., Biasci, D., Roberts, R., Gearry, R. B., Mansfield, J. C., Ahmad, T., Prescott, N. J., Satsangi, J., Wilson, D. C., Jostins, L., Anderson, C. A., Traherne, J. A., Lyons, P. A., Parkes, M., & Smith, K. G. C. (2017). Genome-wide association study identifies distinct genetic contributions to prognosis and susceptibility in Crohn’s disease. Nature Genetics 2017 49:2, 49(2), 262–268. 10.1038/ng.3755

Leitsalu, L., Haller, T., Esko, T., Tammesoo, M. L., Alavere, H., Snieder, H., Perola, M., Ng, P. C., Mägi, R., Milani, L., Fischer, K., & Metspalu, A. (2015). Cohort Profile: Estonian Biobank of the Estonian Genome Center, University of Tartu. International Journal of Epidemiology, 44(4), 1137–1147. 10.1093/IJE/DYT268

Liu, G., Peng, J., Liao, Z., Locascio, J. J., Corvol, J. C., Zhu, F., Dong, X., Maple-Grødem, J., Campbell, M. C., Elbaz, A., Lesage, S., Brice, A., Mangone, G., Growdon, J. H., Hung, A. Y., Schwarzschild,M. A., Hayes, M. T., Wills, A. M., Herrington, T. M., … Scherzer, C. R. (2021). Genome-wide survival study identifies a novel synaptic locus and polygenic score for cognitive progression in Parkinson’s disease. Nature Genetics, 53(6), 787–793. 10.1038/S41588-021-00847-6

Liu, S., Huang, S., Chen, F., Zhao, L., Yuan, Y., Francis, S. S., Fang, L., Li, Z., Lin, L., Liu, R., Zhang, Y., Xu, H., Li, S., Zhou, Y., Davies, R. W., Liu, Q., Walters, R. G., Lin, K., Ju, J., … Xu, X. (2018). Genomic Analyses from Non-invasive Prenatal Testing Reveal Genetic Associations, Patterns of Viral Infections, and Chinese Population History. Cell, 175(2), 347–359.e14. 10.1016/J.CELL.2018.08.016

Loh, P. R., Danecek, P., Palamara, P. F., Fuchsberger, C., Reshef, Y. A., Finucane, H. K., Schoenherr, S., Forer, L., McCarthy, S., Abecasis, G. R., Durbin, R., & Price, A. L. (2016). Reference-based phasing using the Haplotype Reference Consortium panel. Nature Genetics 2016 48:11, 48(11), 1443–1448. 10.1038/ng.3679

Mahajan, A., Taliun, D., Thurner, M., Robertson, N. R., Torres, J. M., Rayner, N. W., Payne, A. J., Steinthorsdottir, V., Scott, R. A., Grarup, N., Cook, J. P., Schmidt, E. M., Wuttke, M., Sarnowski, C., Mägi, R., Nano, J., Gieger, C., Trompet, S., Lecoeur, C., … McCarthy, M. I. (2018). Finemapping type 2 diabetes loci to single-variant resolution using high-density imputation and islet-specific epigenome maps. Nature Genetics 2018 50:11, 50(11), 1505–1513. 10.1038/s41588-018-0241-6

Mahmoud, O., Dudbridge, F., Davey Smith, G., Munafo, M., & Tilling, K. (2022). A robust method for collider bias correction in conditional genome-wide association studies. Nature Communications 2022 13:1, 13(1), 1–13. 10.1038/s41467-022-28119-9

Malik, R., Chauhan, G., Traylor, M., Sargurupremraj, M., Okada, Y., Mishra, A., Rutten-Jacobs, L., Giese, A. K., Van Der Laan, S. W., Gretarsdottir, S., Anderson, C. D., Chong, M., Adams, H. H. H., Ago, T., Almgren, P., Amouyel, P., Ay, H., Bartz, T. M., Benavente, O. R., … Yamaji, T. (2018). Multiancestry genome-wide association study of 520,000 subjects identifies 32 loci associated with stroke and stroke subtypes. Nature Genetics 2018 50:4, 50(4), 524–537. 10.1038/s41588-018-0058-3

Manichaikul, A., Mychaleckyj, J. C., Rich, S. S., Daly, K., Sale, M., & Chen, W. M. (2010). Robust relationship inference in genome-wide association studies. Bioinformatics, 26(22), 2867–2873. 10.1093/BIOINFORMATICS/BTQ559

McCarthy, S., Das, S., Kretzschmar, W., Delaneau, O., Wood, A. R., Teumer, A., Kang, H. M., Fuchsberger, C., Danecek, P., Sharp, K., Luo, Y., Sidore, C., Kwong, A., Timpson, N., Koskinen, S., Vrieze, S., Scott, L. J., Zhang, H., Mahajan, A., … Marchini, J. (2016). A reference panel of 64,976 haplotypes for genotype imputation. Nature Genetics 2016 48:10, 48(10), 1279–1283. 10.1038/ng.3643

Michailidou, K., Lindström, S., Dennis, J., Beesley, J., Hui, S., Kar, S., Lemaçon, A., Soucy, P., Glubb, D., Rostamianfar, A., Bolla, M. K., Wang, Q., Tyrer, J., Dicks, E., Lee, A., Wang, Z., Allen, J., Keeman, R., Eilber, U., … Easton, D. F. (2017). Association analysis identifies 65 new breast cancer risk loci. Nature, 551(7678), 92–94. 10.1038/nature24284

Nelson, C. P., Goel, A., Butterworth, A. S., Kanoni, S., Webb, T. R., Marouli, E., Zeng, L., Ntalla, I., Lai, F. Y., Hopewell, J. C., Giannakopoulou, O., Jiang, T., Hamby, S. E., Di Angelantonio, E., Assimes, T. L., Bottinger, E. P., Chambers, J. C., Clarke, R., Palmer, C. N. A., … Deloukas, P. (2017). Association analyses based on false discovery rate implicate new loci for coronary artery disease. Nature Genetics 2017 49:9, 49(9), 1385–1391. 10.1038/ng.3913

O’Connell, J., Gurdasani, D., Delaneau, O., Pirastu, N., Ulivi, S., Cocca, M., Traglia, M., Huang, J., Huffman, J. E., Rudan, I., McQuillan, R., Fraser, R. M., Campbell, H., Polasek, O., Asiki, G., Ekoru, K., Hayward, C., Wright, A. F., Vitart, V., … Marchini, J. (2014). A General Approach for Haplotype Phasing across the Full Spectrum of Relatedness. PLOS Genetics, 10(4), e1004234. 10.1371/JOURNAL.PGEN.1004234

Pan, G., Simpson, S., Van Der Mei, I., Charlesworth, J. C., Lucas, R., Ponsonby, A. L., Zhou, Y., Wu, F., & Taylor, B. V. (2016). Role of genetic susceptibility variants in predicting clinical course in multiple sclerosis: a cohort study. Journal of Neurology, Neurosurgery, and Psychiatry, 87(11), 1204–1211. 10.1136/JNNP-2016-313722

Patel, R. S., Schmidt, A. F., Tragante, V., McCubrey, R. O., Holmes, M. V., Howe, L. J., Direk, K., Åkerblom, A., Leander, K., Virani, S. S., Kaminski, K. A., Muehlschlegel, J. D., Dubé, M. P., Allayee, H., Almgren, P., Alver, M., Baranova, E. V., Behlouli, H., Boeckx, B., … Asselbergs, F.W. (2019). Association of Chromosome 9p21 With Subsequent Coronary Heart Disease Events: A GENIUS-CHD Study of Individual Participant Data. Circulation. Cardiovascular Genetics, 12(4), e002471. 10.1161/CIRCGEN.119.002471

Pirinen, M. (2023). Genetics and population analysis linemodels: clustering effects based on linear relationships. Bioinformatics, 39(3). 10.1093/bioinformatics/btad115

Prediction within Ancestral Diversity. (n.d.). Retrieved September 13, 2023, from https://opain.github.io/GenoPred/DiverseAncestry.html

Purcell, S., Neale, B., Todd-Brown, K., Thomas, L., Ferreira, M. A. R., Bender, D., Maller, J., Sklar, P., de Bakker, P. I. W., Daly, M. J., & Sham, P. C. (2007). PLINK: A Tool Set for Whole-Genome Association and Population-Based Linkage Analyses. The American Journal of Human Genetics, 81(3), 559–575. 10.1086/519795

Quality Control (QC) | Pan UKBB. (n.d.). Retrieved September 13, 2023, from https://pan-dev.ukbb.broadinstitute.org/docs/qc/index.html

Roselli, C., Chaffin, M. D., Weng, L. C., Aeschbacher, S., Ahlberg, G., Albert, C. M., Almgren, P., Alonso, A., Anderson, C. D., Aragam, K. G., Arking, D. E., Barnard, J., Bartz, T. M., Benjamin, E. J., Bihlmeyer, N. A., Bis, J. C., Bloom, H. L., Boerwinkle, E., Bottinger, E. B., … Ellinor, P. T. (2018). Multi-ethnic genome-wide association study for atrial fibrillation. Nature Genetics 2018 50:9, 50(9), 1225–1233. 10.1038/s41588-018-0133-9

Rubinacci, S., Ribeiro, D. M., Hofmeister, R. J., & Delaneau, O. (2021). Efficient phasing and imputation of low-coverage sequencing data using large reference panels. Nature Genetics 2021 53:1, 53(1), 120–126. 10.1038/s41588-020-00756-0

Schumacher, F. R., Al Olama, A. A., Berndt, S. I., Benlloch, S., Ahmed, M., Saunders, E. J., Dadaev, T., Leongamornlert, D., Anokian, E., Cieza-Borrella, C., Goh, C., Brook, M. N., Sheng, X., Fachal, L., Dennis, J., Tyrer, J., Muir, K., Lophatananon, A., Stevens, V. L., … Eeles, R. A. (2018). Association analyses of more than 140,000 men identify 63 new prostate cancer susceptibility loci.Nature Genetics 2018 50:7, 50(7), 928–936. 10.1038/s41588-018-0142-8

Shah, S., Henry, A., Roselli, C., Lin, H., Sveinbjörnsson, G., Fatemifar, G., Hedman, Å. K., Wilk, J. B., Morley, M. P., Chaffin, M. D., Helgadottir, A., Verweij, N., Dehghan, A., Almgren, P., Andersson, C., Aragam, K. G., Ärnlöv, J., Backman, J. D., Biggs, M. L., … Lumbers, R. T. (2020). Genomewide association and Mendelian randomisation analysis provide insights into the pathogenesis of heart failure. Nature Communications 2020 11:1, 11(1), 1–12. 10.1038/s41467-019-13690-5

Smith, B. H., Campbell, A., Linksted, P., Fitzpatrick, B., Jackson, C., Kerr, S. M., Deary, I. J., MacIntyre, D. J., Campbell, H., McGilchrist, M., Hocking, L. J., Wisely, L., Ford, I., Lindsay, R. S., Morton, R., Palmer, C. N. A., Dominiczak, A. F., Porteous, D. J., & Morris, A. D. (2013). Cohort Profile: Generation Scotland: Scottish Family Health Study (GS:SFHS). The study, its participants and their potential for genetic research on health and illness. International Journal of Epidemiology, 42(3), 689–700. 10.1093/IJE/DYS084

Spiliopoulou, A., Colombo, M., Orchard, P., Agakov, F., & McKeigue, P. (2017). GeneImp: Fast Imputation to Large Reference Panels Using Genotype Likelihoods from Ultralow Coverage Sequencing. Genetics, 206(1), 91–104. 10.1534/GENETICS.117.200063

Su, Z., Marchini, J., & Donnelly, P. (2011). HAPGEN2: simulation of multiple disease SNPs. Bioinformatics, 27(16), 2304–2305. 10.1093/bioinformatics/btr341

Taliun, D., Harris, D. N., Kessler, M. D., Carlson, J., Szpiech, Z. A., Torres, R., Taliun, S. A. G., Corvelo, A., Gogarten, S. M., Kang, H. M., Pitsillides, A. N., LeFaive, J., Lee, S. been Tian, X., Browning, B. L., Das, S., Emde, A. K., Clarke, W. E., Loesch, D. P., … Abecasis, G. R. (2021). Sequencing of 53,831 diverse genomes from the NHLBI TOPMed Program. Nature 2021 590:7845, 590(7845), 290–299. 10.1038/s41586-021-03205-y

Tan, M. M., Lawton, M. A., Pollard, M. I., Brown, E., Bekadar, S., Jabbari, E., Reynolds, R. H., Iwaki, H., Blauwendraat, C., Kanavou, S., Hubbard, L., Malek, N., Grosset, K. A., Bajaj, N., Barker, R. A., Burn, D. J., Bresner, C., Foltynie, T., Wood, N. W., … Morris, H. R. (2022). Genome-wide determinants of mortality and clinical progression in Parkinson’s disease. 10.1101/2022.07.07.22277297

Tcheandjieu, C., Zhu, X., Hilliard, A. T., Clarke, S. L., Napolioni, V., Ma, S., Lee, K. M., Fang, H., Chen, F., Lu, Y., Tsao, N. L., Raghavan, S., Koyama, S., Gorman, B. R., Vujkovic, M., Klarin, D., Levin, M. G., Sinnott-Armstrong, N., Wojcik, G. L., … Assimes, T. L. (2022). Large-scale genome-wide association study of coronary artery disease in genetically diverse populations. Nature Medicine 2022 28:8, 28(8), 1679–1692. 10.1038/s41591-022-01891-3

Timmers, P. R. H. J., Mounier, N., Lall, K., Fischer, K., Ning, Z., Feng, X., Bretherick, A. D., Clark, D. W., Shen, X., Esko, T., Kutalik, Z., Wilson, J. F., & Joshi, P. K. (2019). Genomics of 1 million parent lifespans implicates novel pathways and common diseases and distinguishes survival chances. ELife, 8, 1–40. 10.7554/ELIFE.39856

Turnbull, C. (2018). Introducing whole-genome sequencing into routine cancer care: the Genomics England 100 000 Genomes Project. Annals of Oncology : Official Journal of the European Society for Medical Oncology, 29(4), 784–787. 10.1093/ANNONC/MDY054

UK Biobank. (2015, October). Genotyping and quality control of UK Biobank, a large-scale, extensively phenotyped prospective resource. https://biobank.ctsu.ox.ac.uk/crystal/crystal/docs/genotyping_qc.pdf

Vandebergh, M., Andlauer, T. F. M., Zhou, Y., Mallants, K., Held, F., Aly, L., Taylor, B. V., Hemmer, B., Dubois, B., & Goris, A. (2021). Genetic Variation in WNT9B Increases Relapse Hazard in Multiple Sclerosis. Annals of Neurology, 89(5), 884–894. 10.1002/ANA.26061

Viippola, E., Kuitunen, S., Rodosthenous, R. S., Vabalas, A., Hartonen, T., Vartiainen, P., Demmler, J., Vuorinen, A.-L., Liu, A., Havulinna, A. S., Llorens, V., Detrois, K. E., Wang, F., Ferro, M., Karvanen, A., German, J., Jukarainen, S., Gracia-Tabuenca, J., Hiekkalinna, T., … Perola, M. (2023). Data Resource Profile: Nationwide registry data for high-throughput epidemiology and machine learning (FinRegistry). International Journal of Epidemiology, 52(4), e195–e200. 10.1093/IJE/DYAD091

Wightman, D. P., Jansen, I. E., Savage, J. E., Shadrin, A. A., Bahrami, S., Holland, D., Rongve, A., Børte, S., Winsvold, B. S., Drange, O. K., Martinsen, A. E., Skogholt, A. H., Willer, C., Bråthen, G., Bosnes, I., Nielsen, J. B., Fritsche, L. G., Thomas, L. F., Pedersen, L. M., … Posthuma, D. (2021). A genome-wide association study with 1,126,563 individuals identifies new risk loci for Alzheimer’s disease. Nature Genetics 2021 53:9, 53(9), 1276–1282. 10.1038/s41588-021-00921-z

Willer, C., Li, Y., & Abecasis, G. (2010). METAL: fast and efficient meta-analysis of genomewide association scans. Bioinformatics. http://bioinformatics.oxfordjournals.org/content/26/17/2190.short

World Health Organization. (2004). ICD-10 : international statistical classification of diseases and related health problems : tenth revision. https://apps.who.int/iris/handle/10665/42980

Wu, C., Kraft, P., Stolzenberg-Solomon, R., Steplowski, E., Brotzman, M., Xu, M., Mudgal, P., Amundadottir, L., Arslan, A. A., Bueno-De-Mesquita, H. B., Gross, M., Helzlsouer, K., Jacobs, E. J., Kooperberg, C., Petersen, G. M., Zheng, W., Albanes, D., Boutron-Ruault, M. C., Buring, J. E., … Wolpin, B. M. (2014). Genome-wide association study of survival in patients with pancreatic adenocarcinoma. Gut, 63(1), 152–160. 10.1136/GUTJNL-2012-303477

Wuttke, M., Li, Y., Li, M., Sieber, K. B., Feitosa, M. F., Gorski, M., Tin, A., Wang, L., Chu, A. Y., Hoppmann, A., Kirsten, H., Giri, A., Chai, J. F., Sveinbjornsson, G., Tayo, B. O., Nutile, T., Fuchsberger, C., Marten, J., Cocca, M., … Pattaro, C. (2019). A catalog of genetic loci associated with kidney function from analyses of a million individuals. Nature Genetics 2019 51:6, 51(6), 957–972. 10.1038/s41588-019-0407-x

Yaghootkar, H., Bancks, M. P., Jones, S. E., McDaid, A., Beaumont, R., Donnelly, L., Wood, A. R., Campbell, A., Tyrrell, J., Hocking, L. J., Tuke, M. A., Ruth, K. S., Pearson, E. R., Murray, A., Freathy, R. M., Munroe, P. B., Hayward, C., Palmer, C., Weedon, M. N., … Kutalik, Z. (2017). Quantifying the extent to which index event biases influence large genetic association studies. Human Molecular Genetics, 26(5), 1018–1030. 10.1093/HMG/DDW433

Zhang, H., Ahearn, T. U., Lecarpentier, J., Barnes, D., Beesley, J., Qi, G., Jiang, X., O’Mara, T. A., Zhao, N., Bolla, M. K., Dunning, A. M., Dennis, J., Wang, Q., Ful, Z. A., Aittomäki, K., Andrulis, I. L., Anton-Culver, H., Arndt, V., Aronson, K. J., … García-Closas, M. (2020). Genome-wide association study identifies 32 novel breast cancer susceptibility loci from overall and subtype-specific analyses. Nature Genetics 2020 52:6, 52(6), 572–581. 10.1038/s41588-020-0609-2

Zhang, Q., Privé, F., Vilhjálmsson, B., & Speed, D. (2021). Improved genetic prediction of complex traits from individual-level data or summary statistics. Nature Communications 2021 12:1, 12(1), 1–9. 10.1038/s41467-021-24485-y

